# Computerized Cognitive Training in People with Depression: A Systematic Review and Meta-analysis of Randomized Clinical Trials

**DOI:** 10.1101/2021.03.23.21254003

**Authors:** Eliane Gefen, Nathalie H. Launder, Christopher G. Davey, Mor Nahum, Yafit Gilboa, Carsten Finke, Hanna Malmberg Gavelin, Nicola T. Lautenschlager, Amit Lampit

## Abstract

**Importance:** Cognitive impairment is a common feature of both symptomatic and remitted states of depression that is associated with poorer psychosocial outcomes and treatment non-response. As such, finding treatments to maintain or enhance cognition in people with depression is imperative.

**Objective:** To investigate the efficacy and moderators of computerized cognitive training (CCT) for cognitive and functional outcomes in people with depression.

**Data Sources:** MEDLINE, EMBASE and PsycINFO databases were screened from inception through to 08 September 2022, with no language or publication type restrictions.

**Study Selection:** Two independent reviewers conducted duplicate study screening and assessed against the following inclusion criteria: (1) adults (mean age 18 years or older) with depression, (2) CCT with minimum three hours practice, (3) active or passive control group, (4) cognitive and/or functional outcomes measured at baseline and post-intervention, (5) randomized controlled trials. Of 4245 identified studies, 34 met selection criteria.

**Data Extraction and Synthesis:** The methods used followed the Preferred Reporting Items for Systematic Reviews and Meta-Analyses (PRISMA) guidelines. Data extraction and risk of bias assessment using the revised Cochrane Risk of Bias Tool (RoB2) was conducted independently by two reviewers. Analyses were conducted using robust variance estimation.

**Outcomes:** The primary outcome was change from baseline to post-intervention in overall cognition. Secondary outcomes were depressive symptoms, psychiatric symptoms, psychosocial functioning, daily functioning, subjective cognition, global cognition and domain-specific cognitive function.

**Results:** Thirty-four studies encompassing 39 comparisons and 2041 unique participants met inclusion criteria. The pooled effect size of CCT was small for both overall cognition (*g*=0.28; 95% CI 0.17 to 0.38; *P*<.001; τ^2^=0.078; *I*^2^=47%; 95% prediction interval −0.31 to 0.86) and depressive symptoms (*g*=0.23; 95% CI 0.08 to 0.39; *P*=.004; τ^2^=0.066; *I*^2^=45%; 95% prediction interval −0.32 to 0.78). Benefits of CCT were also found for psychosocial functioning, subjective cognition, fluid reasoning, long-term memory and retrieval, low working memory, shifting, inhibition and processing speed. Greater CCT dose and multidomain programs were associated with greater cognitive response to CCT. There was no evidence for difference across clinical subtypes or between delivery modalities.

**Conclusions and Relevance:** This systematic review and meta-analysis indicates that CCT is an efficacious intervention for overall cognition, depressive symptoms, psychosocial functioning, subjective cognition, and many domain-specific cognitive functions for people with depression.

## Introduction

Cognitive impairment is a central feature of depression,^1^ presented frequently in both symptomatic and remitted states, associated with poorer psychosocial functioning^2^ and treatment non-response^3–5^ and is only partially responsive to antidepressants.^6^ Comorbid depression and cognitive impairment are common in people with chronic diseases and may interfere with the management of medical disorders and treatment adherence, leading to worse functional and medical outcomes.^7^ Moreover, as one of the most robust dementia risk factors, depression increases the risk of dementia in later life by approximately 80%^8^ and given its prevalence may independently account for around 8% of dementia cases worldwide.^9^ Thus, interventions that effectively target cognition alongside other symptoms in people with depression may have a key role in supporting everyday function,^10^ as well as in delaying or preventing cognitive decline and dementia.^8,11^

Computerized cognitive training (CCT) is a safe and scalable cognitive training approach that focuses on repeated and controlled practice on cognitively demanding tasks. CCT is appealing as it can be adapted to individual needs, provides ongoing feedback, is relatively inexpensive and can be delivered flexibly. CCT is arguably the most common intervention in cognitive impairment trials delivered as a standalone or in combination with other approaches such as physical exercise^12^ and cognitive remediation,^13^ with robust evidence for efficacy in ageing,^14,15^ neurodegenerative^12,16^ and psychiatric disorders.^13,17^ However, efficacy for specific outcomes varies across populations, and associated with intervention design factors such as content, dose and delivery. Indirect evidence from meta-analyses in older adults^14^ and schizophrenia^13,18^ suggests that CCT may be efficacious only when combined with behavioral or cognitive remediation techniques.

Several recent meta-analyses investigated the efficacy of cognitive remediation in people with depression, reporting improvements in depressive symptoms and mixed results for objective and subjective cognitive outcomes.^19–23^ However, all of these combined CCT with other cognitive remediation techniques that did not include CCT, only one^19^ was limited to randomized controlled trials (RCTs), and no meta-analysis accounted for non-independence of effect sizes within studies, potentially over-estimating effect estimates in under-estimating heterogeneity.^24^ Moreover, nearly all the studies included in previous meta-analyses excluded older adults and people with comorbidities, thereby limiting the applicability of results to clinical practice.

Therefore we aimed to robustly estimate the efficacy and heterogeneity of CCT as a standalone or component intervention on cognitive, mood and psychosocial outcomes across populations with depression, and to investigate factors associated with response – key evidence gap to guide clinical implementation.^10^

## Methods

This review adheres to the 2020 Preferred Reporting Items for Systematic Reviews and Meta-Analyses (PRISMA 2020) guidelines^25^ and largely follows methods established in our previous reviews of CCT.^14,15,26,27^ The protocol has been prospectively registered with PROSPERO (CRD42020204209) and published previously.^28^

### Eligibility Criteria

We included RCTs studying the effects of CCT compared to control conditions on one or more cognitive, depressive symptoms, psychosocial or functional outcome(s) in adults with depression at baseline (at any clinical stage). Depression was established according to standard diagnostic criteria, diagnostic interviews, expert clinical diagnosis or a mean score greater than a validated cut-off on an established clinical measure (**eTable 1** in the Supplement). There was no restriction on study population apart from studies targeting primarily people with dementia or major psychiatric comorbidities; when the study population included a mixed sample (e.g., as indicated by baseline demographics or when ≥50% of the sample received antipsychotic medication), the study was only included if data of eligible participants could be obtained separately. CCT was defined as a total of minimum of 3 hours of intended practice on standardized computerized tasks or video games with clear cognitive rationale.^14,15,27^ Eligible controls included passive (wait-list, no-contact) and active (e.g., sham CCT, recreational activities) comparison groups. Studies combining CCT with other non-pharmacological interventions (e.g., psychotherapy, physical exercise) or with pharmacological interventions were eligible as long as both arms received the same adjacent interventions (i.e., the difference between the arms is the CCT and not the adjacent intervention). All eligible comparisons in multi-arm studies were included using multivariate models.

### Information Sources and Study Selection

MEDLINE, EMBASE and PsycINFO were searched through the OVID interface for eligible articles from inception through to 8 September 2022. No restrictions on language or type of publication were applied. The electronic search was complemented by hand-searching the references of included studies and previous reviews of CCT^14–16,27^ and cognitive remediation^19–21^ as well as clinical trial registries. The full search strategy is shown in **eTable 2** in the Supplement. Three independent reviewers (NHL, EG and MN) conducted duplicate screening of titles and abstracts as well as full text screening of potentially eligible articles. Disagreements at each stage were resolved by consensus or by involvement of a senior reviewer (AL), who also contacted the corresponding authors of primary studies for additional information. The final list of included studies was reviewed and approved by AL.

### Data Extraction and Coding

Data were extracted and coded in duplicate by two reviewers (NHL, RM or EG), supervised by a neuropsychologist (HMG). Outcome data were extracted as mean and standard deviation (SD) for each group at each time point, or when not available, as measures of mean difference and SD or confidence intervals (CI). Missing or incomplete data were requested from the corresponding authors of the studies. Coding of cognitive outcomes was conducted according to the Cattell-Horn-Carroll-Miyake (CHC-M) framework, tailored specifically for meta-analyses of CCT.^29^ Following this framework, each cognitive outcome was classified into a broad cognitive domain (e.g., executive function) as well as a more specific narrow cognitive domain (e.g., inhibition). Cognitive screening instruments, such as the Mini-Mental State Examination, were classified as global cognition.^14,15^ The classification of individual outcome measures into domains is presented in **eTable 3** in the Supplement. Non-cognitive outcomes included depressive symptoms, psychiatric symptoms, psychosocial functioning, daily functioning, and subjective cognition. The classification of baseline depressive symptoms severity for subgroup analysis is presented in **eTable 4** in the Supplement.

### Risk of Bias Within Studies

Three independent reviewers (NHL, EG and RM) assessed the risk of bias of eligible comparisons within studies using the revised Cochrane Risk of Bias Tool (RoB2).^30^ Disagreements were resolved by consensus or by consultation with a senior reviewer (AL). In contrast to the original RoB2 macros, studies with “some concerns” or “high” risk of bias in domains 3 (bias due to missing outcome data) or 4 (bias in measurement of the outcome) were considered as having some concerns or high risk of bias, respectively.^27^

### Data Synthesis

Analyses were conducted using the packages robumeta,^31^ clubSandwich^32^ and metafor^33^in R, version 4.2.2. Between-group differences in change from baseline to post-intervention were converted to standardized mean differences and calculated as Hedges’ *g* with 95% CI for each eligible outcome measure. Multivariate analyses were performed using robust variance estimation (RVE) based on a correlational model with rho=0.8 to account for the non-independence of multiple effect sizes within studies.^34^ The primary outcome was change from baseline to post-intervention in overall cognition, assessed through one or more non-trained measures of objective cognition using standardized neuropsychological tests. Secondary outcomes included depressive symptoms, psychiatric symptoms, psychosocial functioning, daily functioning, subjective cognition, global cognition, and domain-specific cognitive function.

Heterogeneity across studies was quantified using τ^2^ and expressed as a proportion of overall observed variance using the *I*^2^ statistic.^35,36^ Prediction intervals were calculated to assess the dispersion of true effects across settings.^37^ Univariable RVE meta-regressions of a priori potential moderators (design characteristics, population characteristics and overall risk of bias) were performed for overall cognition and depressive symptoms using robumeta and contrasts formally tested using Hotelling–Zhang test (F-statistic) with clubSandwich.^38^ Small-study effect for primary outcomes was assessed by visually inspecting funnel plots of effect size vs standard error^39^ and formally tested using the Egger’s test as a meta-regression in RVE.^40^ The Duval and Tweedie trim and fill^41^ was also used to assess the magnitude of small-study bias. Two-sided α<.05 indicated statistical significance.

## Results

### Study Selection

After removal of duplicates, we screened 4927 articles for eligibility, of which 657 articles were assessed in full-text screening. A total of 34 studies were found eligible for inclusion (**Figure 1**). A list of studies excluded at the full text screening stage is provided in **eTable 5** in the Supplement. Additionally, the authors of ten eligible studies were contacted for additional data, of which five^42–45^ provided data.

**Figure 1:**
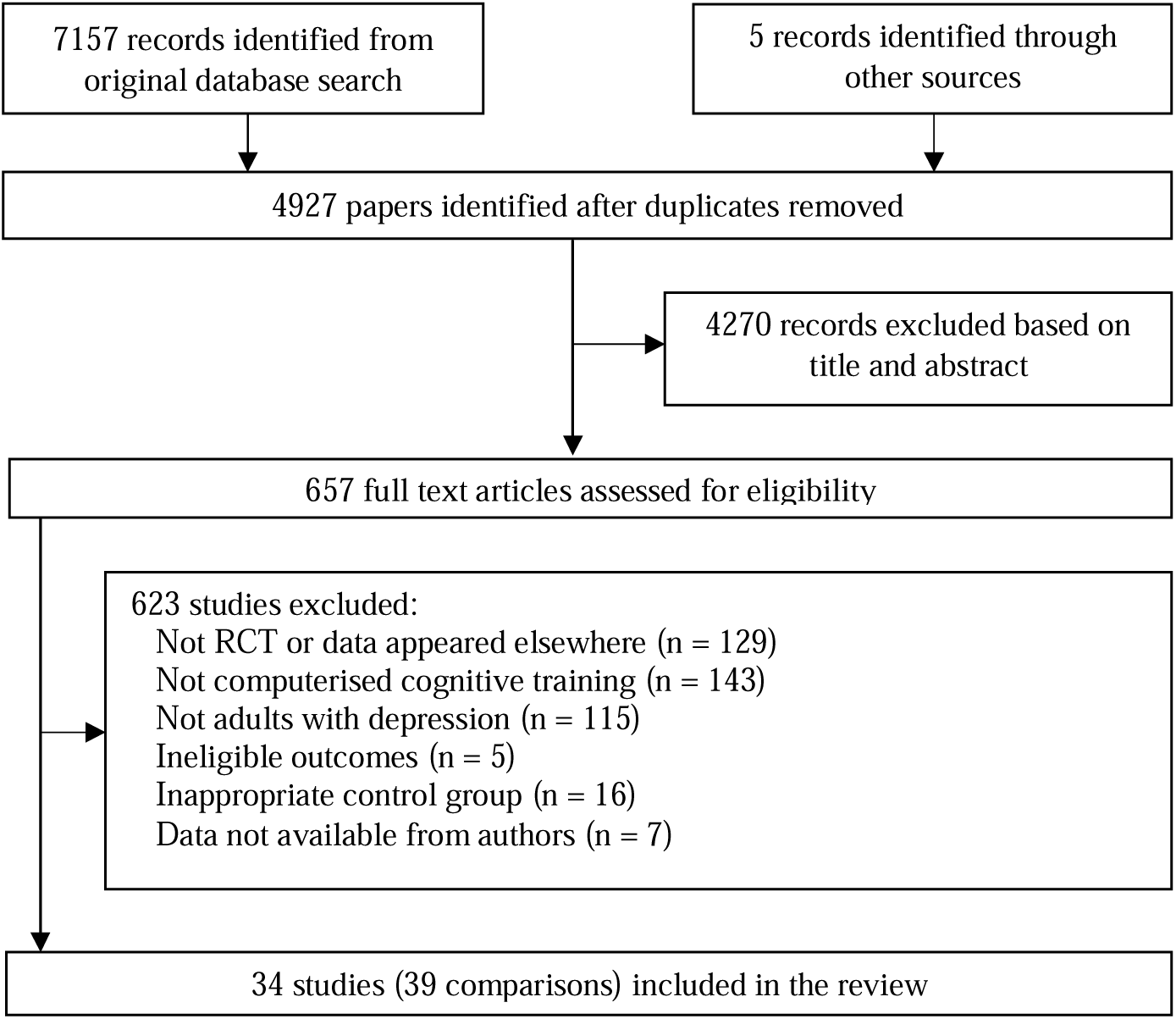
Flowchart of study selection.

### Characteristics of Included Studies

The 34 included studies reported data from 39 eligible comparisons, encompassing 2041 unique participants, with mean age ranging between 19.21 and 74.51 years. Most of the studies included people with a diagnosis of MDD or a current major depressive episode (*k=*13 RCTs; *n*=813). Four studies^46–49^ specifically focused on people with partially and/or fully remitted depression (*n*=207). Seven studies included people with multiple sclerosis^50–56^ (n=360). Three studies included people with Parkinson’s disease (n=153) ^57,58^ and two studies included people with MCI/SCD (*n*=65). Five studies (*n*=188) targeted older adults with the mean age of participants over 65 years of age.^42,57–60^ Mean baseline depression severity was classified as mild depression (*k*=18; *n*=1049) or moderate to severe depression (*k*=16; *n*=992) according to the standard severity cut-offs for the clinical measures used. Studies including participants with fully remitted depression that had mean baseline depression severities below cut-offs^46,47^ were included in the mild depression subgroup. The most common type of CCT was multidomain training (*k*=22*; n*=1083), followed by working memory training (*k*=7*; n*=443), attention training (*k=2; n*=367), memory training (*k*=2*; n*=75) and speed of processing training (*k*=1*; n*=73). Two studies had two CCT arms (*n*=110).^48,61^ Eighteen studies used an active control group (*n*=970), 13 studies had a passive control group (*n*=598), one study had two active control groups (*n*=279)^44^ and two studies had both active and passive control groups (*n*=194).^62^ Overall risk of bias was assessed as low in 15 studies, with some concerns in seven studies, and high risk of bias in 11 studies. Finally, one study^62^ had one comparison assessed as low and the other as high (**eTable 6** in the Supplement).

### Primary Outcome: Overall Cognition

Thirty-one studies reported objective cognitive outcomes at baseline and post-intervention timepoints. The pooled effect size across these 31 studies, with a total of 304 cognitive outcomes, was small and statistically significant with moderate heterogeneity (*g*=0.28; 95% CI 0.17 to 0.38; *P*<.001; τ^2^=0.078; *I*^2^=47%; prediction interval −0.31 to 0.86, **Figure 2**). Funnel plot asymmetry was detected, indicating possible small-study effect (β=-0.105; one-tailed *P*=.03; **eFigure 1** in the Supplement). A trim and fill analysis imputed two studies; the adjusted effect size suggested negligible small-study bias (*g*=0.24; 95% CI 0.14 to 0.34; **eFigure 2** in the Supplement). Sensitivity analyses comparing a hierarchical (*g*=0.22; 95% CI 0.10 to 0.34; *P*=.001; W^2^=0.018; τ^2^=0.061) to the correlational model as well as correlation assumptions revealed the model assumptions of the main analysis to be robust (**eTable 7** in the Supplement). An additional sensitivity analysis excluding studies of remitted depression populations (*k*=2; *n*=78)^47,48^ was also consistent with the overall result (*g*=0.28; 95% CI 0.18 to 0.39; *P*<.001; τ^2^=0.074; *I*^2^=46%). The pooled effect size was similar across active- and passive-controlled comparisons and larger in studies with low risk of bias, but the difference was not statistically significant (**Table 2**). However, trial registration was associated with smaller effect sizes compared to non-registered trials.

**Figure 2:**
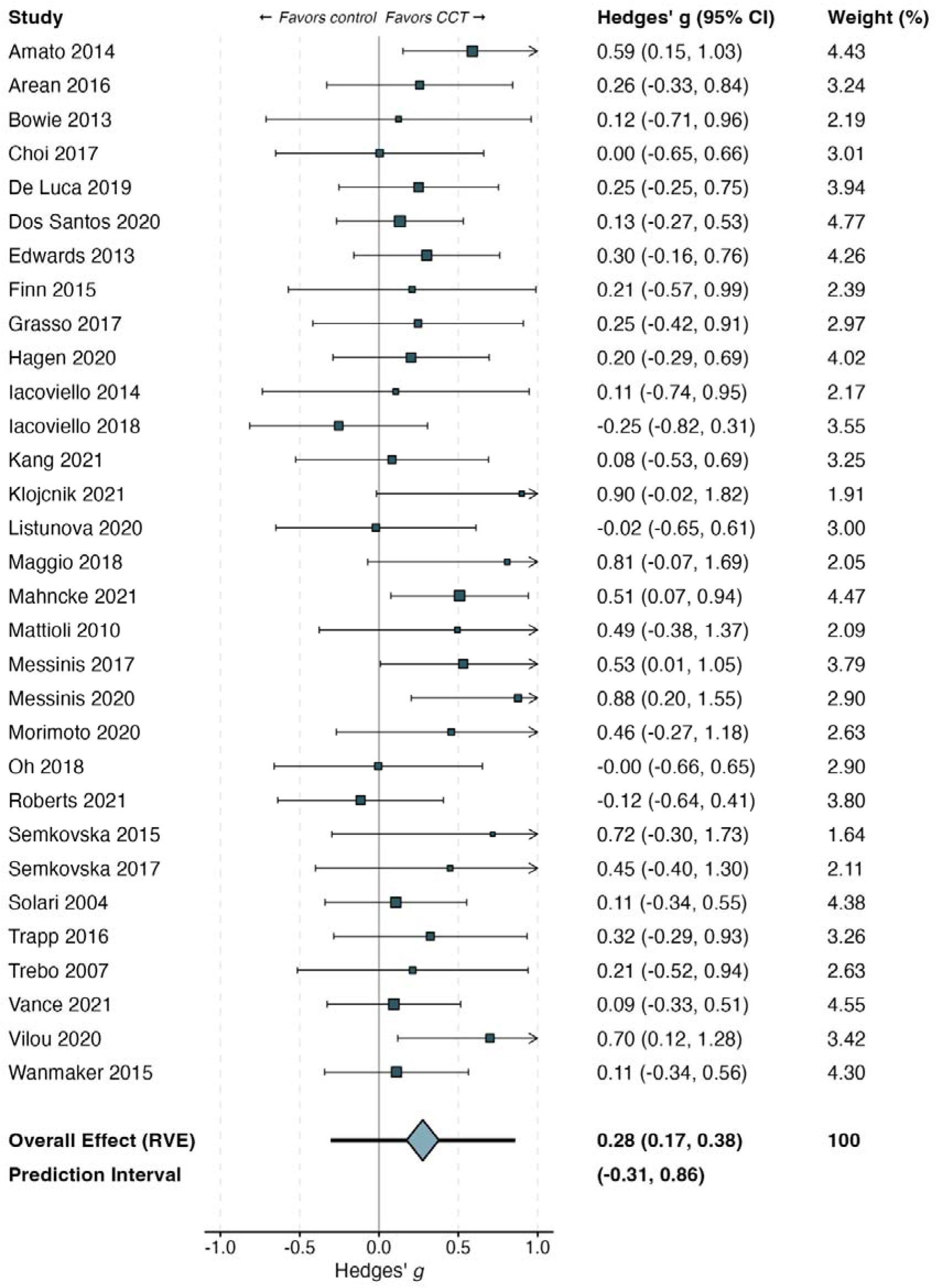
Meta-analysis of overall cognitive outcomes.

**Table 1:** Study characteristics.

| Study | n<sup>a</sup> | % fem | Mean age<sup>b</sup> | Depression severity | Clinical population<sup>c</sup> | Interventions | CCT type | Delivery | Dose | Risk of bias |
| --- | --- | --- | --- | --- | --- | --- | --- | --- | --- | --- |
| Amato 2014<sup>52</sup> | 88 (CCT: 55; Control: 33) | 78 | 40.96 | Mild | Multiple sclerosis | CCT: Attention Processing Training (ST)<br>Control: Non-specific computerized activities (n-ST) | Attention | Home | Dose: 24 h<br>Session: 60 min<br>Frequency: 2<br>Weeks: 12 | Some concerns |
| Arean 2016<sup>44</sup> | 76<sup>d</sup> (CCT: 51; Control: 25); 179<sup>e</sup> (CCT: 100; Control: 79) | 76 | 34.15 | Moderate-severe | MDD | CCT: Project: EVO (EVO)<br>Control: Problem-solving therapy app (iPST) | Attention | Home | Dose: 12 h<br>Session: 30 min<br>Frequency: 6<br>Weeks: 4 | High |
| Arean 2016<sup>44</sup> | 71<sup>d</sup> (CCT: 51; Control: 20); 200<sup>e</sup> (CCT: 100; Control: 100) | 80 | 34.25 | Moderate-severe | MDD | CCT: Project: EVO (EVO)<br>Control: Health tips app (HT) | Attention | Home | Dose: 12 h<br>Session: 30 min<br>Frequency: 6<br>Weeks: 4 | High |
| Bowie 2013<sup>45</sup> | 21 (CCT: 11; Control: 10) | 70 | 45.81 | Moderate-severe | MDD | CCT: Scientific Brain Training Pro (CR)<br>Control: Wait-list (WL) | Multidomain | Supervised | Dose: 9 h<br>Session: 54 min<br>Frequency: 1<br>Weeks: 10 | Low |
| Choi 2017<sup>62</sup> | 33 (CCT: 18; Control: 15) | 51 | 43.42 | Moderate-severe | MDD | CCT: In-house program (Mem-ECT)<br>Control: Computer and pencil-and-paper puzzles (AC) | Memory | Supervised | Dose: 7.88 h<br>Session: 67.5 min<br>Frequency: 2<br>Weeks: 3 | High |
| Choi 2017<sup>62</sup> | 36 (CCT: 18; Control: 18) | 59 | 40.47 | Moderate-severe | MDD | CCT: In-house program (Mem-ECT)<br>Control: Treatment as usual (TAU) | Memory | Supervised | Dose: 7.88 h<br>Session: 67.5 min<br>Frequency: 2<br>Weeks: 3 | Low |
| De Luca 2019<sup>66</sup> | 60 (CCT: 30; Control: 30) | 48 | 62.55 | Mild | Parkinson's disease | CCT: ERICA (EG)<br>Control: Pencil-and-paper cognitive training with therapist (CG) | Multidomain | Supervised | Dose: 24 h<br>Session: 60 min<br>Frequency: 3<br>Weeks: 8 | Some concerns |
| Dos Santos 2020<sup>67</sup> | 92 (CCT: 48; Control: 44) | 96 | 51.30 | Moderate-severe | Cancer-related cognitive impairment | CCT: Computer-assisted cognitive rehabilitation with a neuropsychologist (RehaCom)<br>Control: Pencil-and-paper cognitive exercises at home | Multidomain | Supervised | Dose: 7.88 h<br>Session: 52.5 min<br>Frequency: 0.75<br>Weeks: 12 | High |
| Dos Santos 2020<sup>67</sup> | 99 (CCT: 48; Control: 51) | 96 | 51.20 | Moderate-severe | Cancer-related cognitive impairment | CCT: Computer-assisted cognitive rehabilitation with a neuropsychologist (RehaCom)<br>Control: Phone follow-up | Multidomain | Supervised | Dose: 7.88 h<br>Session: 52.5 min<br>Frequency: 0.75<br>Weeks: 12 | High |
| Edwards 2013<sup>57</sup> | 73 (CCT: 32; Control: 41) | 38 | 68.85 | Mild | Parkinson's disease | CCT: InSight (SOPT)<br>Control: Wait-list (CG) | Speed of processing | Home | Dose: 20 h<br>Session: 60 min<br>Frequency: 2<br>Weeks: 12 | High |
| Ferrari 2021<sup>68</sup> | 115 (CCT: 56; Control: 59) | 63 | 51.31 | Moderate-severe | MDD | CCT: In-house program (adaptive PASAT)<br>Control: Sham PASAT | Working memory | Supervised | Dose: 4.17 h<br>Session: 25 min<br>Frequency: 5<br>Weeks: 2 | Low |
| Finn 2015<sup>59</sup> | 24 (CCT: 12; Control: 12) | 29 | 73.96 | Mild | MCI | CCT: In-house program (repetition lag training)<br>Control: No-contact | Memory | Supervised | Dose: 9 h<br>Session: 90 min<br>Frequency: 2<br>Weeks: 4 | Some concerns |
| Grasso 2017<sup>53</sup> | 34 (CCT: 17; Control: 17) | 65 | 59.11 | Moderate-severe | Multiple sclerosis | CCT: Attention Processing Training (CMD)<br>Control: Non-specific computerized activities (MD) | Multidomain | Supervised | Dose: 36 h<br>Session: 60 min<br>Frequency: 3<br>Weeks: 12 | Low |
| Hagen 2020<sup>69</sup> | 63 (CCT: 28; Control: 35) | 78 | 42.00 | Moderate-severe | MDD | CCT: BrainHQ<br>Control: Goal management training (GMT) | Multidomain | Supervised | Dose: 9 h<br>Session: 60 min<br>Frequency: 2<br>Weeks: 4.5 | High |
| Hoorelbeke 2017<sup>46</sup> | 61 (CCT: 29; Control: 32) | 66 | 46.97 | Mild | Remitted MDD | CCT: In-house program (adaptive PASAT)<br>Control: Sham PASAT | Working memory | Home | Dose: 3.33 h<br>Session: 20 min<br>Frequency: 5<br>Weeks: 2 | High |
| Iacoviello 2014<sup>70</sup> | 21 (CCT: 11; Control: 10) | 52 | 37.84 | Moderate-severe | MDD | CCT: In-house program (Emotional Faces Memory Task training)<br>Control: Sham CCT (CT) | Working memory | Supervised | Dose: 5 h<br>Session: 37.5 min<br>Frequency: 2<br>Weeks: 4 | Low |
| Iacoviello 2018<sup>71</sup> | 48 (CCT: 26; Control: 22) | 69 | 35.04 | Moderate-severe | MDD | CCT: In-house program (Emotional Faces Memory Task training)<br>Control: Sham CCT (CT) | Working memory | Supervised | Dose: 8.25 h<br>Session: 27.5 min<br>Frequency: 3<br>Weeks: 6 | High |
| Kang 2021<sup>60</sup> | 41 (CCT: 23; Control: 18) | 71 | 74.51 | Mild | Older adults with subjective cognitive decline or MCI | CCT: In-house program (VR cognitive training)<br>Control: Treatment as usual (TAU) | Multidomain | Supervised | Dose: 3.33 h<br>Session: 25 min<br>Frequency: 2<br>Weeks: 4 | Some concerns |
| Klojcnik 2021<sup>72</sup> | 20 (CCT: 10; Control: 10) | 55 | 45.30 | Moderate-severe | MDD | CCT: CogniPlus (CCRT)<br>Control: Treatment as usual (TAU) | Multidomain | Supervised | Dose: 9 h<br>Session: 45 min<br>Frequency: 1.2<br>Weeks: 10 | Some concerns |
| Listunova 2020<sup>48</sup> | 39 (CCT: 20; Control: 19) | 72 | 45.41 | Mild | Partially remitted MDD | CCT: CogniPlus individualized training (IT)<br>Control: No-contact (CG) | Multidomain | Supervised | Dose: 13.5 h<br>Session: 60 min<br>Frequency: 3<br>Weeks: 5 | Low |
| Listunova 2020<sup>48</sup> | 37 (CCT: 18; Control: 19) | 73 | 45.10 | Mild | Partially remitted MDD | CCT: CogniPlus generalized training (GT)<br>Control: No-contact (CG) | Multidomain | Supervised | Dose: 13.5 h<br>Session: 60 min<br>Frequency: 3<br>Weeks: 5 | Low |
| Maggio 2018<sup>58</sup> | 20 (CCT: 10; Control: 10) | 50 | 69.40 | Mild | Parkinson's disease | CCT: BTS Nirvana<br>Control: Pencil-and-paper cognitive training with therapist | Multidomain | Supervised | Dose: 24 h<br>Session: 60 min<br>Frequency: 3<br>Weeks: 8 | Low |
| Mahnke 2021<sup>73</sup> | 83 (CCT:41; Control: 42) | 19 | 33.8 | Mild | mTBI | CCT: BrainHQ<br>Control: Computer games | Multidomain | Supervised | Dose: 60 h<br>Session: 60 min<br>Frequency: 5<br>Weeks: 12 | Low |
| Mattioli 2010<sup>54</sup> | 20 (CCT: 10; Control: 10) | 100 | 45.16 | Mild | Multiple sclerosis | CCT: RehaCom (SG)<br>Control: No-contact (CG) | Multidomain | Supervised | Dose: 36 h<br>Session: 60 min<br>Frequency: 3 | High |
|  |  |  |  |  |  |  |  |  | Weeks: 12 |  |
| Messinis 2017<sup>51</sup> | 58 (CCT: 32; Control: 26) | 69 | 45.64 | Mild | Multiple sclerosis | CCT: RehaCom<br>Control: No-contact (CG) | Multidomain | Supervised | Dose: 20 h<br>Session: 60 min<br>Frequency: 2<br>Weeks: 10 | Low |
| Messinis 2020<sup>55</sup> | 36 (CCT: 19; Control: 17) | 67 | 45.91 | Mild | Multiple sclerosis | CCT: RehaCom<br>Control: Sham cognitive intervention (non-specific computer-based activities) | Multidomain | Home | Dose: 18 h<br>Session: 45 min<br>Frequency: 3<br>Weeks: 8 | Low |
| Morimoto 2020<sup>42</sup> | 30 (CCT: 18; Control: 12) | 64 | 73.70 | Moderate-severe | Older adults with MDD | CCT: BrainHQ and in-house program (nCCR)<br>Control: Watching documentaries and answering questions about them (AC) | Multidomain | Supervised | Dose: 30 h<br>Session: 150 min<br>Frequency: 3<br>Weeks: 4 | High |
| Oh 2018<sup>61</sup> | 34 (CCT: 18; Control: 16) | 53 | 59.59 | Mild | Subjective memory complaints | CCT: SMART<br>Control: Wait-list (WL) | Multidomain | Home | Dose: 11.67 h<br>Session: 17.5 min<br>Frequency: 5<br>Weeks: 8 | Some concerns |
| Oh 2018<sup>61</sup> | 35 (CCT: 19; Control: 16) | 54 | 59.32 | Mild | Subjective memory complaints | CCT: Fit Brains<br>Control: Wait-list (WL) | Multidomain | Home | Dose: 11.67 h<br>Session: 17.5 min<br>Frequency: 5<br>Weeks: 8 | Some concerns |
| Roberts 2021<sup>74</sup> | 55 (CCT: 26; Control: 29) | NR | 19.21 | Mild | Young adults | CCT: In-house program (adaptive working memory updating training)<br>Control: Non-adaptive working memory updating training | Working memory | Home | Dose: 10 h<br>Session: 30 min<br>Frequency: 5<br>Weeks: 4 | High |
| Semkovska 2015<sup>43</sup> | 15 (CCT: 8; Control: 7) | 46 | 43.40 | Moderate-severe | MDD | CCT: RehaCom (NCRT)<br>Control: Computer games | Multidomain | Supervised | Dose: 20 h<br>Session: 60 min<br>Frequency: 3<br>Weeks: 6.67 | High |
| Semkovska 2017<sup>47</sup> | 21 (CCT: 11; Control: 10) | 82 | 46.40 | Mild | Remitted MDD | CCT: RehaCom (NCRT)<br>Control: Computer games | Multidomain | Home | Dose: 20 h<br>Session: 60 min<br>Frequency: 4<br>Weeks: 5 | Low |
| Solari 2004<sup>50</sup> | 77 (CCT: 40; Control: 37) | 64 | 43.80 | Moderate-severe | Multiple sclerosis | CCT: RehaCom (SG)<br>Control: RehaCom visuo-constructional and visuo-motor coordination retraining (CG) | Multidomain | Supervised | Dose: 12 h<br>Session: 45 min<br>Frequency: 2<br>Weeks: 8 | Low |
| Trapp 2016<sup>75</sup> | 41 (CCT: 21; Control: 20) | 67 | 35.57 | Moderate-severe | MDD | CCT: X-Cog (EG)<br>Control: Treatment as usual (CG) | Multidomain | Supervised | Dose: 12 h<br>Session: 60 min<br>Frequency: 3<br>Weeks: 4 | Low |
| Trebo 2007<sup>76</sup> | 34 (CCT: 24; Control: 10) | 71 | 51.72 | Moderate-severe | MDD | CCT: COGPACK<br>Control: No-contact (CG) | Multidomain | Supervised | Dose: 12.5 h<br>Session: 37.5 min<br>Frequency: 2<br>Weeks: 10 | High |
| Vance 2021<sup>77</sup> | 109 (CCT: 64; Control: 45) | 30.27 | 53.56 | Mild | HIV | CCT: BrainHQ<br>Control: Passive | Multidomain | Supervised | Dose: 20 h<br>Session: 99.6 min<br>Frequency: 1<br>Weeks: 12 | High |
| Vervake 2021<sup>49</sup> | 68 (CCT: 34; Control: 34) | 64.7 | 46.2 | Mild | Remitted MDD | CCT: aPASAT<br>Control: speed-of-response training task | Working Memory | Home | Dose: 2.5 h<br>Session: 15 min<br>Frequency: ?<br>Weeks: 2 | Low |
| Vilou 2020<sup>56</sup> | 47 (CCT: 23; Control: 24) | 85 | 35.70 | Mild | Multiple sclerosis | CCT: BrainHQ<br>Control: Treatment as usual (TAU) | Multidomain | Home | Dose: 8 h<br>Session: 40 min<br>Frequency: 2<br>Weeks: 6 | Low |
| Wanmaker 2015<sup>78</sup> | 75 (CCT: 36; Control: 39) | 49 | 47.03 | Mild | MDD | CCT: In-house program (EG)<br>Control: Non-adaptive working memory training (PG) | Working memory | Home | Dose: 10 h<br>Session: 25 min<br>Frequency: 6<br>Weeks: 4 | Some concerns |

Abbreviations: CCT = computerized cognitive training. MDD = major depressive disorder. MCI = mild cognitive impairment. NR = not reported.
<sup>a</sup> Sample size used in analysis
<sup>b</sup> Weighted mean age for CCT and control groups
<sup>c</sup> Clinical populations specified in original studies
<sup>d</sup> Mean sample size for cognitive outcomes
<sup>e</sup> Sample size for depressive symptoms outcome

**Table 2:**
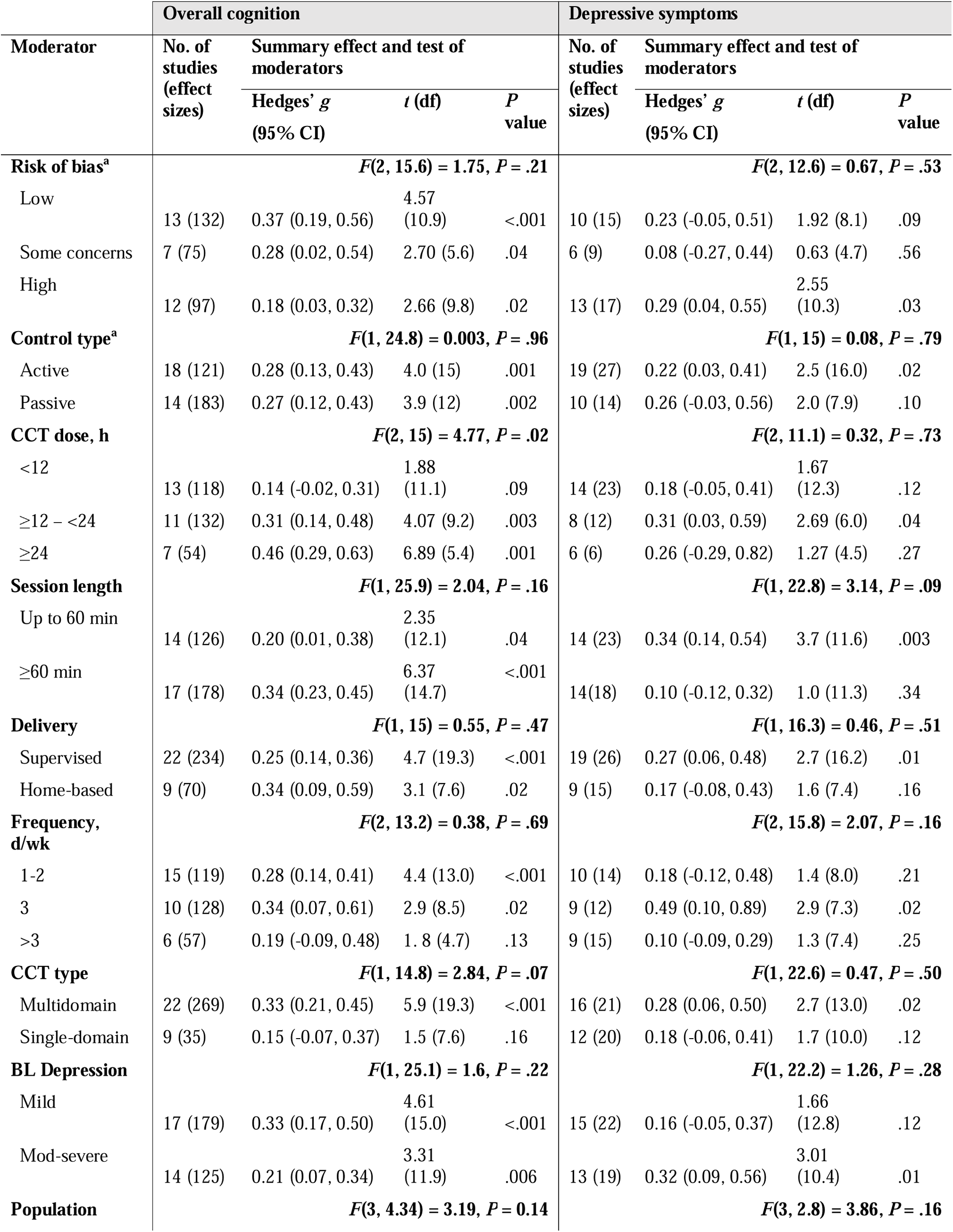
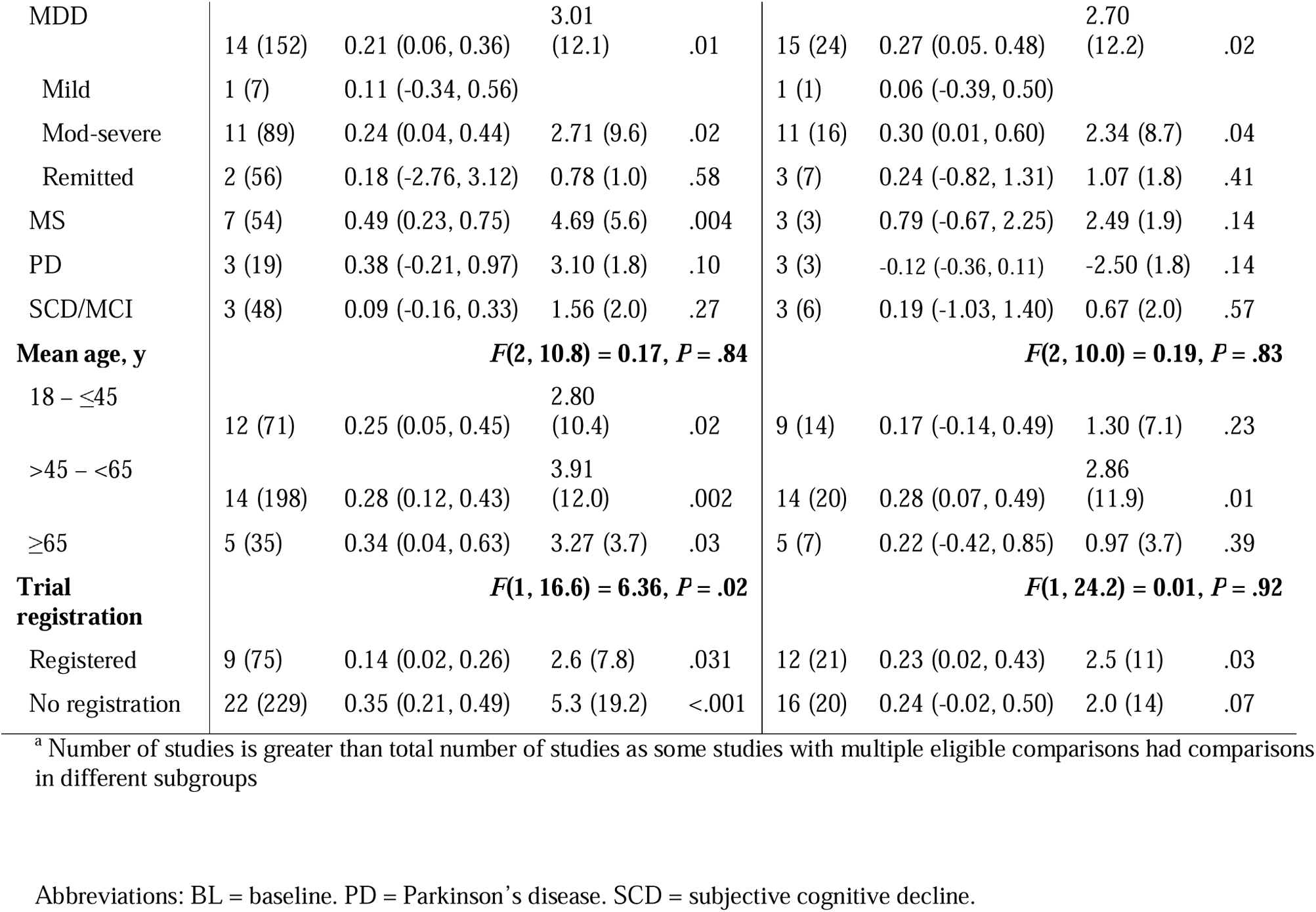
Results of meta-regressions.

### Secondary Outcome: Depressive Symptoms

Twenty-eight studies reported depressive symptoms outcomes at baseline and post-intervention timepoints. The pooled effect size across these 28 studies and 41 effect sizes was small and statistically significant with moderate heterogeneity (*g*=0.23; 95% CI 0.08 to 0.38; *P*=.004; τ^2^=0.066; *I*^2^=45%; prediction interval −0.32 to 0.78). Funnel plot asymmetry was detected, indicating possible small-study effect (β= −0.231; *P*=.08; **eFigure 4** in the Supplement). A trim and fill analysis imputed three studies, with the adjusted effect size suggesting minor small-study bias (*g*=0.17; 95% CI 0.01 to 0.32; **eFigure 4** in the Supplement). Sensitivity analyses comparing a hierarchal (*g*=0.24; 95% CI 0.08 to 0.39; *P*=.004; W^2^=0.0; τ^2^=0.057) to the correlational model as well as correlation assumptions revealed the model assumptions of the main analysis to be robust (**eTable 7** in the Supplement). An additional sensitivity analysis excluding studies of remitted depression populations (*k*=2; *n*=78) further supported this (*g*=0.23; 95% CI 0.07 to 0.39; *P*=.008; τ^2^=0.073; *I*^2^=48%). An additional sensitivity analysis excluding studies of remitted depression populations (*k*=3, *n*=150)^46,47,49^ further supported this (*g*=0.23; 95% CI 0.07 to 0.39; *P*=.008; τ^2^=0.073; *I*^2^=48%). The pooled effect size was similar across studies with high and low risk of bias, as well as across active- and passive-controlled comparisons (**Table 2**).

### Moderator analyses

Results of meta-regressions for the key outcomes of overall cognition and depressive symptoms are provided in **Table 2**. Greater dose (i.e., more training hours) was associated with larger cognitive effect sizes, in patterns suggestive of dose-responsiveness. Of particular importance, training regimes of less than 12 hours was the most common design (13 of the 31 studies reporting cognitive outcomes) and was associated with negligible cognitive benefits. No association was found between population parameters (diagnosis, baseline depression level and age) and CCT benefits.

### Secondary Outcomes: Non-cognitive Endpoints

Analyses of non-cognitive outcomes are provided in eFigures 3 to 11. Small and statistically significant effect sizes were noted for measures of psychosocial functioning (*k*=14; *g*=0.20; 95% CI 0.03 to 0.37; *P*=.03; τ^2^=0.022; *I*^2^=21%) and subjective cognition (*k*=11; *g*=0.21; 95% CI 0.02 to 0.40; *P*=.03; τ^2^=0.019; *I*^2^=20%). Negligible effects sizes were found for psychiatric symptoms (*k*=13; *g*=0.15; 95% CI −0.01 to 0.32; *P*=.07; τ^2^=0.027; *I*^2^=27%), daily function (*k*=5; *g*=0.11; 95% CI −0.19 to 0.51; *P*=.25; τ^2^=0.01; *I*^2^=27%),

**Figure 3:**
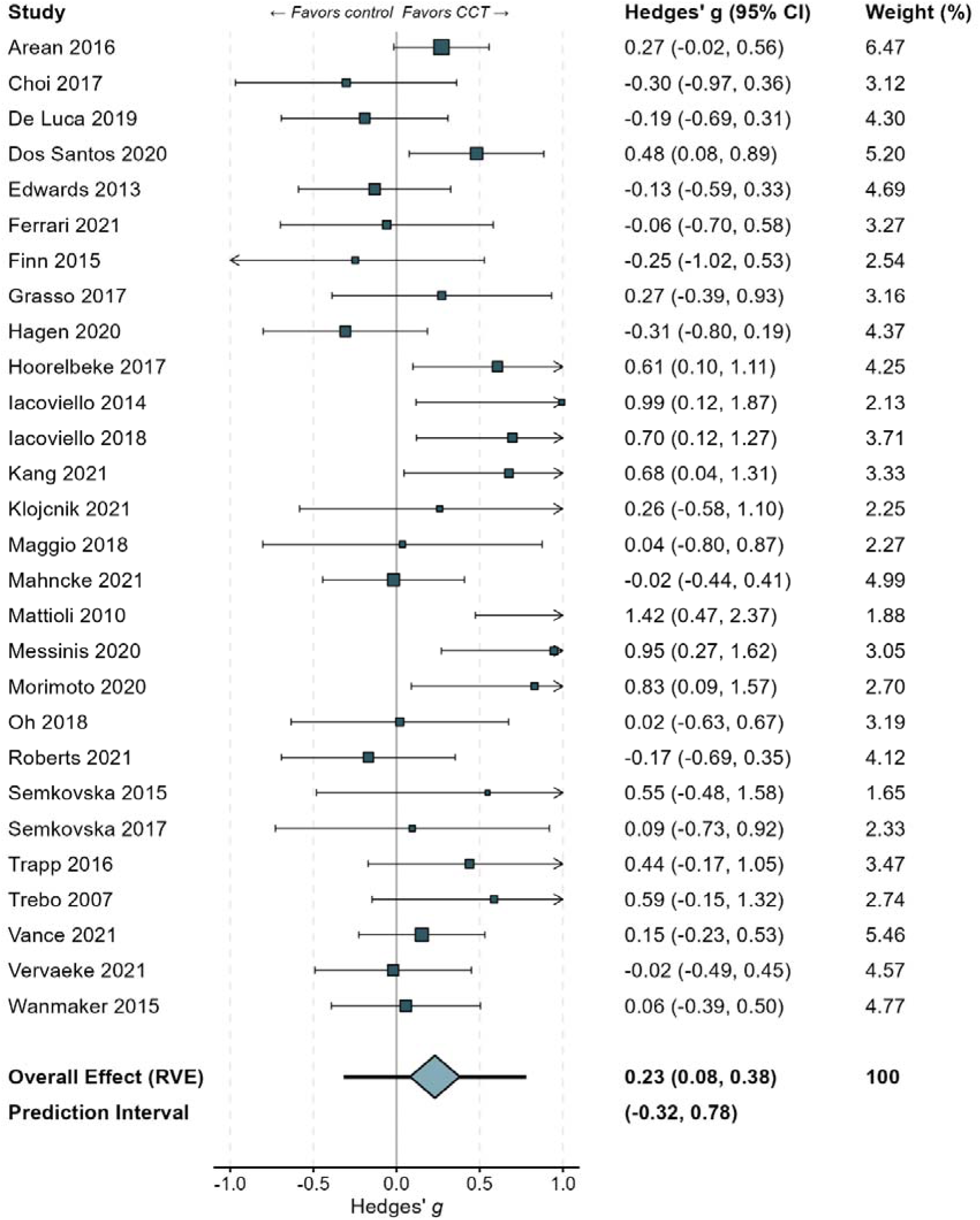
Meta-analysis of depressive symptoms.

### Secondary Outcomes: Specific Cognitive Domains

Results of meta-analyses of individual cognitive domains for which sufficient data were available (i.e., *k*>3) are provided in **Figure 4**. Small-to-moderate effect sizes were found for all the broad CHC-M domains apart from visual processing, which was estimated from 5 studies and was therefore imprecise. Small-to-moderate effect sizes were found for the narrow domains of abstract reasoning, learning, retrieval, low working memory, shifting, inhibition and processing speed. The pooled effect size for global cognition screening tools was small and not statistically significant (*k*=6; *g*=0.21; 95% CI −0.12 to 0.55; *P*=.16; τ^2^=0.041; *I*^2^=35%). Forest and funnel plots for analyses of global cognition and individual cognitive domains are provided in **eFigures 12-45** in the Supplement.

**Figure 4:**
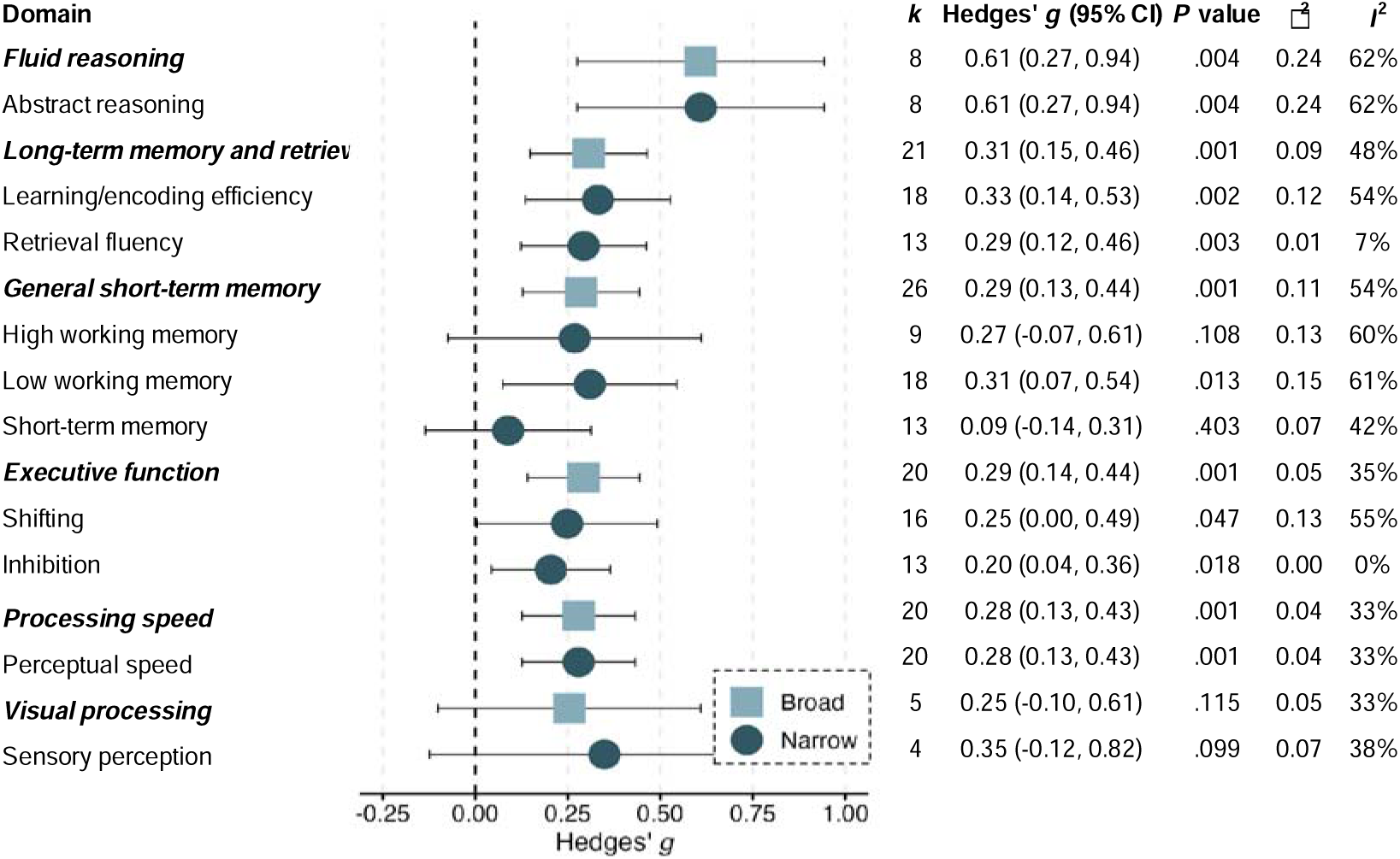
Meta-analyses individual cognitive domains.

## Discussion

To The best of our knowledge, this is the first systematic review with meta-analysis to examine the efficacy of CCT across populations with clinical depression, thereby extending the implications of results into other clinical populations with high prevalence of depressive symptoms, most notably older adults or those with neurodegenerative disorders. It overcomes the limitations of previous meta-analyses in the field by including only RCTs, strictly limiting interventions to CCT rather than other cognitive remediation strategies, and using multivariate methods that substantially improve our ability to detect and investigate heterogeneity. By using more sensitive methods and better controlling for study quality confounders, we were able to increase our certainty in the robustness of the results and examine potential effect moderators with high relevance to clinical practice.

We report robust findings that overall cognitive performance, depressive symptoms and nearly all specific cognitive domains appear to be responsive to CCT. The key findings were not confounded by risk of bias, type of control and population, and the impact of small-study bias on effect estimates was inconsequential. Particularly encouraging is the relatively large effect size for fluid (abstract) reasoning, which is key for everyday reasoning ability and suggested as a predictor of depressive symptoms in later life.^63^ Yet the effect size estimates for cognitive and depression endpoints are noticeably smaller and more heterogenous than those reported in previous meta-analyses in depression.^19–23^ This may be due to limiting the analyses to RCTs, using more robust statistical and the inclusion of more recent and rigorous trials, as evident by a tendency of non-registered and typically older studies to report larger effect sizes compared to preregistered trials. The effect estimates provided here are in line with those of previous meta-analyses of CCT in other clinical populations that used comparable synthesis methods.^14–16,27^ In addition, CCT was associated with small improvements in measures of subjective cognition and psychosocial functioning, but there was insufficient indication that these changes also relate to everyday functioning, social participation and self-care.

Taken together, the findings confirm the role of CCT in management of cognitive disorders in people with depression, but suggest that additional interventions may be needed in order to enhance its efficacy and functional impact. Cognitive remediation techniques have a very likely impact on people’s ability to carry objective cognitive improvement into everyday life,^10^ perhaps even independently from cognitive practice.^13^ Combination with physical exercise may augment cognitive effects, especially when provided simultaneously rather than in separate sessions, at least in older adults.^12^ The evidence regarding potential additive effects between CCT and neuromodulation, behavior change techniques,^64^ psychotherapy and antidepressants such as vortioxetine^65^ is more equivocal at this stage. Even less is known about how to combine interventions effectively given the numerous components and combination approaches (including order and dose) suggested in the literature. It is also not unlikely that some combinations will result in antagonistic effects, for instance by reducing adherence or effort.

Heterogeneity was substantial across studies, with about one-third of the prediction intervals falling around or below zero. CCT dose (i.e., total training hours) appears to play an important role here; nearly half of the studies provided a dose of less than 12 hours, which was associated with negligible overall cognitive and depression effect sizes. Sessions longer than 60 minutes all but eliminated the effect of symptoms, but paradoxically were associated with larger cognitive benefits. Consistent with previous synthesis work on CCT in older adults,^14^ we found that training regimens of >3 sessions per week are less efficacious than less frequent training, and that multidomain training is more efficacious for overall cognition than single-domain programs. Conversely to older adults, however, the mode of supervision was not associated with effect sizes, meaning that CCT by itself could be delivered successfully in in-person as well as home (self-administered or remotely supervised) settings.

While this work addresses some critical limitations of previous meta-analyses in the field and provides a rigorous platform for further designing and investigating the effectiveness of CCT across populations with depression, some limitations are noteworthy. First, since most studies focused on short-term cognitive and functional outcomes, the durability of the observed benefits as well as strategies to maintain them remain unclear. Second, although we were able to detect heterogeneity and investigate its potential sources to guide future intervention design, there are still not enough studies in the field to compare intervention components head-to-head (e.g., using network meta-analysis), meaning that the results of the subgroup analyses may be confounded by common design factors. Third, most studies did not report functional outcomes, leaving the pooled analyses of psychiatric symptoms, psychosocial functioning, daily functioning and subjective cognition underpowered and to be interpreted with caution. Similarly, only four studies^46–48^ specifically focused on people with partially and/or fully remitted depression, leaving subgroup analysis for this population also underpowered. Future studies should make efforts to include functional and patient-centered outcomes alongside clinical measures to investigate the true contribution of CCT to improve social and community participation across depression states.

### Conclusion

CCT could be efficacious for improving overall cognition, specific cognitive domains and mood with potential carry-over effects on indicators of psychosocial functioning in people with clinical depression, including older adults and those with other neurological disorders. Training dose, frequency and content appear to be stronger indicators of outcomes than the type of population or delivery mode. To progress the field, future studies may consider increasing their overall dose, compare CCT approaches and combinations with other common or potential interventions and investigate the long-term implications of CCT effects, with a particular focus on individual factors, goals and trajectories.

## Data Availability

Supplementary tables and figures are available from the link below.

https://figshare.com/s/3becd6c550107121b15f

## Notes

### Competing Interest Statement

The authors have declared no competing interest.

### Clinical Protocols

https://www.researchsquare.com/article/rs-66217/v1

### Funding Statement

This work has been supported by a CR Roper Fellowship from the University of Melbourne provided to AL.

### Summary of Updates

Updated search (October 2022), including 10 additional studies and updated analysis. The results and conclusions are consistent with those of the previous version of this manuscript.

