## Supplementary Information for "Computerized Cognitive Training in People with Depression: A Systematic Review and Meta-analysis of Randomized Clinical Trials"

**Supplementary Online Content**

**eTable 1.** Depression Measures Cut-offs

**eTable 2.** Search Strategy

**eTable 3.** Classification of Cognitive and Non-cognitive Outcomes

**eTable 4.** Classification of Baseline Depressive Symptoms Severity and Population Subgroups

**eTable 5.** List of Studies Excluded During Full Text Screening and Reasons for Exclusion

**eTable 6.** Risk of Bias Within Individual Studies

**eTable 7.** Sensitivity Analyses of Correlation Assumptions

**eFigure 1.** Funnel Plot of Overall Cognition

**eFigure 2.** Trim and Fill Funnel Plot of Overall Cognition

**eFigure 3.** Forest Plot of Depressive Symptoms

**eFigure 4.** Funnel Plot of Depressive Symptoms

**eFigure 5.** Trim and Fill Funnel Plot of Depressive Symptoms

**eFigure 6.** Forest Plot of Psychosocial Functioning

**eFigure 7.** Funnel Plot of Psychosocial Functioning

**eFigure 8.** Forest Plot of Psychiatric Symptoms

**eFigure 9.** Funnel Plot of Psychiatric Symptoms

**eFigure 10.** Forest Plot of Subjective Cognition

**eFigure 11.** Funnel Plot of Subjective Cognition

**eFigure 12.** Forest Plot of Global Cognition

**eFigure 13.** Funnel Plot of Global Cognition

**eFigure 14.** Forest Plot of Fluid Reasoning

**eFigure 15.** Funnel Plot of Fluid Reasoning

**eFigure 16.** Forest Plot of Abstract Reasoning

**eFigure 17.** Funnel Plot of Abstract Reasoning

**eFigure 18.** Forest Plot of Long-term Memory and Retrieval

**eFigure 19.** Funnel Plot of Long-term Memory and Retrieval

**eFigure 20.** Forest Plot of Learning/Encoding Efficiency

**eFigure 21.** Funnel Plot of Learning/Encoding Efficiency

**eFigure 22.** Forest Plot of Retrieval Fluency

**eFigure 23.** Funnel Plot of Retrieval Fluency

**eFigure 24.** Forest Plot of General Short-term Memory

**eFigure 25.** Funnel Plot of General Short-term Memory

**eFigure 26.** Forest Plot of High Working Memory

**eFigure 27.** Funnel Plot of High Working Memory

**eFigure 28.** Forest Plot of Low Working Memory

**eFigure 29.** Funnel Plot of Low Working Memory

**eFigure 30.** Forest Plot of Short-term Memory

**eFigure 31.** Funnel Plot of Short-term Memory

**eFigure 32.** Forest Plot of Executive Function

**eFigure 33.** Funnel Plot of Executive Function

**eFigure 34.** Forest Plot of Shifting

**eFigure 35.** Funnel Plot of Shifting

**eFigure 36.** Forest Plot of Inhibition

**eFigure 37.** Funnel Plot of Inhibition

**eFigure 38.** Forest Plot of Processing Speed

**eFigure 39.** Funnel Plot of Processing Speed

**eFigure 40.** Forest Plot of Perceptual Speed

**eFigure 41.** Funnel Plot of Perceptual Speed

**eFigure 42.** Forest Plot of Visual Processing

**eFigure 43.** Funnel Plot of Visual Processing

**eFigure 44.** Forest Plot of Sensory Perception

**eFigure 45.** Funnel Plot of Sensory Perception

This supplementary material has been provided by the authors to give readers additional information about their work.

**eTable 1.** Depression Measures Cut-offs

| **Depression Rating Scales** | **Rating method** | **Total score** | **Cut-off value for depressive disorder**^1^ |
| --- | --- | --- | --- |
| Beck Depression Inventory (BDI) | Self-rated | 63 | ≥10 |
| Beck Depression Inventory – II (BDI-II) | Self-rated | 63 | ≥14 |
| Beck Depression Inventory – Fast Screen (BDI-FS) | Self-rated | 21 | ≥4 |
| Center for Epidemiologic Studies Depression Scale (CES-D) | Self-rated | 60 | ≥16 |
| Chicago Multiscale Depression Inventory (CMDI) Mood Subscale | Self-rated | 70 | ≥23 |
| Depression Anxiety Stress Scales – 21 items (DASS-21) Depression Subscale | Self-rated | 21 | ≥5 |
| Geriatric Depression Scale – 15 items (GDS-15) | Self-rated | 15 | ≥5 |
| Geriatric Depression Scale – 30 items (GDS-30) | Self-rated | 30 | ≥10 |
| Hamilton Depression Rating Scale – 17 items (HDRS-17) | Clinician rated | 54 | ≥8 |
| Hamilton Depression Rating Scale – 24 items (HDRS-24) | Clinician rated | 76 | ≥10 |
| Hospital Anxiety and Depression Scale (HADS-depression) | Self-rated | 21 | ≥8 |
| Montgomery and Asberg Depression Rating Scale (MADRS) | Clinician rated | 60 | ≥7 |
| Patient Health Questionnaire 9 (PHQ-9) | Self-rated | 27 | ≥5 |

^1^The first cut-off value for major depressive disorder if a scale has multiple cut-off values. Higher scores represent more severe depressive symptoms.

**eTable 2.** Search Strategy

| # 1 | ((cognit* or attention or neurocognit* or neuropsycholog* or memory or reasoning or executive) adj2 (training* or rehabilitat* or remediat* or stimulat* or exercis* or retrain*)).mp. |
| --- | --- |
| # 2 | ((brain) adj2 (training* or rehabilitat* or remediat* or retrain*)).mp. |
| # 3 | (speed adj3 training).mp |
| # 4 | 1 or 2 or 3 |
| # 5 | Exp Depressive Disorder/ |
| # 6 | Depression/ |
| # 7 | Depress*.tw |
| # 8 | Depress*.kw |
| # 9 | 5 or 6 or 7 or 8 |
| # 10 | 4 and 9 |

**eTable 3.** Classification of Cognitive and Non-Cognitive Outcomes

| **Fluid reasoning** | |
| --- | --- |
| **Abstract reasoning** | |
| - Weigl’s Test (WEIGL) - Vienna Test System Tower of London-F (VTS TOL-F) - Wisconsin Card Sorting Test – Total Errors (WCSTte) - Wisconsin Card Sorting Test – Perseverative Errors (WCSTpe) - Wisconsin Card Sorting Test – Perseverative Responses (WCSTpr) - Delis-Kaplan Executive Function System (D-KEFS) 20 Questions - Delis-Kaplan Executive Function System (D-KEFS) Sorting - Delis-Kaplan Executive Function System (D-KEFS) Towers |  |
| **Visual processing** | |
| **Sensory perception** | **Visualization** |
| - Vienna Test System WAF-Divided Attention (VTS WAF-G) – Misses - Vienna Test System WAF-Selective Attention Visual (VTS WAF-S Visual) – False Alarms - Vienna Test System WAF-Selective Attention Auditory (VTS WAF-S Auditory) – False Alarms - Vienna Test System WAF-Selective Attention Crossmodal (VTS WAF-S Crossmodal) – False Alarms - Wechsler Memory Scale-Revised (WMS-R) Visual Reproduction I - Rey-Osterrieth Complex Figure Test (RCFT) copy basic components - Test for Attentional Performance Battery (TAP) Selective Attention subscale | - Wechsler Adult Intelligence Scale - IV (WAIS-IV) Block Design |
| **Processing speed** | |
| **Perceptual speed** | |
| - Symbol Digit Modalities Test (SDMT) - Rao’s Brief Repeatable Battery of Neuropsychological Tests Symbol Digit Modalities Test (BRB-N SDMT) - Stroop Color Score/Stroop Color Naming Test - Stroop Word Score - Symbol Coding Task - Vienna Test System Trail Making Test-A (VTS TMT-A) - Useful Field of View (UFOV) - Connors Continuous Performance Test Version 3 (CPT-3) – Block Change Hit RT - Vienna Test System WAF-Alertness (VTS WAF-A) – Reaction Time - Vienna Test System WAF-Divided Attention (VTS WAF-G) – Reaction Time - Vienna Test System WAF-Selective Attention Visual (VTS WAF-S Visual) – Reaction Time - Test for Attentional Performance Battery (TAP) Alertness subscale | - Vienna Test System WAF-Selective Attention Auditory (VTS WAF-S Auditory) – Reaction Time - Vienna Test System WAF-Selective Attention Crossmodal (VTS WAF-S Crossmodal) – Reaction Time - Memory Diagnostic System Attention Quotient (MDS AQ) – Omission Error - Memory Diagnostic System Attention Quotient (MDS AQ) – Commission Error - Memory Diagnostic System Attention Quotient (MDS AQ) – Reaction Time - Memory Diagnostic System Attention Quotient (MDS AQ) – Reaction Time SD - Wechsler Adult Intelligence Scale-III (WAIS-III) Digit Symbol Substitution - Attention performance - Color-words reading - d2 – Speed - Naming color line |
| **Long-term memory and retrieval** | |
| **Learning/encoding efficiency** | |
| - Hopkins Verbal Learning Test (HVLT) - Hopkins Verbal Learning Test (HVLT) - Delayed - Buschke Selective Reminding Test (BSRT) Total List Recall - Buschke Selective Reminding Test (BSRT) Delayed Recall - Buschke Selective Reminding Test (BSRT) Long-Term Retrieval - Wechsler Memory Scale-IV (WMS-IV) Verbal Paired Associates I - Wechsler Memory Scale-IV (WMS-IV) Verbal Paired Associates II - Rao’s Brief Repeatable Battery of Neuropsychological Tests Spatial Recall Test-Delayed (BRB-N SPART-D) - Rao’s Brief Repeatable Battery of Neuropsychological Tests Selective Reminding Test-Delayed (BRB-N SRT-D) - Rao’s Brief Repeatable Battery of Neuropsychological Tests Selective Reminding Test-Long-Term Storage (BRB-N SRT-LTS) - California Verbal Learning Test (CVLT) Learning Sum - California Verbal Learning Test (CVLT) Delayed Recall - California Verbal Learning Test (CVLT) Immediate Recall - Vienna Test System Figural Memory Test (VTS FMT) Delayed Recall - Vienna Test System Figural Memory Test (VTS FMT) Immediate Recall - Vienna Test System Figural Memory Test (VTS FMT) Learning Sum - Rao’s Brief Repeatable Battery of Neuropsychological Tests Selective Reminding Test Consistent Long-Term Retrieval (BRB-N SRT/CLTR) - Seoul Verbal Learning Test (SVLT) Delayed Recall - Seoul Verbal Learning Test (SVLT) Immediate Recall - Seoul Verbal Learning Test (SVLT) Recognition | - Rao’s Brief Repeatable Battery of Neuropsychological Tests Selective Reminding Test Delayed Retrieval (BRB-N SRT/DR) - Rao’s Brief Repeatable Battery of Neuropsychological Tests 10/36 Spatial Recall Long-Term Retrieval (BRB-N 10/36 SRT/LTR) - Rao’s Brief Repeatable Battery of Neuropsychological Tests 10/36 Spatial Recall Delayed Recall (BRB-N 10/36 SRT/DR) - Rao’s Brief Repeatable Battery of Neuropsychological Tests 10/36 Spatial Recall Immediate Recall (BRB-N 10/36 SRT/IR) - Selective Reminding Test Long-Term Storage (SRT LTS) - Selective Reminding Test Delayed Recall (SRT DR) - Memory Diagnostic System Memory Quotient (MDS MQ) – Auditory-Verbal Immediate - Memory Diagnostic System Memory Quotient (MDS MQ) – Auditory-Verbal Delayed - Memory Diagnostic System Memory Quotient (MDS MQ) – Visual-Spatial Delayed - Wechsler Memory Scale-III (WMS-III) Logical Memory-I - Wechsler Memory Scale-III (WMS-III) Logical Memory-II - Rey-Osterrieth Complex Figure (ROCF) Immediate Recall - Rey-Osterrieth Complex Figure (ROCF) Delayed Recall - Rey Auditory Verbal Learning Test - Wechsler Memory Scale-Revised (WMS-R) Visual Reproduction II - Wechsler Memory Scale-Revised (WMS-R) Logical Memory II - IST-verbal memory - Grober-Buschke Delayed Recall - Grober-Buschke Free Recall - Greek Verbal Learning Test (GVLT) |
| **Retrieval fluency** | |
| - Animal Naming Test - Controlled Oral Word Association Test (COWA) Phonemic Cue - Controlled Oral Word Association Test (COWA) Semantic Cue - Columbia University Autobiographical Memory Interview-Short Form (AMI-SF) - Goldberg Remote Memory Questionnaire (GRMQ) - Addenbrooke’s Cognitive Examination-Revised (ACE-R) Fluency - Addenbrooke’s Cognitive Examination-Revised (ACE-R) Language - Rao’s Brief Repeatable Battery of Neuropsychological Tests (BRB-N) Word List Generation - Korean Boston Naming Test (K-BNT) | - Greek Verbal Fluency Test Phonemic Fluency (Greek VFT Phonemic) - Greek Verbal Fluency Test Semantic Fluency (Greek VFT Semantic) - Controlled Oral Word Association Test (COWA) FAS - California Verbal Learning Test-II (CVLT-II) Semantic Clustering - California Verbal Learning Test-II (CVLT-II) Long Delay Recall - Delis-Kaplan Executive Function System (D-KEFS) Verbal Category Fluency - Delis-Kaplan Executive Function System (D-KEFS) Category Switching Fluency - Delis-Kaplan Executive Function System (D-KEFS) Letter Fluency |
| **General short-term memory** | |
| **Short-term memory** | **High working memory** |
| - Spatial Span - Addenbrooke’s Cognitive Examination-Revised (ACE-R) Memory - Wechsler Memory Scale-IV (WMS-IV) Symbol Span - Addenbrooke’s Cognitive Examination-Revised (ACE-R) Memory - Memory Diagnostic System Memory Quotient (MDS MQ) – Visual Spatial Immediate - Wechsler Adult Intelligence Scale-III (WAIS-III) Digit Span Forwards - Wechsler Memory Scale-Revised (WMS-R) Digit Span Forwards - Wechsler Memory Scale-Revised (WMS-R) Spatial Span Forwards - Wechsler Memory Scale-Revised (WMS-R) Logical Memory I - Digit Span Forwards Score - Benton Visual Retention Test- Repeating numbers | - Rao’s Brief Repeatable Battery of Neuropsychological Tests Paced Auditory Serial Addition Test 2 Seconds (BRB-N PASAT 2) - Rao’s Brief Repeatable Battery of Neuropsychological Tests Paced Auditory Serial Addition Test 3 Seconds (BRB-N PASAT 3) - Paced Auditory Serial Attention Test - Delis-Kaplan Executive Function System (D-KEFS) Number Sequencing - Letter Number Sequencing Test - Auditory-Verbal Working Memory (Auditory-Verbal WM) - Reading Span – Partial-Credit Unit (PCU) Score - Reading Span – Sentence Errors |
| **Low working memory** | |
| - Addenbrooke’s Cognitive Examination-Revised (ACE-R) Attention and Orientation - Addenbrooke’s Cognitive Examination-Revised (ACE-R) Visual Spatial - Rao’s Brief Repeatable Battery of Neuropsychological Tests Spatial Recall Test (BRB-N SPART) - Connors Continuous Performance Test Version 3 (CPT-3) – Commissions subscale - Attention and Working Memory Composite (Digit-Span Forward, Digit-Span Backward, Letter-Number Sequencing subtests of Wechsler Adult Intelligence Scale-III) - Zahlen-Symbol-Test (ZST) Digit Symbol Coding – Number Correct - Digit Span Backwards Score | - Brief Visuospatial Memory Test-Revised (BVMT-R) Total Recall - Wechsler Adult Intelligence Scale-IV (WAIS-IV) Digit Span Backwards - Visual-Spatial Working Memory (Visual-Spatial WM) - Wechsler Adult Intelligence Scale-III (WAIS-III) Digit Span Backwards - Degraded Continuous Performance Test (Degraded CPT) – Commissions - Degraded Continuous Performance Test (Degraded CPT) – Omissions - Wechsler Memory Scale-Revised (WMS-R) Digit Span Backwards - Wechsler Memory Scale-Revised (WMS-R) Spatial Span Backwards |
| **Executive functions** | |
| **Inhibition** | |
| - Stroop Color-Word Interference - Delis-Kaplan Executive Function System (D-KEFS) Color-Word Interference – Condition 3 (CWIT 3) - Delis-Kaplan Executive Function System (D-KEFS) Color-Word Interference – Condition 4 (CWIT) - Delis-Kaplan Executive Function System (D-KEFS) Color-Word Interference | - Vienna Test System (VTS) Inhibition – False Alarms - Vienna Test System (VTS) Inhibition – Reaction Time - d2 – Accuracy - d2 – Deviation - d2 – Percentage of mistakes - Interference trials |
| **Updating** | |
| - Continuous Performance Test (CPT) Identical Pairs Version - Vienna Test System (VTS) N-Back-Verbal – Misses |  |
| **Shifting** | |
| - Task Switch - Vienna Test System Trail Making Test-B (VTS TMT-B) - Delis-Kaplan Executive Function System (D-KEFS) Number-Letter Switching - Rao’s Brief Repeatable Battery of Neuropsychological Tests Test of Everyday Attention – Auditory Stimulus (BRB-N TEAam) - Rao’s Brief Repeatable Battery of Neuropsychological Tests Test of Everyday Attention – Visual Stimulus (BRB-N TEAvm) - Rao’s Brief Repeatable Battery of Neuropsychological Tests Test of Everyday Attention – Total Errors (BRB-N TEAte) | - Rao’s Brief Repeatable Battery of Neuropsychological Tests Test of Everyday Attention – Total Omitted Stimuli (BRB-N TEAto) - Delis-Kaplan Executive Function System (D-KEFS) Design Fluency Switching - d2 Selective Attention Test – % errors - d2 Selective Attention Test – Total Correct - Internal Shift Task (IST) – Emotion Shift Costs - Internal Shift Task (IST) – Gender Shift Costs - Internal Shift Task (IST) – Global Shift Costs - Test for Attentional Performance Battery (TAP) Flexibility subscale - Test for Attentional Performance Battery (TAP) Divided Attention subscale |
| **Global cognition** | **Daily functioning** |
| - Mini Mental State Examination (MMSE) - Modified Mini Mental State Examination (mMMS) - Frontal Assessment Battery (FAB) - Addenbrooke’s Cognitive Examination-Revised (ACE-R) - Global Cognitive Score | - Advanced Finances Task - Longitudinal Interval Follow-up Evaluation Range of Impaired Functioning Tool (LIFE-RIFT) - Barthel Index (BI) - Expanded Disability Scale Score (EDSS) - Rivermead Mobility Index (RMI) - Modified Fatigue Impact Scale (MFIS) Physical subscale - Timed Instrumental Activities of Daily Living (TIADL) - Mayo-Portland Adaptability Index (MPAI) - Modified Lawton and Brody Activities of Daily Living (ADL) questionnaire BADL - Modified Lawton and Brody Activities of Daily Living (ADL) questionnaire IADL |
| **Depressive symptoms** | **Psychiatric symptoms** |
| - Beck Depression Inventory (BDI) - Beck Depression Inventory-II (BDI-II) - Beck Depression Inventory-Fast Screen (BDI-FS) - Center for Epidemiologic Studies Depression Scale (CES-D) - Chicago Multiscale Depression Inventory (CMDI) Mood - The Depression, Anxiety and Stress Scale – 21 items (DASS-21) Depression subscale - Geriatric Depression Scale – 30 items (GDS-30) - Hamilton Depression Rating Scale – 17 items (HDRS-17) - Hamilton Depression Rating Scale – 24 items (HDRS-24) - Montgomery and Ashberg Depression Rating Scale (MADRS) - Patient Health Questionnaire-9 (PHQ-9) - Positive and Negative Affect Schedule-Negative Affect (PANAS-N) - Positive and Negative Affect Schedule-Positive Affect (PANAS-P) | - Hamilton Rating Scale Anxiety (HRS-A) - The Depression, Anxiety and Stress Scale – 21 items (DASS-21) Anxiety subscale - The Depression, Anxiety and Stress Scale – 21 items (DASS-21) Stress subscale - Ruminative Response Scale (RRS) - Ruminative Response Scale (RRS) Brooding subscale - Ruminative Response Scale (RRS) Reflection subscale - Self-referential information processing (SRIP) task - State-Trait Anxiety Inventory (STAI) - State-Trait Anxiety Inventory State Anxiety subscale (STAI-S) - Cognitive Emotion Regulation Questionnaire (CERQ) Adaptive ER subscales - Cognitive Emotion Regulation Questionnaire (CERQ) Maladaptive ER subscales - Momentary Ruminative Self-Focus Inventory (MRSI) - Response Style Questionnaire (RSQ) Distraction subscale - Response Style Questionnaire (RSQ) Rumination subscale - Apathy Evaluation Scale (AES) - Penn State Worry Questionnaire (PSWQ) - Post-Traumatic Stress Disorder Checklist - Civilian version (PCL-C) - The Frontal Symptoms Behavioral Scale (FrSBe) |
| **Psychosocial functioning** | |
| - Social Skills Performance Assessment (SSPA) - Global Assessment of Functioning (GAF) - Medical Outcomes Study Short Form – 12 items (SF-12) - Medical Outcomes Study Short Form – 36 items (SF-36) Mental Health subscales - Medical Outcomes Study Short Form – 36 items (SF-36) Physical Health subscales - The Medical Outcomes Study- HIV (MOS-HIV) - Mini- International Classification of Functioning, Disability and Health Self Assessment (Mini-ICF Self) - Mini- International Classification of Functioning, Disability and Health External assessment (Mini-ICF External) - Quality of Life-Alzheimer Disease (QoL-AD) - EuroQOL 5D Visual Analogue Scale (EQ5VAS) - Modified Fatigue Impact Scale (MFIS) Psychosocial subscale - Modified Fatigue Impact Scale (MFIS) Total - Quality of Life in Depression Scale (QLDS) | - Specific Level of Function Scale (SLOF) - Multiple Sclerosis Quality of Life (MSQoL) - World Health Organization Disability Assessment Schedule-II (WHODAS-II) - Multifactorial Memory Questionnaire Contentment subscale (MMQ-C) - Psychological status – negative - Psychological status – positive - SKI-PC - Functional Assessment of Cancer Therapy-Anemia (FACT-An) - Functional Assessment of Cancer Therapy-Cognitive Function (FACT-Cog) impact on quality of life subscale - Functional Assessment of Cancer Therapy-General (FACT-G) emotional well-being subscale - Functional Assessment of Cancer Therapy-General (FACT-G) functional well-being subscale - Functional Assessment of Cancer Therapy-General (FACT-G) physical well-being subscale - Functional Assessment of Cancer Therapy-General (FACT-G) social well-being subscale - Functional Assessment of Cancer Therapy-General (FACT-G) total |
| **Subjective cognition** | |
| - Cognitive Failures Questionnaire (CFQ) - Squire Subjective Memory Questionnaire (SSMQ) - Cognitive Self-Report Questionnaire - Behavior Rating Inventory of Executive Function-Adult version (BRIEF-A) Inhibit subscale - Behavior Rating Inventory of Executive Function-Adult version (BRIEF-A) Self-Monitor subscale - Behavior Rating Inventory of Executive Function-Adult version (BRIEF-A) Plan/Organize subscale - Behavior Rating Inventory of Executive Function-Adult version (BRIEF-A) Shift subscale - Behavior Rating Inventory of Executive Function-Adult version (BRIEF-A) Initiate subscale - Behavior Rating Inventory of Executive Function-Adult version (BRIEF-A) Task Monitor subscale - Behavior Rating Inventory of Executive Function-Adult version (BRIEF-A) Emotional Control subscale - Modified Fatigue Impact Scale (MFIS) Cognitive subscale | - Behavior Rating Inventory of Executive Function-Adult version Working Memory subscale (BRIEF-A WM) - Behavior Rating Inventory of Executive Function-Adult version Organization of Materials subscale (BRIEF-A OoM) - Behavior Rating Inventory of Executive Function-Adult version Behavioral Regulation Index (BRIEF-A BRI) - Behavior Rating Inventory of Executive Function-Adult version Metacognitive Index (BRIEF-A MI) - Behavior Rating Inventory of Executive Function-Adult version Global Executive Composite (BRIEF-A GEC) - Multifactorial Memory Questionnaire Ability subscale (MMQ-A) - Multifactorial Memory Questionnaire Strategy subscale (MMQ-S) - Functional Assessment of Cancer Therapy-Cognitive Function (FACT-Cog) comments from others subscale - Functional Assessment of Cancer Therapy-Cognitive Function (FACT-Cog) perceived cognitive abilities subscale - Functional Assessment of Cancer Therapy-Cognitive Function (FACT-Cog) perceived cognitive impairments subscale |

**eTable 4.** Classification of Baseline Depressive Symptoms Severity and Population Subgroups

| **Study** | **Comparison** | **Depression measure** | **Baseline depressive symptoms severity *M^a^*** | **Subgroup depressive symptoms severity classification** | **Subgroup population classification** |
| --- | --- | --- | --- | --- | --- |
| Amato 2014 | ST vs n-ST | MADRS | 8.51 | Mild | Multiple sclerosis |
| Arean 2016 | EVO vs iPST | PHQ-9 | 13.63 | Moderate-severe | MDD |
| Arean 2016 | EVO vs HT | PHQ-9 | 13.70 | Moderate-severe | MDD |
| Bowie 2013 | CR vs WL | MADRS | 24.00 | Moderate-severe | MDD |
| Choi 2017 | M-ECT vs AC | HDRS-24 | 25.69 | Moderate-severe | MDD |
| Choi 2017 | M-ECT vs TAU | HDRS-24 | 25.05 | Moderate-severe | MDD |
| De Luca 2019 | EG vs CG | GDS-30 | 19.00 | Mild | Parkinson’s disease |
| Dos Santos 2020a | CR vs Exercises at Home | CES-D | 21.04 | Moderate-severe |  |
| Dos Santos 2020b | CR vs Phone Call | CES-D | 22.05 | Moderate-severe |  |
| Edwards 2013 | SOPT vs CG | CES-D | 17.18 | Mild | Parkinson’s disease |
| Ferrari 2021 | Active-training vs Sham-training | BDI-II | 23.22 | Moderate-severe | MDD |
| Finn 2015 | Training vs Control | DASS-21 Depression | 6.17 | Mild | SCD/MCI |
| Grasso 2017 | CMD vs MD | MADRS | 20.67 | Moderate-severe | Multiple sclerosis |
| Hagen 2020 | CCT vs GMT | BDI | 16.30 | Moderate-severe | MDD |
| Hoorelbeke 2017 | CCT vs Control | BDI-II | 8.02 | Mild | Remitted MDD |
| Iacoviello 2014 | EMFT vs CT | HDRS-17 | 20.72 | Moderate-severe | MDD |
| Iacoviello 2018 | EMFT vs CT | HDRS-17  BDI-II | 19.35  30.12 | Moderate-severe | MDD |
| Kang 2021 | VR Cognitive Training vs Usual Care | GDS-30 | 13.76 | Mild | SCD/MCI |
| Klojčnik 2021 | CCRT vs Control | BDI-II | 23.70 | Moderate-severe | MDD |
| Listunova 2020 | IT vs CG | HDRS-24  BDI-II | 10.46  18.46 | Mild | Remitted MDD |
| Listunova 2020 | GT vs CG | HDRS-24  BDI-II | 10.32  18.48 | Mild | Remitted MDD |
| Maggio 2018 | Experimental vs Control | GDS-30 | 11.00 | Mild | Parkinson’s disease |
| Mahnke 2021 | BrainHQ vs Computer games | BDI-II | 18.7 | Mild |  |
| Mattioli 2010 | SG vs CG | MADRS | 10.83 | Mild | Multiple sclerosis |
| Messinis 2017 | RehaCom vs CG | BDI-FS | 4.42 | Mild | Multiple sclerosis |
| Messinis 2020 | RehaCom vs CG | BDI-FS | 6.36 | Mild | Multiple sclerosis |
| Morimoto 2020 | nCCR vs AC | MADRS | 25.54 | Moderate-severe | MDD |
| Oh 2018 | SMART vs WL | CES-D | 18.09 | Mild | SCD/MCI |
| Oh 2018 | Fit Brains vs WL | CES-D | 16.20 | Mild | SCD/MCI |
| Roberts 2021 | NEUA vs NEU | PHQ-9 | 5.90 | Mild |  |
| Semkovska 2015 | NCRT vs Games | HDRS-17  BDI-II | 19.20  33.05 | Moderate-severe | MDD |
| Semkovska 2017 | NCRT vs Games | HDRS-17  BDI-II | 4.25  6.40 | Mild | Remitted MDD |
| Solari 2004 | SG vs CG | CMDI-Mood | 28.66 | Moderate-severe | Multiple sclerosis |
| Trapp 2016 | EG vs CG | BDI-II  HDRS-17 | 27.7  12.04 | Moderate-severe | MDD |
| Trebo 2007 | Training Group vs CG | BDI | 23.85 | Moderate-severe | MDD |
| Vance 2021 | BrainHQ vs Passive Control | CES-D | 18.11 | Mild |  |
| Vervake 2021 | aPASAT vs speed-of-response training task | BDI-II | 9.75 | Mild | Remitted unipolar and bipolar depression |
| Vilou 2020 | Intervention Group vs Control Group | BDI-FS | 4.14 | Mild | Multiple sclerosis |
| Wanmaker 2015 | EG vs PG | BDI-II | 21.46 | Mild | MDD |

^a^ Weighted mean of baseline depression severity for CCT and control groups

Severity was classified according to validated cut-off scores for depression rating scales (see eTable 1 in the Supplement). Studies with remitted depression populations with mean baseline depressive symptoms scores below the cut-offs for mild depression were grouped with the mild depression subgroup for meta-regression of baseline depressive symptom severity due to only two studies (that focused exclusively on people with fully remitted depression) having scores below cut-offs. Only one study included a mix of people with partially and/or fully remitted depression, so this study was included in the subgroup ‘remitted MDD’. Four studies (Dos Santos 2020, Roberts 2021, Mahnke 2021 and Vance 2022) were not included in population subgroups as they did not specifically recruit participants with a formal current or past diagnosis of major depressive disorder and did not fit into any of the other population subgroups, and consequently were not included in the meta-regression of population.

**eTable 5.** List of Studies Excluded During Full Text Screening and Reasons for Exclusion

| **Study** | **Titles of Ineligible studies** | **Exclusion reason** |
| --- | --- | --- |
| Aasvik 2003 | Effectiveness of Working Memory Training among Subjects Currently on Sick Leave Due to Complex Symptoms | Data also elsewhere |
| Aben 2014 | Long-lasting effects of a new Memory Self-efficacy training for stroke patients: A randomized controlled trial | Not adults with dep. |
| AkhlaghiJami 2020 | Efficacy of cognitive rehabilitation therapy on stress and anxiety of the high school second level female students | Not adults with dep. |
| Akin-Sari 2022 | Cognitive training using a mobile app as a coping tool against COVID-19 distress: A crossover randomized controlled trial | Not adults with dep. |
| Akodu 2021 | Effects of core stabilization exercise and cognitive behavioural therapy in the management of patients with non-specific chronic low back pain | Not adults with dep. |
| AlQasem 2022 | Working Memory and Transcranial-Alternating Current Stimulation-State of the Art: Findings, Missing, and Challenges | Not RCT |
| Alaimo 2021 | Cognitive Tele-Enhancement in Healthy Older Adults and Subjects With Subjective Memory Complaints: A Review | Not RCT |
| Alexopoulos 2021 | Mechanisms and treatment of late-life depression | Not RCT |
| Alizadeh 2022 | The effect of functional training on level of brain-derived neurotrophic factor and functional performance in women with obesity | Not CCT |
| Alketbi 2021 | The added value of cognition-targeted exercise versus symptom-targeted exercise for multiple sclerosis fatigue: A randomized controlled pilot trial | Not CCT |
| Allan 2018 | Cognitive Remediation Therapy for Psychotic Major Depressive Disorder | Not RCT |
| Allen 2017 | Effects of computer cognitive training on depression in cognitively impaired seniors | Not RCT |
| Alon 2022 | A randomized controlled trial of supervised remotely delivered attention bias modification for posttraumatic stress disorder | Not CCT |
| Alvarez 2008 | Computer program in the treatment for major depression and cognitive impairment in university students | Data not available |
| Amini 2022 | The effectiveness of cognitive-motor training on reconstructing cognitive health components in older male adults, recovered from the COVID-19 | Not RCT |
| Anguera 2016 | Conducting a fully mobile and randomised clinical trial for depression: access, engagement, and expense | Ineligible outcomes |
| Ansari 2015 | The therapeutic potential of working memory training for treating mental disorders | Not RCT |
| Araujo-Pedrosa 2022 | The usefulness of e-mental health tools to promote maternal mental health in the context of the COVID-19 pandemic | Not CCT |
| ArditteHall 2018 | Positive memory enhancement training for individuals with major depressive disorder | Not CCT |
| Arean 2015 | Targeted treatments for late life depression: From stepped psychotherapy to computer games | Data also elsewhere |
| ArianDarestani 2020 | The therapeutic effect of treatment with RehaCom software on verbal performance in patients with multiple sclerosis | Ineligible outcomes |
| Arjmandnia 2015 | Efficacy of memory specificity training (MEST) on underlying mechanisms of overgeneral autobiographical memory (OGM) in people with major depression and childhood traumatic experience. | Not CCT |
| Assonov 2021 | Two-step resilience-oriented intervention for veterans with traumatic brain injury: A pilot randomized controlled trial | Not CCT |
| Au 2021 | Brief report: A randomized controlled trial of a compensatory cognitive training to improve prospective memory performance in people with schizophrenia or depression | Not CCT |
| Azimi 2019 | Effectiveness of cognitive behavioral therapy for insomnia (traditional and Internet-based) on everyday memory of people with insomnia and comorbid depression | Not CCT |
| Bae 2020 | The effect of a multicomponent dual-task exercise on cortical thickness in older adults with cognitive decline: A randomized controlled trial | Not adults with dep. |
| Bai 2021 | A narrative review of risk factors and interventions for cancer-related cognitive impairment | Not adults with dep. |
| Bail 2020 | Cancer-Related Symptoms and Cognitive Intervention Adherence Among Breast Cancer Survivors: A Mixed-Methods Study | Not adults with dep. |
| Ballantyne 2021 | Preliminary Support for a Cognitive Remediation Intervention for Women During the Menopausal Transition: A Pilot Study | Not RCT |
| Ballantyne 2021 | Cognitive remediation for women during the menopausal transition: a pilot study | Not RCT |
| Ballesteros 2015 | "Brain training with non-action video games enhances aspects of cognition in older adults: A randomized controlled trial": Corrigendum | Not adults with dep. |
| Banerjee 2021 | Digital gaming interventions: a novel paradigm in mental health? Perspectives from India | Not RCT |
| Barkus 2020 | Effects of working memory training on emotion regulation: Transdiagnostic review | Not RCT |
| Barth 2019 | Shifting Instead of Drifting - Improving Attentional Performance by Means of the Attention Training Technique | Not adults with dep. |
| Barry 2021 | The current state of memory Specificity Training (MeST) for emotional disorders | Not RCT |
| Bartlett 2020 | Multidisciplinary rehabilitation reduces hypothalamic grey matter volume loss in individuals with preclinical Huntington's disease: A nine-month pilot study | Not adults with dep. |
| Baune 2017 | Does training of cognitive functions help affective remediation? | Not RCT |
| Beddig 2020 | Mindfulness-based focused attention training versus progressive muscle relaxation in remitted depressed patients: Effects on salivary cortisol and associations with subjective improvements in daily life | Not CCT |
| Behrouian 2020 | The Effect of Emotion Regulation Training on Stress, Anxiety, and Depression in Family Caregivers of Patients with Schizophrenia: A Randomized Controlled Trial | Not CCT |
| Bergamaschi 2021 | ReBrain: A multidisciplinary cognitive rehabilitation protocol for people with Multiple Sclerosis, clinical practice and effectiveness | Not CCT |
| Bernhardt 2021 | Subjective cognitive and neurocognitive functions over the course of CBT | Not RCT |
| Besnier 2021 | Investigation of the Effects of Home-Based Exercise and Cognitive Training on Cognitive and Physical Functions in Cardiac Patients: The COVEPICARDIO Study Protocol of a Randomized Clinical Trial | Not RCT |
| Bettis 2017 | Comparison of two approaches to prevention of mental health problems in college students: Enhancing coping and executive function skills | Not RCT |
| Binarelli 2021 | Multimodal Web-Based Intervention for Cancer-Related Cognitive Impairment in Breast Cancer Patients: Cog-Stim Feasibility Study Protocol | Not RCT |
| Bischoff 2014 | Acceptance of smartphone-aided rehabilitation aftercare in depressive patients | Not CCT |
| Blackwell 2015 | Positive Imagery-Based Cognitive Bias Modification as a Web-Based Treatment Tool for Depressed Adults: A Randomized Controlled Trial | Not CCT |
| Blair 2021 | Does cognitive training improve attention/working memory in persons with MS? A pilot study using the Cogmed Working Memory Training program | Not adults with dep. |
| Blanca 2020 | Face to face vs. online cognitive stimulation for people with cognitive impairment. A controlled trial | Inappropriate control |
| Bockting 2022 | Augmenting neurocognitive remediation therapy to Preventive Cognitive Therapy for partially remitted depressed patients: protocol of a pragmatic multicentre randomised controlled trial | Not RCT |
| Bomyea 2011 | The Effect of an Executive Functioning Training Program on Working Memory Capacity and Intrusive Thoughts | Not adults with dep. |
| Bowie 2013 | Cognitive remediation therapy for mood disorders: rationale, early evidence, and future directions | Not RCT |
| Bowie 2016 | Cognitive remediation for major depressive disorder | Not RCT |
| Bowie 2017 | Action-based cognitive remediation for individuals with serious mental illnesses: Effects of real-world simulations and goal setting on functional and vocational outcomes | Not RCT |
| Bowie 2017 | Action-based cognitive remediation for individuals with serious mental illnesses: Effects of real-world simulations and goal setting on functional and vocational outcomes | Not RCT |
| Bowie 2022 | Cognitive remediation | Not RCT |
| Boyd 2022 | A Pilot Study Assessing the Effects of Goal Management Training on Cognitive Functions among Individuals with Major Depressive Disorder and the Effect of Post-Traumatic Symptoms on Response to Intervention | Not CCT |
| Brown 2021 | Protocol of a 12-month multifactorial eHealth programme targeting balance, dual-tasking and mood to prevent falls in older people: The StandingTall + randomised controlled trial | Not CCT |
| Browning 2011 | Using an experimental medicine model to explore combination effects of pharmacological and cognitive interventions for depression and anxiety | Not CCT |
| Brum 2009 | Cognitive training in older adults with Mild Cognitive Impairment: Impact on cognitive and functional performance | Not CCT |
| Buccellato 2020 | A randomized feasibility trial of a novel, integrative, and intensive virtual rehabilitation program for service members post-acquired brain injury | Not adults with dep. |
| Buettner 2011 | Cognitive stimulation for apathy in probable early-stage Alzheimer's | Not CCT |
| Buitenweg 2019 | Does cognitive flexibility training enhance subjective mental functioning in healthy older adults? | Not adults with dep. |
| Bulut 2022 | The reliability, validity, and responsiveness of Cognitive Exercise Therapy Approach: Biopsychosocial Questionnaire for patients with psoriatic arthritis | Not CCT |
| Bures 2016 | The effect of cognitive training on the subjective perception of well-being in older adults | Not adults with dep. |
| Bursky 2022 | Mindfulness-Enhanced Computerized Cognitive Training for Depression: An Integrative Review and Proposed Model Targeting the Cognitive Control and Default-Mode Networks | Not RCT |
| Burt 2020 | The Effects of Music-Contingent Gait Training on Cognition and Mood in Parkinson Disease: A Feasibility Study | Not adults with dep. |
| Burton 2020 | Combining tDCS and Cognitive Training for People With Severe Mental illness: Preliminary Findings | Not adults with dep. |
| Butler 2020 | Trauma, treatment and Tetris: video gaming increases hippocampal volume in male patients with combat-related posttraumatic stress disorder | Not adults with dep. |
| Cabral 2021 | Harnessing Neuroplasticity to Promote Brain Health in Aging Adults: Protocol for the MOVE-Cog Intervention Study | Not CCT |
| Calatayud 2021 | Analysis of the effect of cognitive stimulation program in older adults with normal cognition: randomized clinical trial | Not CCT |
| Calkins 2013 | Testing the boundaries of computerized cognitive control training on symptoms of obsessive-compulsive disorder | Not adults with dep. |
| Calkins 2015 | The Effects of Computerized Cognitive Control Training on Community Adults with Depressed Mood | Not CCT |
| Calkins 2019 | Effects of a neurobehavioral intervention for depressive and obsessive-compulsive symptoms | Not CCT |
| Cameron 2020 | A pilot study examining the use of Goal Management Training in individuals with obsessive-compulsive disorder | Not CCT |
| Canesi 2021 | Editorial: Integrated Motor-Cognitive Aerobic Rehabilitation Approaches in Parkinson's Disease | Not RCT |
| Carballo-Garcia 2013 | Effects of non-pharmacological therapy on normal ageing and on cognitive decline: Reflections on treatment objectives | Not RCT |
| Carbone 2019 | Working Memory Training for Older Adults After Major Surgery: Benefits to Cognitive and Emotional Functioning | Not adults with dep. |
| Carcelen-Fraile 2022 | Cognitive Stimulation as Alternative Treatment to Improve Psychological Disorders in Patients with Mild Cognitive Impairment | Not CCT |
| Carlson 2020 | Mobile intervention for depression benefits middle aged and older adults | Not CCT |
| Carney 2000 | Change in heart rate variability during treatment for depression in patients with coronary heart disease | Not CCT |
| Chan 2020 | Cognitive training interventions and depression in mild cognitive impairment and dementia: a systematic review and meta-analysis of randomized controlled trials | Not RCT |
| Chariglione 2020 | Cognitive interventions and performance measures: A longitudinal study in elderly women | Not RCT |
| Chen 2020 | Efficacy of multidomain interventions to improve physical frailty, depression and cognition: data from cluster-randomized controlled trials. | Not adults with dep. |
| Chen 2021 | Non-pharmacological treatment for Parkinson disease patients with depression: a meta-analysis of repetitive transcranial magnetic stimulation and cognitive-behavioral treatment | Not RCT |
| Chen 2022 | Effects of Tabletop Games on Cognition in Older Adults: A Systematic Review and Meta-Analysis | Not CCT |
| Cheng 2018 | Comprehensive Rehabilitation Training Decreases Cognitive Impairment, Anxiety, and Depression in Poststroke Patients: A Randomized, Controlled Study | Not CCT |
| Cheng 2021 | Reminiscence therapy-based care program relieves post-stroke cognitive impairment, anxiety, and depression in acute ischemic stroke patients: a randomized, controlled study | Not CCT |
| Cheng 2022 | An exercise cum cognitive-behavioral intervention for older adults with chronic pain: A cluster-randomized controlled trial | Not CCT |
| Cheng 2022 | Do caregiver interventions improve outcomes in relatives with dementia and mild cognitive impairment? A comprehensive systematic review and meta-analysis | Not RCT |
| Chiang 2020 | Happiness or hopelessness in late life: A cluster RCT of the 3L-Mind-Training program among the institutionalized older people | Not adults with dep. |
| Chow 2020 | Effectiveness of psychosocial interventions among older adults with mild cognitive impairment: a systematic review and meta-analysis | Not RCT |
| Christensen 2019 | Effects of Individual Placement and Support Supplemented with Cognitive Remediation and Work-Focused Social Skills Training for People with Severe Mental Illness: A Randomized Clinical Trial | Not adults with dep. |
| Cimermanova 2011 | Personalized computer rehabilitation of cognitive functions: The results of the study with Cognifit program | Not RCT |
| Coelho 2020 | Exercise with music: An innovative approach to increase cognition and reduce depression in institutionalized elderly | Not CCT |
| Collier 2015 | Individual differences in response to prediction bias training | Not CCT |
| Concepcion 2021 | Moderating effect of the COVID-19 pandemic on the efficacy of cognitive stimulation. A controlled trial | Not RCT |
| Contreras 2014 | Is cognitive remediation therapy an effective intervention in enhancing vocational outcomes for people with severe mental illness? | Not RCT |
| Cooper 2014 | Training attention improves decision making in individuals with elevated self-reported depressive symptoms | Not CCT |
| Costas 2017 | Could cognitive remediation therapy increase psychological treatment adherence in depression disorders? the role of cognitive impairment (pilot study) | Not CCT |
| Costas-Carrera 2020 | Cognitive remediation therapy for depressive disorders: Pilot study on the improvement and maintenance of cognitive and psychosocial functioning | Not CCT |
| Cripe 2021 | Improved Mild Closed Head Traumatic Brain Injury Outcomes with a Brain-Computer Interface Amplified Cognitive Remediation Training | Not RCT |
| Crowe 2020 | Interpersonal and Social Rhythm Therapy for Patients With Major Depressive Disorder | Ineligible outcomes |
| Crowe 2021 | Patients' Perceptions of Functional Improvement in Psychotherapy for Mood Disorders | Not CCT |
| Daches 2019 | Training to inhibit negative content affects memory and rumination | Not adults with dep. |
| Daglas-Georgiou 2021 | Treatments for objective and subjective cognitive functioning in young people with depression: Systematic review of current evidence | Not RCT |
| Daley 2011 | Exercise-based cognitive therapy as a novel treatment for insomnia and depression | Not CCT |
| Dalgleish 2013 | Method-of-loci as a mnemonic device to facilitate access to self-affirming personal memories for individuals with depression | Not CCT |
| Dalgleish 2014 | A comparison of MEmory Specificity Training (MEST) to education and support (ES) in the treatment of recurrent depression: study protocol for a cluster randomised controlled trial | Not CCT |
| Dallaway 2021 | Concurrent brain endurance training improves endurance exercise performance Concurrent brain endurance training improves endurance exercise performance | Inappropriate control |
| Dammen 2022 | The Attention Training Technique Reduces Anxiety and Depression in Patients With Coronary Heart Disease: A Pilot Feasibility Study | Not CCT |
| Dammen 2022 | A feasibility study of the Attention Training Technique in a group format for anxiety and depression in outpatients with coronary heart disease | Not RCT |
| Datko 2022 | Increased Body Trusting Associated With Increased Insula Response to Interoception After Mindfulness Training in Patients With Depression and Anxiety | Not CCT |
| Davies 2021 | Altering Dynamics of Autonomic Processing Therapy (ADAPT) trial: a novel, targeted treatment for reducing anxiety in joint hypermobility | Not CCT |
| DeFreitas 2021 | Effects of transcranial direct current stimulation (tDCS) and concurrent cognitive training on episodic memory in patients with traumatic brain injury: A double-blind, randomised, placebo-controlled study | Inappropriate control |
| DeLuca 2021 | 'Online therapy' to reduce caregiver's distress and to stimulate post-severe acquired brain injury motor and cognitive recovery: A Sicilian hospital experience in the COVID era | Not RCT |
| DeWit 2021 | Memory Support System training in mild cognitive impairment: Predictors of learning and adherence | Not adults with dep. |
| Dehabadi 2021 | Comparison of the acceptance and commitment-based therapy and cognitive rehabilitation of working memory on anxiety and depression of girl | Not RCT |
| Dehn 2018 | Training in a comprehensive everyday-like virtual reality environment compared to computerized cognitive training for patients with depression | Not RCT |
| Dehn 2020 | Cognitive training in an everyday-like virtual reality enhances visual-spatial memory capacities in stroke survivors with visual field defects. | Not RCT |
| DeJong 1986 | Effectiveness of two psychological treatments for inpatients with severe and chronic depressions | Not CCT |
| Delahunt 2012 | Computer-based cognitive stimulation programs to remediate age-related cognitive decline: What makes a program effective? | Not adults with dep. |
| Demeyer 2020 | Cognitive Control Training in Healthy Older Adults: A Proof-of-Concept Study on the Effects on Cognitive Functioning, Emotion Regulation and Affect | Not adults with dep. |
| DePutter 2015 | Combining tDCS and Working Memory Training to Down Regulate State Rumination: A Single-Session Double Blind Sham-Controlled Trial | Not CCT |
| Dequanter 2020 | The Effectiveness of E-Health Solutions for Ageing With Cognitive Impairment: A Systematic Review | Not RCT |
| Dequanter 2021 | The Effectiveness of e-Health Solutions for Aging with Cognitive Impairment: A Systematic Review | Not RCT |
| DeRaedt 2012 | Changes in attentional processing of emotional information following mindfulness-based cognitive therapy in people with a history of depression: Towards an open attention for all emotional experiences | Not RCT |
| Derakhshan 2017 | Targeting cognitive control to reduce anxiety vulnerability: Implications for treatment efficacy | Not RCT |
| Desbordes 2012 | Effects of mindful-attention and compassion mediation training on amygdala response to emotional stimuli in an ordinary, non-meditative state | Not CCT |
| Diamond 2015 | Randomized controlled trial of a healthy brain ageing cognitive training program: effects on memory, mood, and sleep | Not adults with dep. |
| DiazBaquero 2022 | The Effectiveness of GRADIOR: A Neuropsychological Rehabilitation Program for People with Mild Cognitive Impairment and Mild Dementia. Results of a Randomized Controlled Trial After 4 and 12 Months of Treatment | Not adults with dep. |
| Dietrichkeit 2021 | Side effects of the metacognitive training for depression compared to a cognitive remediation training in patients with depression. | Inappropriate control |
| Dorociak 2022 | Cognitive and psychological improvements following CogSMART in veterans with mental health diagnoses | Not CCT |
| Doshi 2021 | Mindfulness-Based Training Does Not Improve Neuropsychological Outcomes in Mild Cognitive Impairment More Than Spontaneous Reversion Rates: A Randomized Controlled Trial | Not CCT |
| Douglas 2015 | Psychosocial and cognitive remediation for recurrent mood disorders: Pilot findings and project outline | Not adults with dep. |
| Douglas 2020 | Cognitive Remediation for Outpatients with Recurrent Mood Disorders: A Feasibility Study | Not adults with dep. |
| Douglas 2020 | Continuing the debate: Cognitive enhancement therapy for mood disorders | Not RCT |
| Douglas 2022 | Cognitive remediation for mood disorders: Clinical, cognitive and psychological factors related to treatment response | NOT RCT |
| Douglas 2022 | A randomised controlled trial of psychotherapy and cognitive remediation to target cognition in mood disorders | Inappropriate control |
| Douglas 2022 | Randomised controlled trial of Interpersonal and Social Rhythm Therapy and group-based Cognitive Remediation versus Interpersonal and Social Rhythm Therapy alone for mood disorders: study protocol | Not RCT |
| Dubuson 2021 | Transcranial direct current stimulation combined with alcohol cue inhibitory control training reduces the risk of early alcohol relapse: A randomized placebo-controlled clinical trial | Not CCT |
| Duff 2022 | Computerized Cognitive Training in Amnestic Mild Cognitive Impairment: A Randomized Clinical Trial | Not adults with dep. |
| Dupuy 2021 | COVEPIC (Cognitive and spOrt Virtual EPIC training) investigating the effects of home-based physical exercise and cognitive training on cognitive and physical functions in community-dwelling older adults: study protocol of a randomized single-blinded clinical trial | Not adults with dep. |
| Dwolatzky 2021 | Changes in brain volume resulting from cognitive intervention by means of the feuerstein instrumental enrichment program in older adults with mild cognitive impairment (Mci): A pilot study | Not RCT |
| Eigenhuis 2017 | Feasibility and Effectiveness of Memory Specificity Training in Depressed Outpatients: A Pilot Study | Not CCT |
| Ekkers 2011 | Competitive Memory Training for treating depression and rumination in depressed older adults: a randomized controlled trial | Not CCT |
| Ekman 2020 | Evaluation of a Novel Psychological Intervention Tailored for Patients With Early Cognitive Impairment (PIPCI): Study Protocol of a Randomized Controlled Trial | Not RCT |
| Elgamal 2007 | Successful computer-assisted cognitive remediation therapy in patients with unipolar depression: a proof of principle study | Not RCT |
| Elliott 2016 | Computer-based cognitive training for patients with unipolar depression | Data not available |
| Erten 2018 | Memory Specificity Training for Depression and Posttraumatic Stress Disorder: A Promising Therapeutic Intervention | Not RCT |
| Estrada-Plana 2021 | Cognitive training with modern board and card games in healthy older adults: two randomized controlled trials | Not CCT |
| Everaert 2020 | Mapping dynamic interactions among cognitive biases in depression. | Not RCT |
| Fairchild 2008 | Team (training to enhance adult memory): Investigation of the efficacy of an in-home memory enhancement program | Not CCT |
| Farahimanesh 2021 | Autobiographical memory bias in cancer-related post traumatic stress disorder and the effectiveness of competitive memory training. | Not CCT |
| Farahimanesh 2021 | Comparing the Efficacy of Competitive Memory Training (COMET) and Memory Specificity Training (MEST) on Posttraumatic Stress Disorder Among Newly Diagnosed Cancer Patients | Not CCT |
| Farkhani 2022 | Computerized Cognitive Training Interventions in Major Depression | Not RCT |
| Farragher 2020 | Cognitive interventions for adults with chronic kidney disease: Protocol for a scoping review | Not RCT |
| Feinstein 2020 | Study protocol: improving cognition in people with progressive multiple sclerosis: a multi-arm, randomized, blinded, sham-controlled trial of cognitive rehabilitation and aerobic exercise (COGEx). | Not RCT |
| Feinstein 2020 | Improving cognition in people with progressive multiple sclerosis: A multi-arm, randomized, blinded, sham-controlled trial of cognitive rehabilitation and aerobic exercise (COGEx) | Not RCT |
| Felix 2019 | Depressive symptoms may hamper long-term maintenance of cognitive training gains and downstream functional benefits in older adults at-risk for dementia: Active 10-year follow-up study | Not adults with dep. |
| Fellman 2020 | Training working memory updating in Parkinson's disease: A randomised controlled trial. | Not adults with dep. |
| FerreiraSantana 2016 | Maintenance of functional capacity in cognitive stimulation subgroups | Not RCT |
| Ford 2020 | An investigation of the relation between mindfulness and self-esteem stability | Not CCT |
| Fritze 1988 | Cognitive training adjunctive to pharmacotherapy in schizophrenia and depression: a pilot study on the lateralization hypothesis of schizophrenia and depression and on cognitive therapy as adjunctive to pharmacotherapy | Not CCT |
| Fry 1984 | Cognitive training and cognitive-behavioral variables in the treatment of depression in the elderly | Not CCT |
| Galletly 2016 | Assessing the Effects of Repetitive Transcranial Magnetic Stimulation on Cognition in Major Depressive Disorder Using Computerized Cognitive Testing | Not CCT |
| Gamito 2020 | Virtual Reality-Based Cognitive Stimulation to Improve Cognitive Functioning in Community Elderly: A Controlled Study | Not adults with dep. |
| Gandy 2020 | A feasibility trial of an internet-delivered psychological intervention to manage mental health and functional outcomes in neurological disorders | Not RCT |
| Gao 2021 | Effects of Virtual Reality-Based Intervention on Cognition, Motor Function, Mood, and Activities of Daily Living in Patients With Chronic Stroke: A Systematic Review and Meta-Analysis of Randomized Controlled Trials | Not RCT |
| Garcia-Molina 2022 | Neuropsychological rehabilitation for post-COVID-19 syndrome: Results of a clinical program and six-month follow up | Not RCT |
| Garcia-Palacios 2015 | Integrating virtual reality with activity management for the treatment of fibromyalgia: Acceptability and preliminary efficacy | Not CCT |
| Georgiades 2007 | Changes in depressive symptoms and glycemic control in diabetes mellitus | Not CCT |
| Gil 2022 | Effectiveness of Reminiscence Therapy versus Cognitive Stimulation Therapy in Older Adults with Cognitive Decline: A Quasi-Experimental Pilot Study | Not RCT |
| Giudici 2020 | Effect of long-term omega-3 supplementation and a lifestyle multidomain intervention on intrinsic capacity among community-dwelling older adults: Secondary analysis of a randomized, placebo-controlled trial (MAPT study) | Inappropriate control |
| Glenthøj 2015 | The FOCUS trial: cognitive remediation plus standard treatment versus standard treatment for patients at ultra-high risk for psychosis: study protocol for a randomised controlled trial | Not adults with dep. |
| Godara 2021 | Looking for carrots, watching out for sticks: A gaze-contingent approach towards training contextual goal-dependent affective attention flexibility. | Not CCT |
| Gomez-Soria 2020 | Cognitive stimulation program in mild cognitive impairment A randomized controlled trial | Not adults with dep. |
| Gomez-Soria 2021 | Long-term effect analysis of a cognitive stimulation program in mild cognitive impairment elderly in Primary Care: A randomized controlled trial | Not adults with dep. |
| Gomez-Soria 2022 | Effectiveness of Personalized Cognitive Stimulation in Older Adults with Mild Possible Cognitive Impairment: A 12-month Follow-up Cognitive Stimulation in Mild Cognitive Impairment | Not CCT |
| Gonzalez-Palau 2014 | The effects of a computer-based cognitive and physical training program in a healthy and mildly cognitive impaired aging sample | Inappropriate control |
| Granland 2022 | "Train Your Brain" Cognitive Intervention Group Program for Singaporean Older Adult Patients With Mild Cognitive Impairment: A Pilot Feasibility Study | Not RCT |
| Grinberg 2021 | Mobile cognitive training for the cognitive symptoms of depression in young adults: A double-blind, randomized pilot study with active control | Inappropriate control |
| Gromisch 2020 | The effects of cognitive-focused interventions on cognition and psychological well-being in persons with multiple sclerosis: A meta-analysis | Not RCT |
| Gumport 2018 | Patient learning of treatment contents in cognitive therapy | Not CCT |
| Gyurak 2013 | Cognitive-affective remediation training intervention in anxiety and depression | Data not available |
| Haeffel 2010 | When self-help is no help: traditional cognitive skills training does not prevent depressive symptoms in people who ruminate | Not CCT |
| Hagen 2020 | Predictors of Long-Term Improvement Following Cognitive Remediation in a Sample With Elevated Depressive Symptoms | Not RCT |
| Hagen 2021 | Goal Management Training and Computerized Cognitive Training in Depression-a 2-Year Follow-Up of a Randomized Controlled Trial | Already included in previous update |
| Hallford 2020 | A study protocol for a randomised trial of adjunct computerised memory specificity training (c-MeST) for major depression in youth: targeting cognitive mechanisms to enhance usual care outcomes in mental health settings | Not RCT |
| Hallford 2021 | Computerized Memory Specificity Training (c-MeST) for major depression: A randomised controlled trial | Not CCT |
| Hallgren 2016 | Exercise and internet-based cognitive-behavioural therapy for depression: Multicentre randomised controlled trial with 12-month follow-up | Not CCT |
| Hammar 2020 | A pilot study of cognitive remediation in remitted major depressive disorder patients | Inappropriate control |
| Hammar 2022 | A pilot study of cognitive remediation in remitted major depressive disorder patients | Not RCT |
| Han 2018 | Effects of a Manualized, Group-Based Cognitive Training on Cognitive Performance in Community-Dwelling Elderly | Not CCT |
| Harvey 2020 | Training engagement, baseline cognitive functioning, and cognitive gains with computerized cognitive training: A cross-diagnostic study | Not RCT |
| Haukaas 2018 | A Randomized Controlled Trial Comparing the Attention Training Technique and Mindful Self-Compassion for Students With Symptoms of Depression and Anxiety | Not CCT |
| Hirsch 2021 | Internet-delivered interpretation training reduces worry and anxiety in individuals with generalized anxiety disorder: A randomized controlled experiment | Not CCT |
| Hitchcock 2015 | Memory Flexibility training (MemFlex) to reduce depressive symptomatology in individuals with major depressive disorder: study protocol for a randomised controlled trial | Not CCT |
| Hitchcock 2018 | Study protocol for a randomised, controlled platform trial estimating the effect of autobiographical Memory Flexibility training (MemFlex) on relapse of recurrent major depressive disorder | Not CCT |
| Hitchcock 2018 | A randomised controlled trial of memory flexibility training (MemFlex) to enhance memory flexibility and reduce depressive symptomatology in individuals with major depressive disorder | Not CCT |
| Hitchcock 2021 | A randomized, controlled proof-of-concept trial evaluating durable effects of memory flexibility training (MemFlex) on autobiographical memory distortions and on relapse of recurrent major depressive disorder over 12 months | Not CCT |
| Hoch 2018 | Brain connectivity changes associated with a cognitive-emotional training intervention for depression | Not CCT |
| Hoch 2019 | Initial Evidence for Brain Plasticity Following a Digital Therapeutic Intervention for Depression | Not RCT |
| Holmqvist 2021 | Does Intensive Training of Attention Influence Cognitive Fatigability in Patients With Acquired Brain Injury? | Not adults with dep. |
| Hoorelbeke 2021 | Online Cognitive Control Training for Remitted Depressed Individuals: A Replication and Extension Study | Not CCT |
| Hoorelbeke 2021 | Preventing recurrence of depression: Long-term effects of a randomized controlled trial on cognitive control training for remitted depressed patients | Ineligible outcomes |
| Hoorelbeke 2022 | Regaining control of your emotions? Investigating the mechanisms underlying effects of cognitive control training for remitted depressed patients | Ineligible outcomes |
| Houston 2021 | Individualized Web-Based Attention Training With Evidence-Based Counseling to Address HIV Treatment Adherence and Psychological Distress: Exploratory Cohort Study | Not RCT |
| Huang 2016 | Evaluation of a combined cognitive-behavioural and exercise intervention to manage fear of falling among elderly residents in nursing homes | Not CCT |
| Ikebuchi 2017 | Does improvement of cognitive functioning by cognitive remediation therapy effect work outcomes in severe mental illness? A secondary analysis of a randomized controlled trial | Inappropriate control |
| Jagawat 2022 | A Double-Blind Randomized Sham Control Study to Assess the Effects of rTMS (Repetitive Transcranial Magnetic Stimulation) on Executive Functioning in Treatment Resistant Depression | Not CCT |
| Jannati 2020 | Effectiveness of an app-based cognitive behavioral therapy program for postpartum depression in primary care: A randomized controlled trial | Not CCT |
| Javidi 2021 | A randomized controlled trial of self-compassion versus cognitive therapy for complex psychopathologies | Not CCT |
| Jaywant 2020 | The Structural and Functional Neuroanatomy of Post-Stroke Depression and Executive Dysfunction: A Review of Neuroimaging Findings and Implications for Treatment | Not RCT |
| Jefferies 2021 | Rethinking cognitive training: The moderating roles of emotional vulnerability and perceived cognitive impact of training in high worriers | Not adults with dep. |
| Jelinek 2016 | Efficacy of metacognitive training for depression: A randomized controlled trial | Not CCT |
| Jeong 2021 | Multi-Component Intervention Program on Habitual Physical Activity Parameters and Cognitive Function in Patients with Mild Cognitive Impairment: A Randomized Controlled Trial | Not adults with dep. |
| Jimenez-Morales 2021 | Cognitive rehabilitation program in patients with multiple sclerosis: A pilot study | Not CCT |
| Jones 2013 | Online cognitive behaviour training for the prevention of postnatal depression in at-risk mothers: a randomised controlled trial protocol | Not CCT |
| Jopling 2020 | Effects of working memory training on cognitive, affective, and biological responses to stress in major depression: A novel cognitive bias modification protocol | Not CCT |
| Jopowicz 2022 | Cognitive and Physical Intervention in Metals' Dysfunction and Neurodegeneration | Not RCT |
| Josephs 2020 | An investigation of Cogmed working memory training for neurological surgery patients | Not RCT |
| Joshi 2021 | Expanding the scope of auditory-based targeted cognitive training for transdiagnostic applications in va settings: A pilot study | Inappropriate control |
| Jung 2019 | Preliminary results from a randomized controlled trial of serious games for the improvement of cognitive function in chronic stroke survivors | Not adults with dep. |
| Justo-Henriques 2021 | Effect of long-term individual cognitive stimulation intervention for people with mild neurocognitive disorder | Not RCT |
| Kado 2002 | Computer-assisted cognitive rehabilitation treatment and outcomes | Not RCT |
| Kalbe 2020 | Enhancement of executive functions but not memory by multidomain group cognitive training in patients with Parkinson's disease and mild cognitive impairment: A multicenter randomized controlled trial | Not adults with dep. |
| Kamble 2021 | Efficacy of virtual reality induced environmental and habitual navigation on psychological, cognitive function that impacts on physical recovery in patients with stroke | Not CCT |
| Kanemaru 2019 | The Twelve Months' Follow-up after the Intervention Using a Rehabilitation Program of Physical and Cognitive Rec-Xercise (Repcrec) for the Elderly with Mild Cognitive Impairment (MCI) | Not adults with dep. |
| Keefe 2019 | A randomized, double-blind, controlled, 6-week trial to assess a novel digital intervention designed to improve cognitive dysfunction as adjunct therapy to antidepressant medication in adults with major depressive disorder | Data not available |
| Keil 2021 | Morning bright light therapy for sleep to augment cognitive rehabilitation in Veterans with comorbid traumatic brain injury and post-traumatic stress disorder: A pilot study | Not CCT |
| Kelberer 2021 | Comparing attention-training methods in attention bias modification for depression | Not CCT |
| Keller 2021 | fMRI Neurofeedback-Enhanced Cognitive Reappraisal Training in Depression: A Double-Blind Comparison of Left and Right vlPFC Regulation | Not CCT |
| Khanipour 2022 | The investigation of the effects of occupation-based intervention on anxiety, depression, and sleep quality of subjects with hand and upper extremity burns: A randomized clinical trial | Not CCT |
| Khayati 2020 | The Effect of Cognitive-Behavioral Training Versus Conventional Training on Self-care and Depression Severity in Heart Failure Patients with Depression: A Randomized Clinical Trial. | Not CCT |
| Kidd 2020 | A comparison of compensatory and restorative cognitive interventions in early psychosis | Not adults with dep. |
| Kim 2020 | Cognitive Improvement Effects of Electroacupuncture Combined with Computer-Based Cognitive Rehabilitation in Patients with Mild Cognitive Impairment: A Randomized Controlled Trial | Not adults with dep. |
| Kim 2021 | Efficacy of Smart Speaker-Based Metamemory Training in Older Adults: Case-Control Cohort Study | Not adults with dep. |
| Kim 2022 | Effects of a cognitive rehabilitation program based on mnemonic skills and memory compensatory strategies for older adults: A pilot study | Not CCT |
| Kim 2022 | Effectiveness of group metacognitive training and cognitive-behavioural therapy in a transdiagnostic manner for young patients with psychotic and non-psychotic disorders | Not CCT |
| Kleijn 2021 | A randomized controlled trial on the efficacy of life review therapy targeting incurably ill cancer patients: do their informal caregivers benefit? | Not CCT |
| Kletzel 2021 | Effectiveness of Brain Gaming in Older Adults With Cognitive Impairments: A Systematic Review and Meta-Analysis | Not RCT |
| Knight 2017 | Psychosocial Dysfunction in Major Depressive Disorder-Rationale, Design, and Characteristics of the Cognitive and Emotional Recovery Training Program for Depression (CERT-D) | Inappropriate control |
| Knight 2019 | Contemporary methods of improving cognitive dysfunction in clinical depression | Not RCT |
| Knight 2021 | Psychological training to improve psychosocial function in patients with major depressive disorder: A randomised clinical trial | Not CCT |
| Koivisto 2020 | Gamification for Older Adults: A Systematic Literature Review. | Not RCT |
| Kolbe 2021 | Use of virtual reality in the inpatient rehabilitation of COVID-19 patients | Not RCT |
| Korrelboom 2009 | Competitive memory training for treating low self-esteem: A pilot study in a routine clinical setting | Not CCT |
| Korrelboom 2012 | Competitive memory training (COMET) for treating low self-esteem in patients with depressive disorders: a randomized clinical trial | Not CCT |
| Korrelboom 2022 | The Effectiveness of Transdiagnostic Applications of Competitive Memory Training (COMET) on Low Self-Esteem and Comorbid Depression: A Meta-analysis of Randomized Controlled Trials | Not RCT |
| Kotov 2020 | Possibilities for Correcting Emotional and Behavioral Impairments in Stroke Patients during Rehabilitation Therapy | Not adults with dep. |
| Kotov 2021 | The effectiveness of multimodal stimulation in the correction of cognitive and affective disorders in poststroke patients | Inappropriate control |
| Kraiwong 2021 | Effects of physical-cognitive training on physical and psychological functions among older adults with type 2 diabetes and balance impairment: a randomized controlled trial | Not CCT |
| Krause-Sorio 2022 | Yoga Prevents Gray Matter Atrophy in Women at Risk for Alzheimer's Disease: A Randomized Controlled Trial | Not CCT |
| Krings 2022 | Is the combination of behavioral activation and attention training technique effective to reduce depressive symptomatology? A multiple case study | Not RCT |
| Krause-Sorio 2022 | Yoga Prevents Gray Matter Atrophy in Women at Risk for Alzheimer's Disease: A Randomized Controlled Trial | Not CCT |
| Kristensen 2020 | Effects of cognitive remediation on white matter in individuals at ultra-high risk for psychosis-a randomized, controlled clinical trial | Inappropriate control |
| Lampit 2022 | Computerized cognitive training in people with depression: a protocol for a systematic review and meta-analysis | Not RCT |
| Lang 2022 | Implementation of a Telephone-Based Cognitive Rehabilitation Group for Older Adult Veterans | Not RCT |
| Lau 2021 | Computer-Assisted Cognitive Training for Patients with Severe Mental Illness: a Retrospective Study | Not RCT |
| Lee 2013 | Cognitive remediation improves memory and psychosocial functioning in first-episode psychiatric out-patients | Not CCT |
| Lee 2019 | Combination Theta-Burst Stimulation and Cognitive Training for Youth Depression | Data not available |
| Lee 2020 | Effectiveness of virtual reality based cognitive rehabilitation on cognitive function, motivation and depression in stroke patients | Not adults with dep. |
| Lee 2020 | Qigong reduces depressive symptoms of Taiwanese elderly with chronic physical illness: A randomized controlled trial | Not CCT |
| Lee 2021 | Digital healthcare for memory training | Not RCT |
| Legemaat 2021 | Effectiveness of cognitive remediation in depression: a meta-analysis. | Not RCT |
| Lemos 2016 | A neuropsychological group rehabilitation program with institutionalized elderly | Not adults with dep. |
| LeMoult 2014 | Neural effects of working memory training in major depressive disorder: Facilitating scientific translation | Not RCT |
| Lenze 2019 | MECHANISMS INFORMING INTERVENTIONS: NEW APPROACHES TO TREATING LATE-LIFE DEPRESSION: Session 107 | Not RCT |
| Li 2017 | Mood improvement boosted cognitive training gains through hippocampus-amygdala connectivity in elderly with subjective memory complaints | Not adults with dep. |
| Lin 2005 | Influence of post-stroke depression on the effects of rehabilitation. | Not CCT |
| Lin 2020 | Effects of Creative Expressive Arts-based Storytelling (CrEAS) programme on older adults with mild cognitive impairment: Protocol for a randomised, controlled three-arm trial | Not RCT |
| Lindenmayer 2008 | A randomized controlled trial of cognitive remediation among inpatients with persistent mental illness | Not adults with dep. |
| Liossi 2020 | Internet-delivered attentional bias modification training (iABMT) for the management of chronic musculoskeletal pain: a protocol for a randomised controlled trial | Not RCT |
| Listunova 2018 | Cognitive Impairment Along the Course of Depression: Non-Pharmacological Treatment Options | Not RCT |
| Lohman 2013 | Depressive symptoms and memory performance among older adults: results from the ACTIVE memory training intervention | Not CCT |
| Loo 2015 | Improvement of psychotherapeutic techniques through TDCS in depressive patients | Not CCT |
| Lopes 2016 | Cognitive training in the elderly and its effect on the executive functions | Not CCT |
| Lu 2020 | Qigong for the treatment of depressive symptoms: Preliminary evidence of neurobiological mechanisms | Not CCT |
| Lubarda 2017 | Improving management of major depressive disorder through virtual patient simulation | Not CCT |
| Maggio 2022 | Cognitive rehabilitation outcomes in patients with Multiple sclerosis: Preliminary data about the potential role of personality traits | Not RCT |
| Maier 2020 | Adaptive conjunctive cognitive training (ACCT) in virtual reality for chronic stroke patients: a randomized controlled pilot trial | Not adults with dep. |
| Mansbach 2020 | Integrating Working Memory Exercises with Nursing Home Rehabilitation to Achieve "better, Faster" Functional Outcomes | Data not available |
| Marciniak 2020 | The Effect of Mindfulness-Based Stress Reduction (MBSR) on Depression, Cognition, and Immunity in Mild Cognitive Impairment: A Pilot Feasibility Study | Not adults with dep. |
| Martens 2019 | The transportability of Memory Specificity Training (MeST): adapting an intervention derived from experimental psychology to routine clinical practices | Not CCT |
| Martin 2018 | Clinical pilot study of transcranial direct current stimulation combined with Cognitive Emotional Training for medication resistant depression | Not RCT |
| Maruyama 2021 | Improvement in executive functioning after Goal-Oriented Attentional Self-Regulation training is associated with reduction in PTSD hyperarousal symptoms among veterans with comorbid PTSD and mild TBI | Not CCT |
| Matos 2017 | Psychological and physiological effects of compassionate mind training: A pilot randomised controlled study | Not CCT |
| Mattioli 2011 | "Efficacy and specificity of intensive cognitive rehabilitation of attention and executive functions in multiple sclerosis": Erratum | Data also elsewhere |
| McGlinchey 2014 | Internet-based cognitive training enhances attention and functional outcomes in OEF/OIF/ OND veterans | Not adults with dep. |
| McGurk 2005 | Cognitive training and supported employment for persons with severe mental illness: one-year results from a randomized controlled trial | Not adults with dep. |
| McGurk 2007 | Cognitive training for supported employment: 2-3 year outcomes of a randomized controlled trial | Not adults with dep. |
| McLaughlin 2018 | The Feasibility and Potential Impact of Brain Training Games on Cognitive and Emotional Functioning in Middle-Aged Adults | Not adults with dep. |
| Meca-Lallana 2020 | A Pilot Study to Explore Patient Satisfaction With a Virtual Rehabilitation Program in Multiple Sclerosis: The RehabVR Study Protocol | Not RCT |
| Merom 2008 | Promoting walking as an adjunct intervention to group cognitive behavioral therapy for anxiety disorders--A pilot group randomized trial | Not CCT |
| Meuleman 2021 | A randomized controlled trial of cognitive control training (CCT) as an add-on treatment for late-life depression: a study protocol | Not RCT |
| Meusel 2009 | Evidence for sustained improvement in memory deficits following computer-assisted cognitive remediation in patients with a mood disorder | Not RCT |
| Meusel 2010 | Improvement in memory deficits following computer assisted cognitive remediation in patients with a mood disorder | Not RCT |
| Meusel 2013 | Neural correlates of cognitive remediation in patients with mood disorders | Not RCT |
| Mewton 2020 | A randomised double-blind trial of cognitive training for the prevention of psychopathology in at-risk youth | Not adults with dep. |
| Miles 2014 | The effects of gentle yoga vs. cognitive behavioral therapy on physical and psychological symptoms; neurocognitive functioning; and physiology in women with Fibromyalgia | Not CCT |
| Millan-Calenti 2015 | Efficacy of a computerized cognitive training application on cognition and depressive symptomatology in a group of healthy older adults: A randomized controlled trial | Not adults with dep. |
| Mohlman 2017 | Initial outcomes of a combined cognitive-behavioral therapy and attention process training intervention for older adults with Parkinson's disease | Not RCT |
| Moncrief 2021 | Self-rated executive dysfunction in adults with epilepsy and effects of a cognitive-behavioral intervention (HOBSCOTCH) | Not CCT |
| Morimoto 2012 | Neuroplasticity-based computerized cognitive remediation for geriatric depression | Not RCT |
| Morimoto 2013 | Computerized cognitive remediation for geriatric depression | Not RCT |
| Morimoto 2014 | Neuroplasticity-based computerized cognitive remediation for treatment resistant geriatric depression | Inappropriate control |
| Morimoto 2014 | Neuroplasticity-based computerized cognitive remediation for treatment-resistant geriatric depression | Not RCT |
| Morimoto 2016 | Executive Dysfunction Predicts Treatment Response to Neuroplasticity-Based Computerized Cognitive Remediation (nCCR-GD) in Elderly Patients with Major Depression | Not RCT |
| Moritz 2018 | Metacognitive Training for Depression (D-MCT) reduces false memories in depression. A randomized controlled trial | Not CCT |
| Moshier 2015 | Assessing the effects of depressed mood and cognitive control training on memory confidence and accuracy following repeated checking | Not CCT |
| Moshier 2016 | Cognitive control training as an adjunct to behavioral activation therapy in the treatment of depression | Not CCT |
| Moshier 2017 | Behavioral activation treatment for major depression: A randomized trial of the efficacy of augmentation with cognitive control training | Not CCT |
| Motter 2015 | Computerized cognitive training for major depressive disorder: What's next? | Not RCT |
| Motter 2019 | Computerized cognitive training in young adults with depressive symptoms: Effects on mood, cognition, and everyday functioning | Inappropriate control |
| Mroczkowska 2019 | Neuropsychological rehabilitation of patients with symptoms of depression after ischemic stroke | Not adults with dep. |
| Mueser 2013 | Effectiveness of cognitive remediation in improving employment outcomes in Supported Employment Non Responders | Not adults with dep. |
| Muller 2012 | The effect of cognitive remediation with biotic designed computer based training (CBT) VS. Non biotic designed CBT on global working memory of depressive patients | Not RCT |
| Munger 2021 | Impact of cognitive rehabilitation on quality of life in patients with multiple sclerosis | Not RCT |
| Myklebost 2021 | An open pilot study of an internet-delivered intervention targeting self-perceived residual cognitive symptoms after major depressive disorder | Not RCT |
| Myklebost 2022 | Developing an internet-delivered intervention targeting residual cognitive symptoms after major depressive disorder: a person-based approach | Not RCT |
| Myklebost 2022 | Predictors of Treatment Response to an Internet-Delivered Intervention Targeting Residual Cognitive Symptoms After Major Depressive Disorder | Not RCT |
| Naismith 2009 | Can a healthy brain aging group program improve cognition in late-life depression? | Not RCT |
| Naismith 2010 | Cognitive training in affective disorders improves memory: a preliminary study using the NEAR approach | Not RCT |
| Naismith 2011 | Enhancing memory in late-life depression: the effects of a combined psychoeducation and cognitive training program | Not RCT |
| Naismith 2016 | Moving beyond mood: Is it time to recommend cognitive training for depression in older adults? | Not RCT |
| Navarra-Ventura 2021 | Virtual Reality-Based Early Neurocognitive Stimulation in Critically Ill Patients: A Pilot Randomized Clinical Trial | Not adults with dep. |
| Nejati 2019 | Cognitive training for modifying interpretation and attention bias in depression: Relevance to mood improvement and implications for cognitive intervention in depression | Not CCT |
| Neshat-Doost 2013 | Enhancing autobiographical memory specificity through cognitive training: An intervention for depression translated from basic science | Not CCT |
| Netto 2012 | Memory rehabilitation of elderly adults with mnemonic complaints and depressive symptoms: A pilot study | Not RCT |
| Neuvonen 2022 | Associations of Depressive Symptoms and Cognition in the FINGER Trial: A Secondary Analysis of a Randomised Clinical Trial | Not CCT |
| Ng 2017 | Multi-Domains Lifestyle Interventions Reduces Depressive Symptoms among Frail and Pre-Frail Older Persons: Randomized Controlled Trial | Not CCT |
| Nieman 2015 | Cognitive remediation in psychiatric patients with an online cognitive game and assessment tool | Not RCT |
| Nijmeijer 2021 | Foreign Language Learning as Cognitive Training to Prevent Old Age Disorders? Protocol of a Randomized Controlled Trial of Language Training vs. Musical Training and Social Interaction in Elderly With Subjective Cognitive Decline | Not RCT |
| Nikolin 2019 | Clinical effects of transcranial direct current stimulation combined with cognitive emotional training in patients with treatment resistant depression | Not RCT |
| Nikolin 2020 | Assessing neurophysiological changes associated with combined transcranial direct current stimulation and cognitive-emotional training for treatment-resistant depression | Not RCT |
| Niles 2020 | Randomized controlled trial testing mobile-based attention-bias modification for posttraumatic stress using personalized word stimuli | Not CCT |
| Nouchi 2016 | Small Acute Benefits of 4 Weeks Processing Speed Training Games on Processing Speed and Inhibition Performance and Depressive Mood in the Healthy Elderly People: Evidence from a Randomized Control Trial | Not adults with dep. |
| Nouchi 2021 | Brain Training Game Improved Cognitive Health | Not adults with dep. |
| Nousia 2022 | The Effectiveness of Non-Invasive Brain Stimulation Alone or Combined with Cognitive Training on the Cognitive Performance of Patients With Traumatic Brain Injury: Alpha Systematic Review | Not RCT |
| Oh 2021 | Long-term effect of a 24-week multicomponent intervention on physical performance and frailty in community-dwelling older adults | Not RCT |
| OliveiradeLimaQueiroz 2013 | Prevention of cognitive impairment through a cognitive stimulation and rehabilitation program mediated by computers and internet | Not adults with dep. |
| Olukolade 2017 | Efficacy of Cognitive Rehabilitation Therapy on Poststroke Depression among Survivors of First Stroke Attack in Ibadan, Nigeria | Not CCT |
| Onraedt 2014 | Training working memory to reduce rumination | Not adults with dep. |
| Optale 2010 | Controlling memory impairment in elderly adults using virtual reality memory training: A randomized controlled pilot study | Not adults with dep. |
| Ordonez 2017 | Actively station: Effects on global cognition of mature adults and healthy elderly program using eletronic games | Not adults with dep. |
| Orel 2014 | Training of attention in patients with remitting-relapsing multiple sclerosis | Not adults with dep. |
| Otero 2021 | Development of a Videogame for the Promotion of Active Aging Through Depression Prevention, Healthy Lifestyle Habits, and Cognitive Stimulation for Middle-to-Older Aged Adults | Not RCT |
| Ouyang 2001 | The effect of sports training with cognitive therapy about mild depression in university students | Not CCT |
| Overman 2021 | Inducing Affective Learning Biases with Cognitive Training and Prefrontal tDCS: A Proof-of-Concept Study | Not CCT |
| Owens 2013 | Improving attention control in dysphoria through cognitive training: transfer effects on working memory capacity and filtering efficiency | Not RCT |
| Ownby 2021 | Cognitive training with and without transcranial direct current stimulation in older adults with HIV-related cognitive deficits: Delayed impact on mood | Not CCT |
| Pabel 2018 | Olfactory training with clients suffering from depressive disorders | Not CCT |
| Pabel 2020 | Null Effect of Olfactory Training With Patients Suffering From Depressive Disorders-An Exploratory Randomized Controlled Clinical Trial | Not CCT |
| Palladini 2022 | Cognitive remediation therapy for post-acute persistent cognitive deficits in COVID-19 survivors: A proof-of-concept study | Not RCT |
| Pang 2021 | Effects of Smartphone-Based Compensatory Cognitive Training and Physical Activity on Cognition, Depression, and Self-Esteem in Women with Subjective Cognitive Decline | Not CCT |
| Papageorgiou 2000 | Treatment of recurrent major depression with attention training | Not RCT |
| Parial 2022 | Dual-task Zumba Gold for improving the cognition of people with mild cognitive impairment: A pilot randomized controlled trial | Not CCT |
| Parisi 2014 | Depressive symptoms and inductive reasoning performance: findings from the ACTIVE reasoning training intervention | Not CCT |
| Park 2009 | The effects of cognitive training on community-dwelling elderly Koreans | Not CCT |
| Park 2020 | South Korean Study to Prevent Cognitive Impairment and Protect Brain Health Through Lifestyle Intervention in At-Risk Elderly People: Protocol of a Multicenter, Randomized Controlled Feasibility Trial | Not RCT |
| Park 2021 | The Humanoid Robot Sil-Bot in a Cognitive Training Program for Community-Dwelling Elderly People with Mild Cognitive Impairment during the COVID-19 Pandemic: A Randomized Controlled Trial | Not adults with dep. |
| Parola 2017 | The effect of a cognitive stimulation program on institutionalized elderly: A randomized controlled trial | Not CCT |
| Paulewicz 2015 | Memory bias training by means of the emotional short-term memory task | Not CCT |
| Paulo 2012 | Elderly individuals with diabetes: Adding cognitive training to psychoeducational intervention | Not adults with dep. |
| Payzieva 2014 | NIRS Study of the Effects of Computerized Brain Training Games for Cognitive Rehabilitation of Major Depressive Disorder Patients in Remission: A Pilot Study | Not RCT |
| Peng 2019 | HRV evidence for the improvement of emotion regulation in university students with depression tendency by working memory training | Not RCT |
| Perez-Martin 2017 | Efficacy of a short cognitive training program in patients with multiple sclerosis | Not adults with dep. |
| Perlman 2022 | Inferences training affects memory, rumination, and mood | Not CCT |
| Perna 2010 | The effect of a cognitive behavioral exercise intervention on clinical depression in a multiethnic sample of women with breast cancer: A randomized controlled trial | Not CCT |
| Perna 2017 | Short-Term Psychiatric Rehabilitation in Major Depressive and Bipolar Disorders: Neuropsychological-Psychosocial Outcomes | Not CCT |
| Pile 2020 | Harnessing Mental Imagery and Enhancing Memory Specificity: Developing a Brief Early Intervention for Depressive Symptoms in Adolescence | Not adults with dep. |
| Pino 2019 | The humanoid robot nao as trainer in a memory program for elderly people with mild cognitive impairment | Not RCT |
| Plechata 2021 | Cognitive Remediation in Virtual Environments for Patients with Schizophrenia and Major Depressive Disorder: A Feasibility Study | Not adults with dep. |
| Podda 2022 | Focus on neglected features of cognitive rehabilitation in MS: Setting and mode of the treatment | Not RCT |
| Porter 2017 | Cognitive and affective remediation training for mood disorders | Not RCT |
| Powell 2013 | Effectiveness of a web-based cognitive-behavioral tool to improve mental well-being in the general population: Randomized controlled trial | Not CCT |
| Pratap 2018 | Using Mobile Apps to Assess and Treat Depression in Hispanic and Latino Populations: Fully Remote Randomized Clinical Trial | Ineligible outcomes |
| Preiss 2009 | Cognitive functions in patients with unipolar depression in remission - Main project results | Not RCT |
| Preiss 2012 | Computerised cognitive training relieves depression and improves executive control in depressive disorder | Not RCT |
| Priyamvada 2015 | Cognitive rehabilitation of attention and memory in depression | Not RCT |
| Protopopescu 2022 | A Pilot Randomized Controlled Trial of Goal Management Training in Canadian Military Members, Veterans, and Public Safety Personnel Experiencing Post-Traumatic Stress Symptoms | Not CCT |
| Rahman 2017 | BDNF Val66Met and childhood adversity on response to physical exercise and internet-based cognitive behavioural therapy in depressed Swedish adults | Not CCT |
| Rajji 2015 | Preventing cognitive decline in older patients with depression using cognitive remediation and transcranial direct current stimulation | Data also elsewhere |
| Rajji 2020 | Design and Rationale of the PACt-MD Randomized Clinical Trial: Prevention of Alzheimer's dementia with Cognitive remediation plus transcranial direct current stimulation in Mild cognitive impairment and Depression | Inappropriate control |
| Ramio 2010 | Efficacy of a cognitive rehabilitation programme: "EM line! project" | Not adults with dep. |
| Redondo 2004 | Long-Term Efficacy of Therapy in Patients With Fibromyalgia: A Physical Exercise-Based Program and a Cognitive-Behavioral Approach | Not CCT |
| Reitano 2021 | Effectiveness of a novel cognitive rehabilitation program based on mindfulness and reminiscence in patients with Parkinson's disease and mild cognitive impairment: A pilot study | Not CCT |
| RequenaHernandez 2008 | The effect of motor activity on improved memory and emotional well-being in elderly women | Not CCT |
| Reynolds 2021 | Mindfulness and Cognitive Training Interventions in Mild Cognitive Impairment: Impact on Cognition and Mood. | Not RCT |
| Ridsdale 2012 | The effect of counselling, graded exercise and usual care for people with chronic fatigue in primary care: A randomized trial | Not adults with dep. |
| Rigaud 2008 | Mental compensation strategies and gerontechnology | Not RCT |
| Roberts 2021 | Working memory updating training reduces state repetitive negative thinking: Proof-of-concept for a novel cognitive control training | Not adults with dep. |
| Roheger 2020 | Predictors of changes after reasoning training in healthy adults | Not adults with dep. |
| Roheger 2020 | Lower cognitive baseline scores predict cognitive training success after 6 months in healthy older adults: Results of an online RCT | Not adults with dep. |
| Romanopoulou 2021 | Technology Enhanced Health and Social Care for Vulnerable People During the COVID-19 Outbreak | Not RCT |
| Ronold 2022 | Computerized Working Memory Training in Remission From Major Depressive Disorder: Effects on Emotional Working Memory, Processing Speed, Executive Functions, and Associations With Symptoms | Not RCT |
| Rosenberg 2010 | Exergames for subsyndromal depression in older adults: A pilot study of a novel intervention | Not CCT |
| Routledge 2021 | The impact of online brain training exercises on experiences of depression, anxiety and emotional wellbeing in a twin sample. | Not adults with dep. |
| Rozzini 2007 | Efficacy of cognitive rehabilitation in patients with mild cognitive impairment treated with cholinesterase inhibitors | Not adults with dep. |
| Ruehlman 2021 | A pilot test of Internet-delivered brief interactive training sessions for depression: Evaluating dropout, uptake, adherence, and outcome | Not CCT |
| Rushia 2020 | Testing the Mechanism of Action of Computerized Cognitive Training in Young Adults with Depression: Protocol for a Blinded, Randomized, Controlled Treatment Trial | Not RCT |
| Rutherford 2022 | Working memory in depression | Not RCT |
| SadatZia 2021 | A brief clinical report documenting a novel therapeutic technique (MEmory Specificity Training, MEST) for depression: a summary of two pilot randomized controlled trials | Not CCT |
| Sakakibara 2022 | Telehealth coaching to improve self-management for secondary prevention after stroke: A randomized controlled trial of Stroke Coach | Not CCT |
| Sanchez-Luengos 2021 | Effectiveness of Cognitive Rehabilitation in Parkinson's Disease: A Systematic Review and Meta-Analysis | Not RCT |
| Sangi 2021 | Design and effectiveness of an educational package based on increased cognitive, emotional, and neuromuscular activity in depression in the elderly with mild cognitive impairment Kahrizak Nursing Home for the elderly and disabled in Tehran | Not RCT |
| Santos 2021 | Memory Support System in Spanish: A Pilot Study | Not CCT |
| SargeniusLandahl 2021 | Comparison of attention process training and activity-based attention training after acquired brain injury: A randomized controlled study | Not adults with dep. |
| Schirda 2020 | Mindfulness training for emotion dysregulation in multiple sclerosis: A pilot randomized controlled trial | Not adults with dep. |
| Schmadeke 2015 | Effects of Smartphone-supported Rehabilitation Aftercare (eATROS) for Depressive Patients | Not CCT |
| Schmidt 2021 | Memory enhancement by multidomain group cognitive training in patients with Parkinson's disease and mild cognitive impairment: long-term effects of a multicenter randomized controlled trial | Not CCT |
| Schneider 2019 | Cognitive remediation therapy modulates intrinsic neural activity in patients with major depression | Data also elsewhere |
| Schoene 2015 | Interactive cognitive-motor step training improves cognitive risk factors of falling in older adults-A randomized controlled trial. | Not adults with dep. |
| Schuster 2017 | Exploring blended group interventions for depression: Randomised controlled feasibility study of a blended computer- and multimedia-supported psychoeducational group intervention for adults with depressive symptoms | Not CCT |
| Scogin 1985 | Memory-skills training, memory complaints, and depression in older adults | Not CCT |
| Scogin 2014 | Cognitive bibliotherapy and memory training for older adults with depressive symptoms | Not CCT |
| Segrave 2014 | Concurrent cognitive control training augments the antidepressant efficacy of tDCS: a pilot study | Not CCT |
| Segrave 2015 | Retraining the brain to beat depression: tDCS and cognitive control training | Data also elsewhere |
| Semkovska 2012 | Neurocognitive remediation for depression: A feasibility study | Not RCT |
| Semkovska 2015 | Efficacy of neurocognitive remediation therapy during an acute depressive episode and following remission: Results from two randomised pilot studies | Not RCT |
| Serrat 2021 | Effectiveness of a Multicomponent Treatment Based on Pain Neuroscience Education, Therapeutic Exercise, Cognitive Behavioral Therapy, and Mindfulness in Patients With Fibromyalgia (FIBROWALK Study): A Randomized Controlled Trial | Not CCT |
| Seshadri 2020 | Feasibility Study of Stress Management and Resiliency Training (SMART) in Patients With Major Depressive Disorder | Not CCT |
| Sharma 2021 | Enhancing memory and quality of life through novel home-based neuropsychological rehabilitation for epilepsy: A randomized controlled trial | Not CCT |
| Shelly 2022 | Clinical Outcomes of a Brief Group CBT as a Stepped Care Approach to Cancer-Related Cognitive Impairment | Not CCT |
| Shahpouri 2020 | Comparison of Cognitive Rehabilitation versus Donepezil Therapy on Memory Performance, Attention, Quality of Life, and Depression among Multiple Sclerosis Patients | Not CCT |
| Sheykhan 2013 | Self-focused attention in treatment of social anxiety: A controlled clinical trial | Not adults with dep. |
| Shiasy 2020 | The Effectiveness of Attention Bias Modification with and without Trans Cranial Direct Current Stimulation in Chronic Low Back Pain. | Not adults with dep. |
| Shin 2020 | Effects of Process-Based Cognitive Training on Memory in the Healthy Elderly and Patients with Mild Cognitive Impairment: A Randomized Controlled Trial | Not adults with dep. |
| Shu 2022 | Targeting disrupted rich-club network organization with neuroplasticity-based computerized cognitive remediation in major depressive disorder patients | Not RCT |
| Siegle 2014 | You gotta work at it: Pupillary indices of task focus are prognostic for response to a neurocognitive intervention for rumination in depression | Not CCT |
| Silva 2021 | A Home-Based Individual Cognitive Stimulation Program for Older Adults With Cognitive Impairment: A Randomized Controlled Trial | Not CCT |
| Simblett 2017 | Computerized Cognitive Behavioral Therapy to Treat Emotional Distress After Stroke: A Feasibility Randomized Controlled Trial | Not adults with dep. |
| Simonetto 2018 | Gender disparities in cognitive, motor and mood outcomes: Preliminary data from the bugher foundation's combined aerobic and resistance exercise training (CARET) program and cognitive training intervention (CTI) study | Not adults with dep. |
| Sit 2022 | The effect of positive mental imagery training on Chinese university students with depression: A pilot study | Not CCT |
| Slyunkova 2021 | Effectiveness of biofeedback training based on the brain-computer interface in poststroke patients | Not CCT |
| Smith 2009 | A cognitive training program based on principles of brain plasticity: Results from the improvement in memory with plasticity-based adaptive cognitive training (IMPACT) study | Not adults with dep. |
| Smith 2018 | Computerized Cognitive Training to Improve Mood in Senior Living Settings: Design of a Randomized Controlled Trial | Not adults with dep. |
| Smith 2019 | Speed of processing training and depression in assisted and independent living: A randomized controlled trial | Not adults with dep. |
| Smolarczyk-Kosowska 2021 | Assessment of the Impact of a Daily Rehabilitation Program on Anxiety and Depression Symptoms and the Quality of Life of People with Mental Disorders during the COVID-19 Pandemic | Not CCT |
| Smucny 2020 | Can pharmacological augmentation of cognitive training remediate age-related cognitive decline? | Inappropriate control |
| So 2021 | A randomised controlled trial of metacognitive training for psychosis, depression, and belief flexibility | Not CCT |
| Sociali 2022 | What role for cognitive remediation in the treatment of depressive symptoms? A superiority and noninferiority meta-analysis for clinicians | Not RCT |
| Sok 2021 | Effects of Cognitive/Exercise Dual-Task Program on the Cognitive Function, Health Status, Depression, and Life Satisfaction of the Elderly Living in the Community | Not CCT |
| Sommer 2021 | Depression treatment by tDCS-enhanced cognitive control training: A test of two stimulation intensities | Ineligible outcomes |
| Soumet-Leman 2015 | Use of digital and therapeutic relationship in telemedicine: The example of computer assisted cognitive remediation applied to depression | Inappropriate control |
| Soumet-Leman 2016 | Metacognition and cognitive remediation in the treatment of depression | Not RCT |
| Sprague 2021 | Cognitive Training Attenuates Decline in Physical Function Across 10 Years | Ineligible outcomes |
| Srisuwan 2020 | Effects of a Group-Based 8-Week Multicomponent Cognitive Training on Cognition, Mood and Activities of Daily Living among Healthy Older Adults: A One-Year Follow-Up of a Randomized Controlled Trial | Not CCT |
| Stenlund 2009 | Cognitively oriented behavioral rehabilitation in combination with Qigong for patients on long-term sick leave because of burnout: REST-A randomized clinical trial | Not CCT |
| Streltzov 2022 | Effectiveness of a Self-Management Program to Improve Cognition and Quality of Life in Epilepsy: A Pragmatic, Randomized, Multicenter Trial | Not CCT |
| Stuerz 2022 | Effects of a Two-Step Cognitive and Relaxation Training Program in Care Home Residents with Mild Cognitive Impairment | Not CCT |
| Sturz 2011 | [Computer assisted cognitive training advances mood and psychological wellbeing - a comparison to paper pencil training relating to neuropsychological parameters, mood and cognitions] | Not adults with dep. |
| Sturz 2015 | Influence of a relaxation program, cognitive training and a combination of both intervention forms on neuropsychological and affective parameters in elderly care home residents | Not adults with dep. |
| Suddell 2021 | Emotional bias training as a treatment for anxiety and depression: evidence from experimental medicine studies in healthy and medicated samples | Not CCT |
| Sukontapol 2021 | "The effectiveness of a cognitive training program in people with mild cognitive impairment: A study in urban community": Erratum | Not adults with dep. |
| Suojanen 2012 | Neuroplasticity-based computerized cognitive training, depressive symptoms, and functional outcome in schizophrenia | Not adults with dep. |
| SvaerkeSchioler 2022 | Computer-Assisted Self-Training to Improve Executive Function Versus Unspecific Training in Patients after Stroke, Cardiac Arrest or in Parkinson's Disease: A Randomized Controlled Trial: The Compex-Trial | Not RCT |
| Szesny 2011 | Effectiveness of a standardised cognitive training in depression and the interaction with HPA-axis regulation | Not RCT |
| Tan 2021 | A method of VR-EEG scene cognitive rehabilitation training | Not RCT |
| Taylor 2016 | Computerized Cognitive Remediation for Geriatric Depression: Dawn of a New Treatment Modality? | Not RCT |
| TebartzvanElst 2021 | FASTER and SCOTT&EVA trainings for adults with high-functioning autism spectrum disorder (ASD): study protocol for a randomized controlled trial | Not CCT |
| Thams 2022 | Feasibility of Cognitive Training in Combination With Transcranial Direct Current Stimulation in a Home-Based Context (TrainStim-Home): study protocol for a randomised controlled trial | Inappropriate control |
| Therond 2021 | The Efficacy of Cognitive Remediation in Depression: A Systematic Literature Review and Meta-Analysis | Not RCT |
| Thomas 2017 | Age as a moderator of change following compensatory cognitive training in individuals with severe mental illnesses | Not CCT |
| Tiersky 2005 | A trial of neuropsychologic rehabilitation in mild-spectrum traumatic brain injury | Not CCT |
| Timm 2018 | Mindfulness-Based Attention Training Improves Cognitive and Affective Processes in Daily Life in Remitted Patients with Recurrent Depression: A Randomized Controlled Trial | Not CCT |
| Tlach 2011 | Long-term effects of a cognitive-behavioral training program for the management of depressive symptoms among patients in orthopedic inpatient rehabilitation of chronic low back pain: a 2-year follow-up | Not CCT |
| Tonga 2021 | Managing depressive symptoms in people with mild cognitive impairment and mild dementia with a multicomponent psychotherapy intervention: a randomized controlled trial | Not CCT |
| Topper 2017 | Prevention of anxiety disorders and depression by targeting excessive worry and rumination in adolescents and young adults: A randomized controlled trial | Not CCT |
| Trapp 2022 | Cognitive Remediation in Psychiatric Disorders: State of the Evidence, Future Perspectives, and Some Bold Ideas | Not RCT |
| Trombini-Souza 2020 | Dual-task training with progression from variable- to fixed-priority instructions versus dual-task training with variable-priority on gait speed in community-dwelling older adults: A protocol for a randomized controlled trial : Variable- and fixed-priorit | Not RCT |
| Tsumura 2012 | The effects of attention retraining on depressive mood and cortisol responses to depression-related stimuli | Not CCT |
| Tully 2015 | A dynamic view of comorbid depression and generalized anxiety disorder symptom change in chronic heart failure: The discrete effects of cognitive behavioral therapy, exercise, and psychotropic medication | Not CCT |
| Twamley 2019 | Compensatory cognitive training for people with severe mental illnesses in supported employment: A randomized controlled trial | Not CCT |
| Tyrrell 2019 | Cognitive Self-Efficacy and Mental Health Ratings after a Memory Skills Group for Older Veterans with PTSD | Not CCT |
| Ullmann 2021 | Study protocol of a randomized intervention study to explore effects of a pure physical training and a mind-body exercise on cognitive executive function in independent living adults age 65-85 | Not CCT |
| Ulrichsen 2022 | No add-on effect of tDCS on fatigue and depression in chronic stroke patients: A randomized sham-controlled trial combining tDCS with computerized cognitive training | Ineligible outcomes |
| Vaia 2022 | Computer-aided cognitive training in patients with neurocognitive vascular impairment: effects on cognition, depression and behavior | Not RCT |
| Vaishnavi 2020 | Cognitive Training During TMS Does Not Improve Depression Outcomes | Not RCT |
| Vajawat 2021 | Digital Gaming Interventions in Psychiatry: Evidence, Applications and Challenges | Not RCT |
| vanBeers 2021 | Working memory training efficacy in COPD: the randomised, double-blind, placebo-controlled Cogtrain trial | Not adults with dep. |
| VanBreda 2021 | Does cognitive behavioral therapy improve psychosocial outcome in rheumatoid arthritis: A systematic literature review | Not CCT |
| VanSchooten 2021 | Protocol of a 12-month multifactorial eHealth programme targeting balance, dual-tasking and mood to prevent falls in older people: The StandingTall + randomised controlled trial | Not CCT |
| Vance 2021 | Targeting HIV-Related Neurocognitive Impairments with Cognitive Training Strategies: Insights from the Cognitive Aging Literature | Not RCT |
| VandeVelde 2020 | Cognitive remediation following electroconvulsive therapy in patients with treatment resistant depression: randomized controlled trail of an intervention for relapse prevention - study protocol | Not RCT |
| VandenBergh 2018 | Remediation of depression-related cognitive impairment: cognitive control training as treatment augmentation | Not RCT |
| VandenBergh 2020 | Cognitive Control Training as an Augmentation Strategy to CBT in the Treatment of Fear of Failure in Undergraduates | Not adults with dep. |
| Vanderhasselt 2016 | Emotional reactivity to valence-loaded stimuli are related to treatment response of neurocognitive therapy | Inappropriate control |
| Vanderhasselt 2021 | Cognitive Control Training in Healthy Older Adults: A Proof of Concept Study on the Effects on Cognitive Functioning, Emotion Regulation and Affect | Not adults with dep. |
| VanderZwalmen 2022 | Cognitive remediation for depression vulnerability: Current challenges and new directions | Not RCT |
| VandeVen 2017 | The influence of computer-based cognitive flexibility training on subjective cognitive wellbeing after stroke: A multi-center randomized controlled trial | Not adults with dep. |
| vanEeden 2015 | An economic evaluation of an augmented cognitive behavioural intervention vs. computerized cognitive training for post-stroke depressive symptoms | Not adults with dep. |
| Varalta 2021 | Physiotherapy versus Consecutive Physiotherapy and Cognitive Treatment in People with Parkinson's Disease: A Pilot Randomized Cross-Over Study | Not CCT |
| Verghese 2016 | Cognitive remediation to enhance mobility in older adults: The CREM study | Not adults with dep. |
| Vervaeke 2018 | Gamified Cognitive Control Training for Remitted Depressed Individuals: User Requirements Analysis | Not RCT |
| Vicent-Gil 2019 | Testing the efficacy of INtegral Cognitive REMediation (INCREM) in major depressive disorder: study protocol for a randomized clinical trial | Data not available |
| Vicent-Gil 2022 | Randomized clinical trial of integral cognitive remediation program for major depression (INCREM) | Not CCT |
| Vieira 2018 | Virtual reality exercise on a home-based phase III cardiac rehabilitation program, effect on executive function, quality of life and depression, anxiety and stress: a randomized controlled trial | Not adults with dep. |
| VillasanRueda 2021 | Improvement of the Quality of Life in Aging by Stimulating Autobiographical Memory | Not CCT |
| Visser 2020 | A Pilot Study of Smartphone-Based Memory Bias Modification and Its Effect on Memory Bias and Depressive symptoms in an Unselected Population | Not adults with dep. |
| Vrijsen 2018 | Cognitive bias modification as an add-on treatment in clinical depression: Results from a placebo-controlled, single-blinded randomized control trial | Not CCT |
| Wang 2018 | The Effects of mindfulness-based cognitive therapy (MBCT) on anxiety and depression among professional women: Increased EEG gamma and alpha brainwave amplitude | Not CCT |
| Wang 2022 | The antidepressant effect of cognitive reappraisal training on individuals cognitively vulnerable to depression: Could cognitive bias be modified through the prefrontal-amygdala circuits? | Not CCT |
| Waters 2016 | A preliminary evaluation of a home-based, computer-delivered attention training treatment for anxious children living in regional communities. | Not adults with dep. |
| Watkins 2009 | Concreteness training reduces dysphoria: A pilot proof-of-principle study | Not CCT |
| Webb 2021 | Cognition-oriented treatments for older adults: A systematic review of the influence of depression and self-efficacy individual differences factors | Not RCT |
| Welch 2019 | Feasibility of Computerized Cognitive-Behavioral Therapy Combined With Bifrontal Transcranial Direct Current Stimulation for Treatment of Major Depression | Not CCT |
| Wells 2010 | Biased attention and dysphoria: Manipulating selective attention reduces subsequent depressive symptoms | Not CCT |
| Werner-Seidler 2018 | A cluster randomized controlled platform trial comparing group MEmory specificity training (MEST) to group psychoeducation and supportive counselling (PSC) in the treatment of recurrent depression | Not CCT |
| White 2007 | Protocol for the PACE trial: A randomised controlled trial of adaptive pacing, cognitive behaviour therapy, and graded exercise as supplements to standardised specialist medical care versus standardised specialist medical care alone for patients with the chronic fatigue syndrome/myalgic encephalomyelitis or encephalopathy | Not CCT |
| Wilhelm 2022 | Efficacy of App-Based Cognitive Behavioral Therapy for Body Dysmorphic Disorder with Coach Support: Initial Randomized Controlled Clinical Trial | Not CCT |
| Winningham 2003 | MemAerobics: A Cognitive Intervention to Improve Memory Ability and Reduce Depression in Older Adults | Not RCT |
| Winningham 2007 | A cognitive intervention to enhance institutionalized older adults' social support networks and decrease loneliness | Not RCT |
| Wolinsky 2009 | The effect of speed-of-processing training on depressive symptoms in ACTIVE | Not adults with dep. |
| Wolinsky 2009 | The ACTIVE cognitive training interventions and the onset of and recovery from suspected clinical depression | Not adults with dep. |
| Wolinsky 2015 | The effect of cognitive speed of processing training on the development of additional IADL difficulties and the reduction of depressive symptoms: results from the IHAMS randomized controlled trial | Not adults with dep. |
| Woodford 2021 | Internet-Administered Cognitive Behavioral Therapy for Common Mental Health Difficulties in Parents of Children Treated for Cancer: Intervention Development and Description Study | Not CCT |
| Woolf 2020 | Study protocol for a feasibility RCT of Club Connect, a healthy brain ageing cognitive training program for older adults with depression | Not CCT |
| Woolf 2020 | A systematic review and meta-analysis of cognitive training in adults with major depressive disorder | Not RCT |
| Woolf 2021 | A Systematic Review and Meta-Analysis of Cognitive Training in Adults with Major Depressive Disorder | Not RCT |
| Woolf 2022 | A Systematic Review and Meta-Analysis of Cognitive Training in Adults with Major Depressive Disorder | Not RCT |
| Wright 2021 | Does comprehensive cognitive remediation improve emotion perception? | Not adults with dep. |
| Wuthrich 2018 | Reducing risk factors for cognitive decline through psychological interventions: a pilot randomized controlled trial | Not CCT |
| Wyant 2018 | Feasibility and acceptability of a computerized working memory training in breast cancer survivors | Not adults with dep. |
| Xavier 2011 | Lower incidence of new depressive episodes in elderly with subjective cognitive impairment in a cognitive rehabilitation program | Not adults with dep. |
| Xavier 2013 | The influence of former tobacco exposition in a cognitive stimulation and rehabilitation program, based in computers and internet | Not RCT |
| Xiao 2005 | Cognitive Function of Patients with Schizophrenia: Test of a Neuropsychological Training System | Not adults with dep. |
| Xing 2021 | Investigating the Impact of Cognitive Training for Individuals With Bothersome Tinnitus: A Randomized Controlled Trial | Not adults with dep. |
| Xu 2021 | A method of VR-EEG scene cognitive rehabilitation training | Not CCT |
| Xu 2021 | Comparative effectiveness of non-pharmacological interventions for depressive symptoms in mild cognitive impairment: systematic review with network meta-analysis | Not RCT |
| Yang 2007 | Influence of laudatory training on psychological health in medical college students | Not CCT |
| Yang 2020 | Cognitive and Psychosocial Outcomes of Self-Guided Executive Function Training and Low-Intensity Aerobic Exercise in Healthy Older Adults | Not adults with dep. |
| Yang 2021 | Nurse-led exercise and cognitive-behavioral care against nurse-led usual care between and after chemotherapy cycles in Han Chinese women of ovarian cancer with moderate to severe levels of cancer-related fatigue: A retrospective analysis of the effectiveness | Not CCT |
| Yin 2022 | Alleviated Anxiety Boosts Memory Training Gain in Older Adults with Subjective Memory Complaints: A Randomized Controlled Trial. | Not adults with dep. |
| Yokoi 2021 | Dual-Task Training Combining Cognitive Tasks and Occupations among Japanese Community-Dwelling Older Adults: A Pilot Study | Not RCT |
| Yu 2019 | Effect of a novel designed intensive patient care program on cognitive impairment, anxiety, depression as well as relapse free survival in acute ischemic stroke patients: a randomized controlled study | Not adults with dep. |
| Zahid 2022 | The reliability, validity, and responsiveness of Cognitive Exercise Therapy Approach-Biopsychosocial Questionnaire for patients with fibromyalgia | Not RCT |
| Zamirinejad 2014 | Effectiveness of resilience training versus cognitive therapy on reduction of depression in female Iranian college students | Not CCT |
| Zarit 1981 | Memory training in the community aged: Effects on depression, memory complaint, and memory performance | Not CCT |
| Zeiss 1979 | Nonspecific improvement effects in depression using interpersonal skills training, pleasant activity schedules, or cognitive training | Not CCT |
| Zhang 2000 | Psychotherapy on negative emotions of coronary heart disease and its clinical implications | Not CCT |
| Zhang 2019 | Working memory training can improve anhedonia in college students with subsyndromal depressive symptoms | Not RCT |
| Zhao 2020 | Effects of working memory training on EEG, cognitive performance, and self-report indices potentially relevant for social anxiety | Not adults with dep. |
| Zhou 2022 | Envision A Bright Future to Heal Your Negative Mood: A Trial in China | Not CCT |
| Zhou 2022 | Efficacy of computerized cognitive training on improving cognitive functions of stroke patients: A systematic review and meta-analysis of randomized controlled trials | Not RCT |
| Zhu 2011 | Effect of computer-assisted cognitive training on the cognitive function and depression in patients with brain injury | Not adults with dep. |
| Zhu 2022 | Immersive Virtual Reality-Based Cognitive Intervention for the Improvement of Cognitive Function, Depression, and Perceived Stress in Older Adults With Mild Cognitive Impairment and Mild Dementia: Pilot Pre-Post Study | Not RCT |
| Zhu 2022 | A Multimodal Intervention to Improve Cognition in Community-dwelling Older Adults | Not RCT |
| Zweerings 2020 | Rt-fMRI neurofeedback-guided cognitive reappraisal training modulates amygdala responsivity in posttraumatic stress disorder | Not CCT |

**eTable 6.** Risk of Bias Within Individual Studies

| **Study** | **Comparison** | **Randomization process** | **Deviations from intended intervention** | **Missing outcome data** | **Measurement of the outcome** | **Selection of the reported results** | **Overall risk of bias** |
| --- | --- | --- | --- | --- | --- | --- | --- |
| Amato 2014 | ST vs n-ST | Some concerns | Some concerns | Low | Low | Some concerns | Some concerns |
| Arean 2016 | EVO vs iPST | Low | Some concerns | High | Low | Some concerns | High |
| Arean 2016 | EVO vs HT | Low | Some concerns | High | Low | Some concerns | High |
| Bowie 2013 | CR vs WL | Low | Some concerns | Low | Low | Some concerns | Low |
| Choi 2017 | M-ECT vs AC | Some concerns | High | High | Low | Some concerns | High |
| Choi 2017 | M-ECT vs TAU | High | Some concerns | Low | Low | Some concerns | Low |
| De Luca 2019 | EG vs CG | Some concerns | Low | Low | Some concerns | Some concerns | Some concerns |
| Dos Santos 2020a | CR vs Exercises at Home | Some concerns | High | High | Some concerns | Some concerns | High |
| Dos Santos 2020b | CR vs Phone Call | Some concerns | High | High | Some concerns | Some concerns | High |
| Edwards 2013 | SOPT vs CG | Low | High | High | Some concerns | Low | High |
| Ferrari 2021 | Active-training vs Sham-training | Some concerns | Low | Low | Low | Some concerns | Low |
| Finn 2015 | Training vs Control | Low | High | Low | Some concerns | Some concerns | Some concerns |
| Grasso 2017 | CMD vs MD | Some concerns | Some concerns | Low | Low | Some concerns | Low |
| Hagen 2020 | CCT vs GMT | Low | Some concerns | Low | High | Low | High |
| Hoorelbeke 2017 | CCT vs Control | Low | Some concerns | Low | Low | High | Low |
| Iacoviello 2014 | EMFT vs CT | Some concerns | Some concerns | Low | Low | Some concerns | Low |
| Iacoviello 2018 | EMFT vs CT | Low | High | High | Low | Some concerns | High |
| Kang 2021 | VR Cognitive Training vs Usual Care | Low | Some concerns | Low | Some concerns | Some concerns | Some concerns |
| Klojcnik 2021 | CCRT vs Control | Some concerns | High | Low | Some concerns | High | Some concerns |
| Listunova 2020 | IT vs CG | Some concerns | Some concerns | Low | Low | Some concerns | Low |
| Listunova 2020 | GT vs CG | Some concerns | Some concerns | Low | Low | Some concerns | Low |
| Maggio 2018 | Experimental vs Control | Some concerns | Low | Low | Low | Some concerns | Low |
| Mahnke 2021 | BrainHQ vs Computer games | Low | Low | Low | Low | Some concerns | Low |
| Mattioli 2010 | SG vs CG | Some concerns | Some concerns | High | Low | Some concerns | High |
| Messinis 2017 | RehaCom vs CG | Some concerns | Some concerns | Low | Low | Some concerns | Low |
| Messinis 2020 | RehaCom vs CG | Some concerns | Low | Low | Low | Some concerns | Low |
| Morimoto 2020 | nCCR vs AC | Some concerns | Some concerns | High | Low | Some concerns | High |
| Oh 2018 | SMART vs WL | Some concerns | High | Some concerns | Some concerns | Some concerns | Some concerns |
| Oh 2018 | Fit Brains vs WL | Some concerns | High | Low | Some concerns | Some concerns | Some concerns |
| Roberts 2021 | NEUA vs NEU | Low | Low | High | Low | Some concerns | High |
| Semkovska 2015 | NCRT vs Games | Some concerns | High | High | Low | Some concerns | High |
| Semkovska 2017 | NCRT vs Games | Low | Some concerns | Low | Low | Some concerns | Low |
| Solari 2004 | SG vs CG | Some concerns | Low | Low | Low | Some concerns | Low |
| Trapp 2016 | EG vs CG | Low | Low | Low | Low | Low | Low |
| Trebo 2007 | Training Group vs CG | High | High | High | Some concerns | Some concerns | High |
| Vance 2021 | BrainHQ vs Passive Control | Some concerns | High | High | Some concerns | Low | High |
| Vervake 2021 | aPASAT vs speed-of-response training task | Low | Low | Low | Low | Low | Low |
| Vilou 2020 | Intervention Group vs Control Group | Some concerns | Low | Low | Low | Some concerns | Low |
| Wanmaker 2015 | EG vs PG | Some concerns | High | Some concerns | Low | Some concerns | Some concerns |

**eTable 7.** Sensitivity Analyses – Correlation Assumptions

|  | **Rho = 0** | **Rho = 0.2** | **Rho = 0.4** | **Rho = 0.6** | **Rho = 0.8** | **Rho = 1** |
| --- | --- | --- | --- | --- | --- | --- |
| **Overall cognition** |  |  |  |  |  |  |
| **Hedges’ *g*** | 0.276 | 0.276 | 0.276 | 0.276 | 0.276 | 0.276 |
| **Standard error** | 0.050 | 0.050 | 0.050 | 0.050 | 0.050 | 0.050 |
| τ^2^ | 0.077 | 0.077 | 0.078 | 0.078 | 0.078 | 0.079 |
| **Depressive symptoms** |  |  |  |  |  |  |
| **Hedges’ *g*** | 0.231 | 0.231 | 0.231 | 0.231 | 0.231 | 0.231 |
| **Standard error** | 0.072 | 0.072 | 0.072 | 0.072 | 0.072 | 0.072 |
| τ^2^ | 0.066 | 0.066 | 0.066 | 0.066 | 0.066 | 0.066 |

**eFigure 1.** Funnel Plot of Overall Cognition

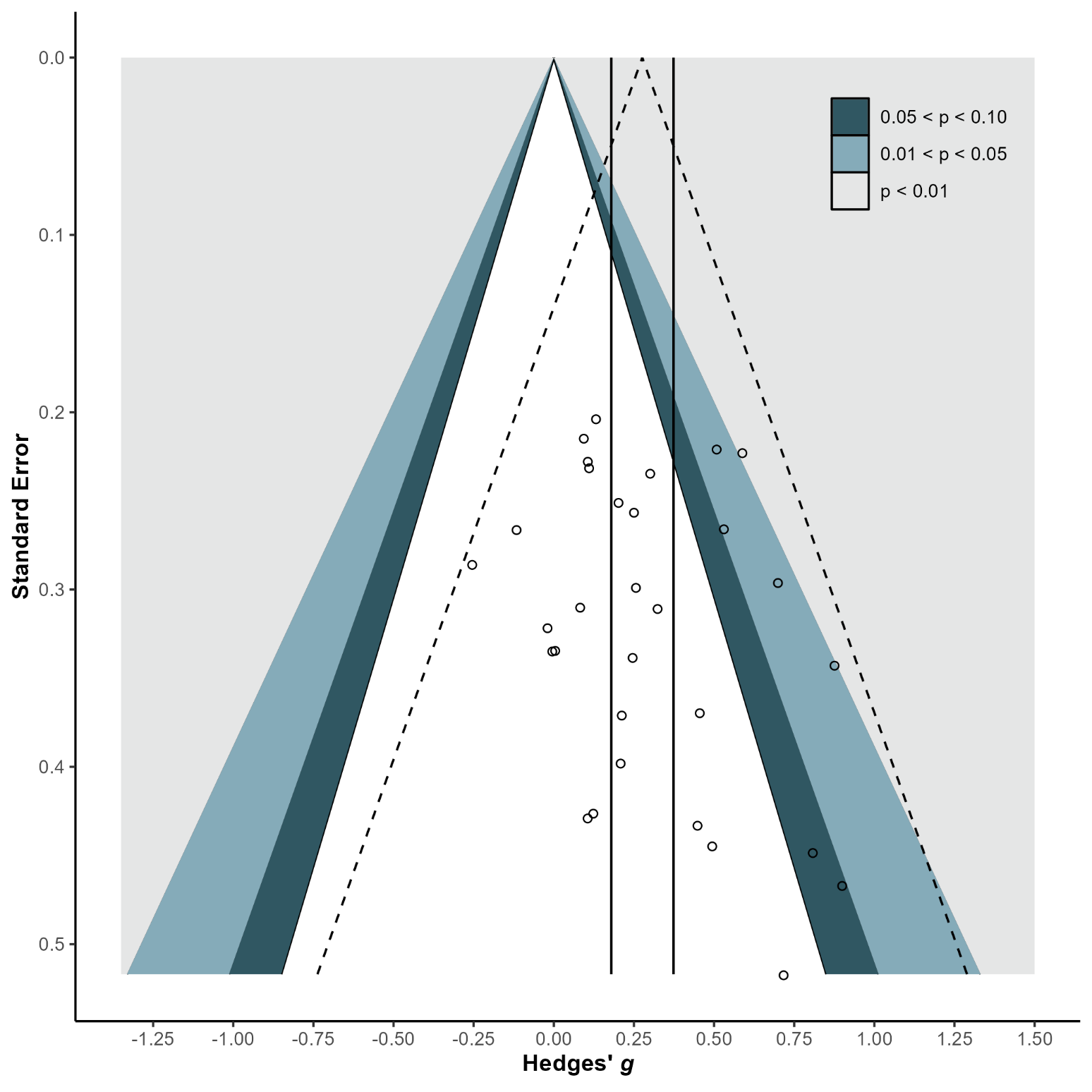

**eFigure 2.** Trim and Fill Funnel Plot of Overall Cognition

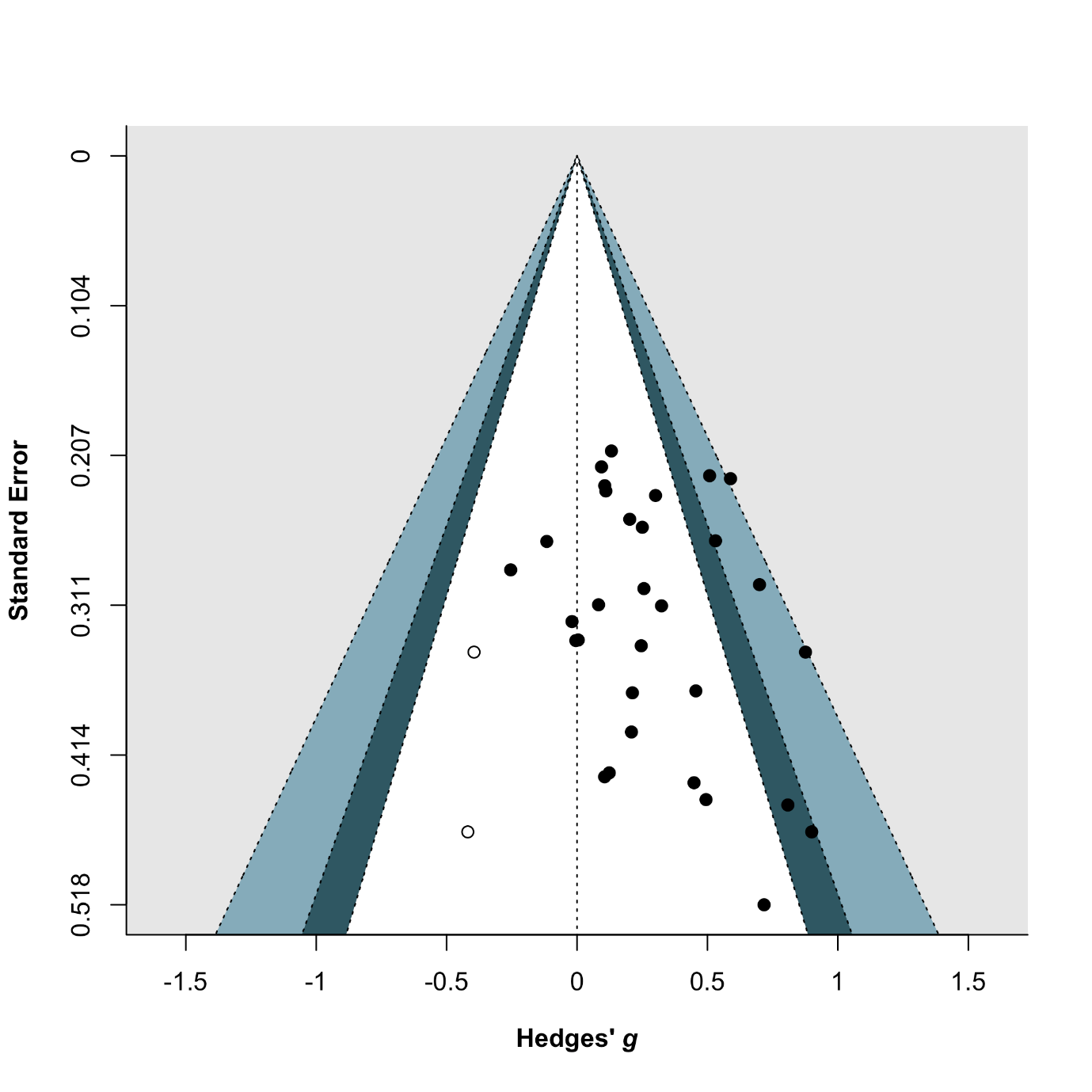

**eFigure 3.** Forest Plot of Depressive Symptoms

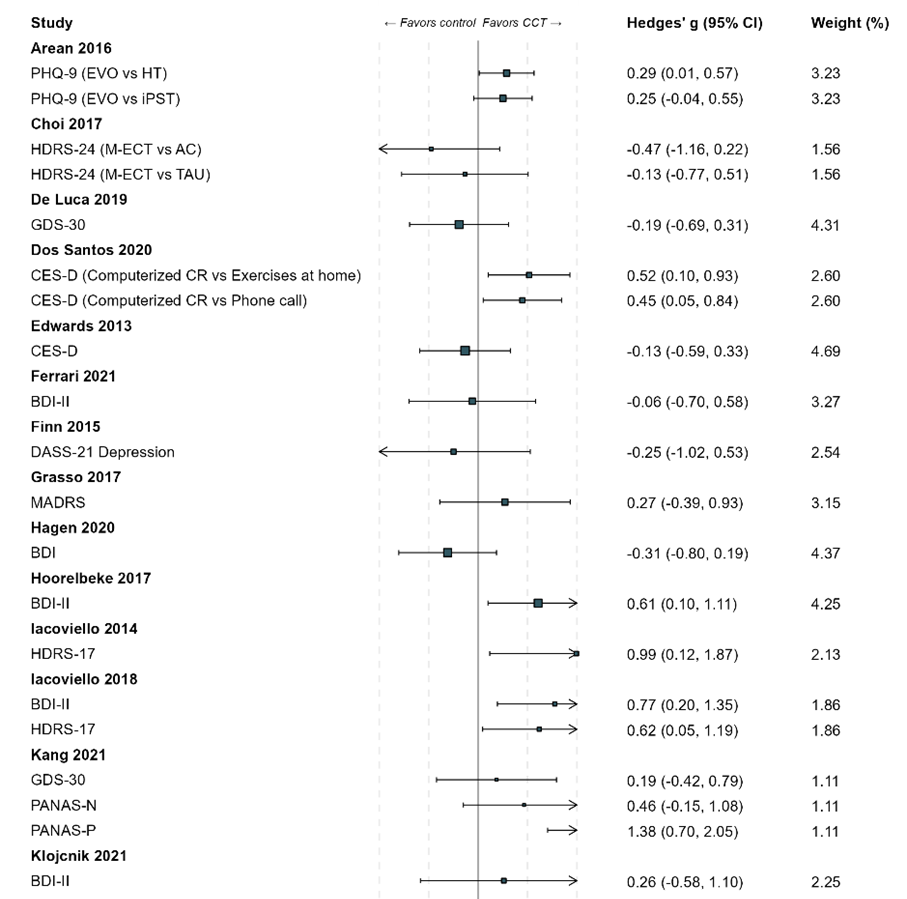

**eFigure 3.** Forest Plot of Depressive Symptoms …continued…

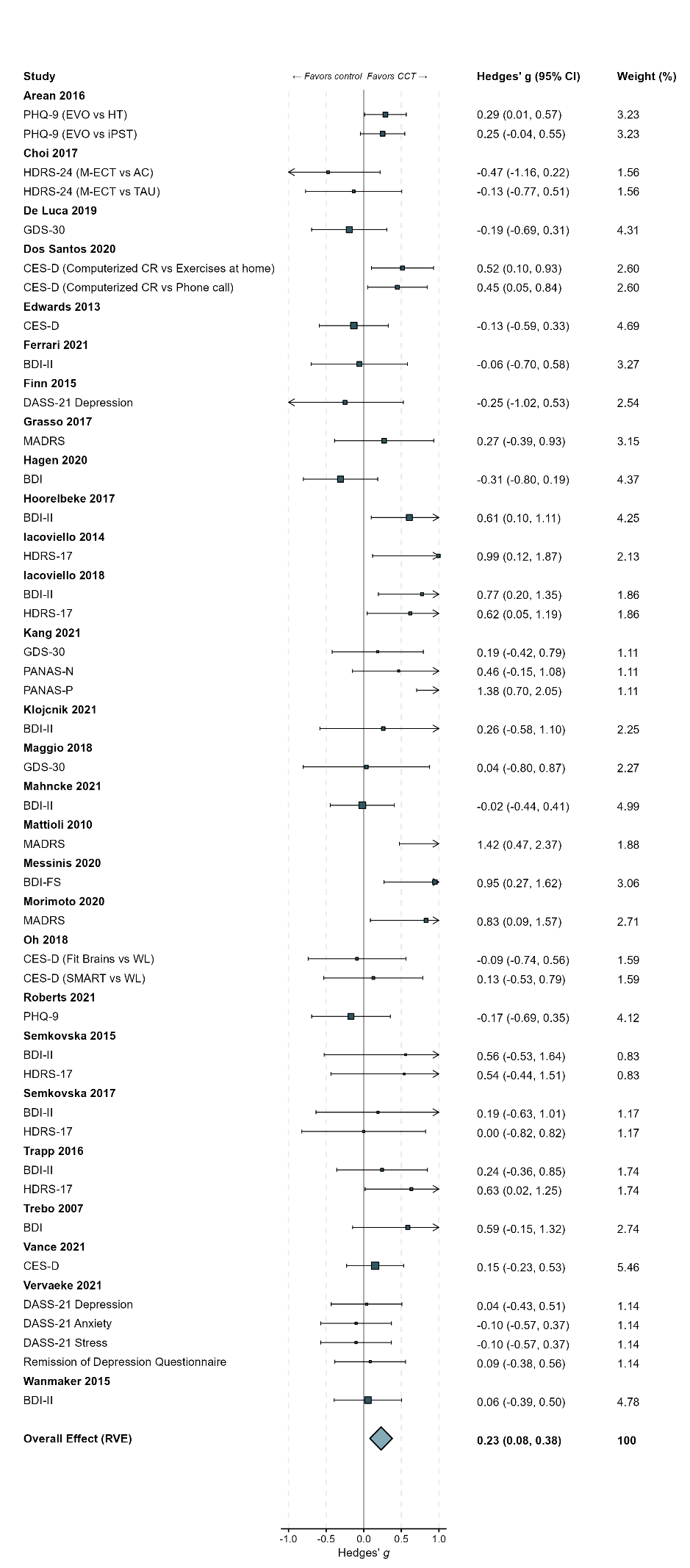

**eFigure 4.** Funnel Plot of Depressive Symptoms

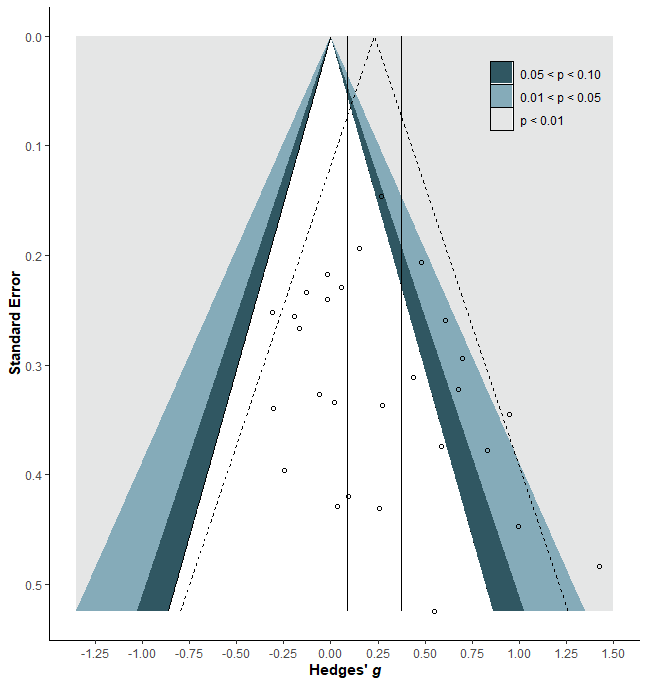

**eFigure5.** Trim and Fill Funnel Plot of Depressive Symptoms

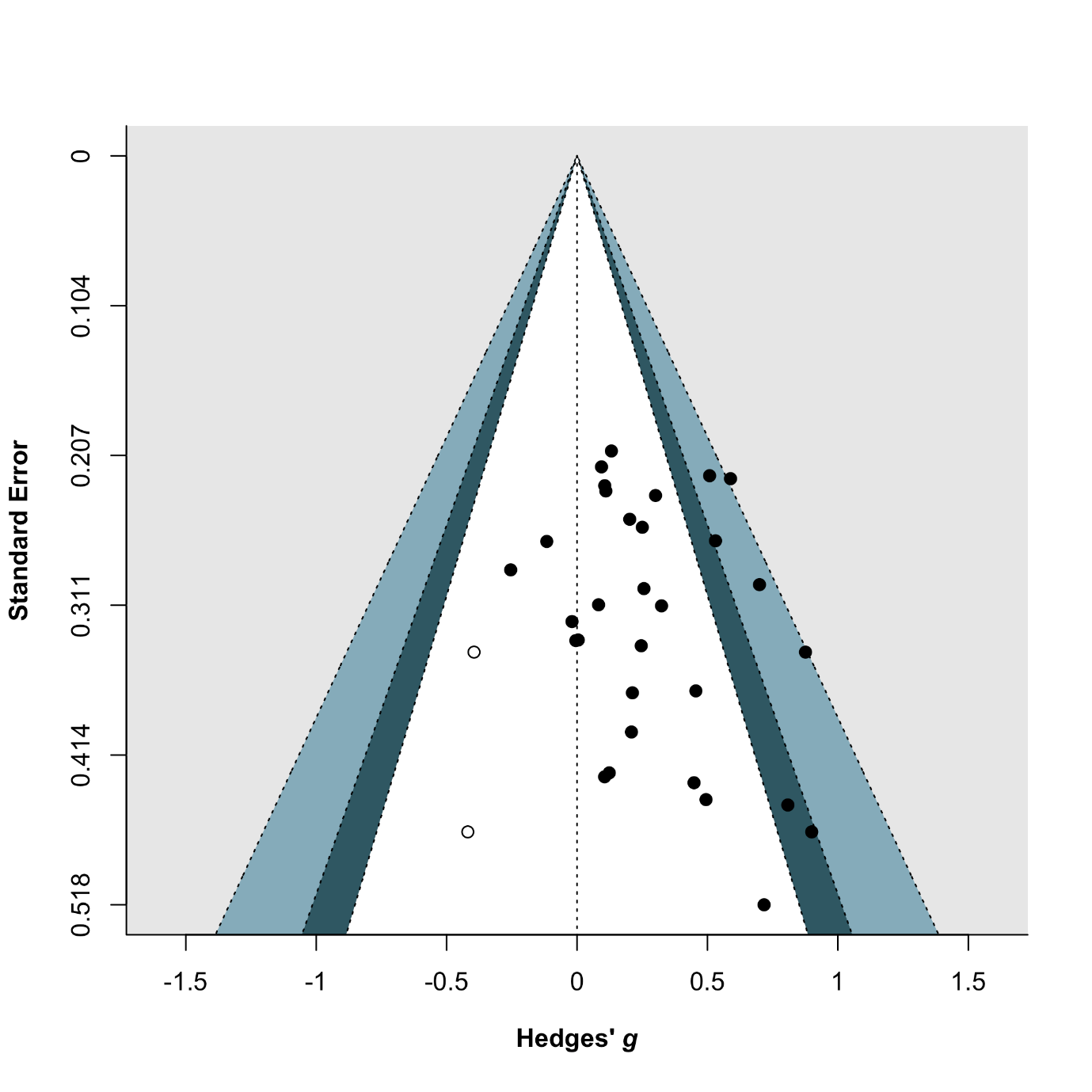

**eFigure 6.** Forest Plot of Psychosocial Functioning

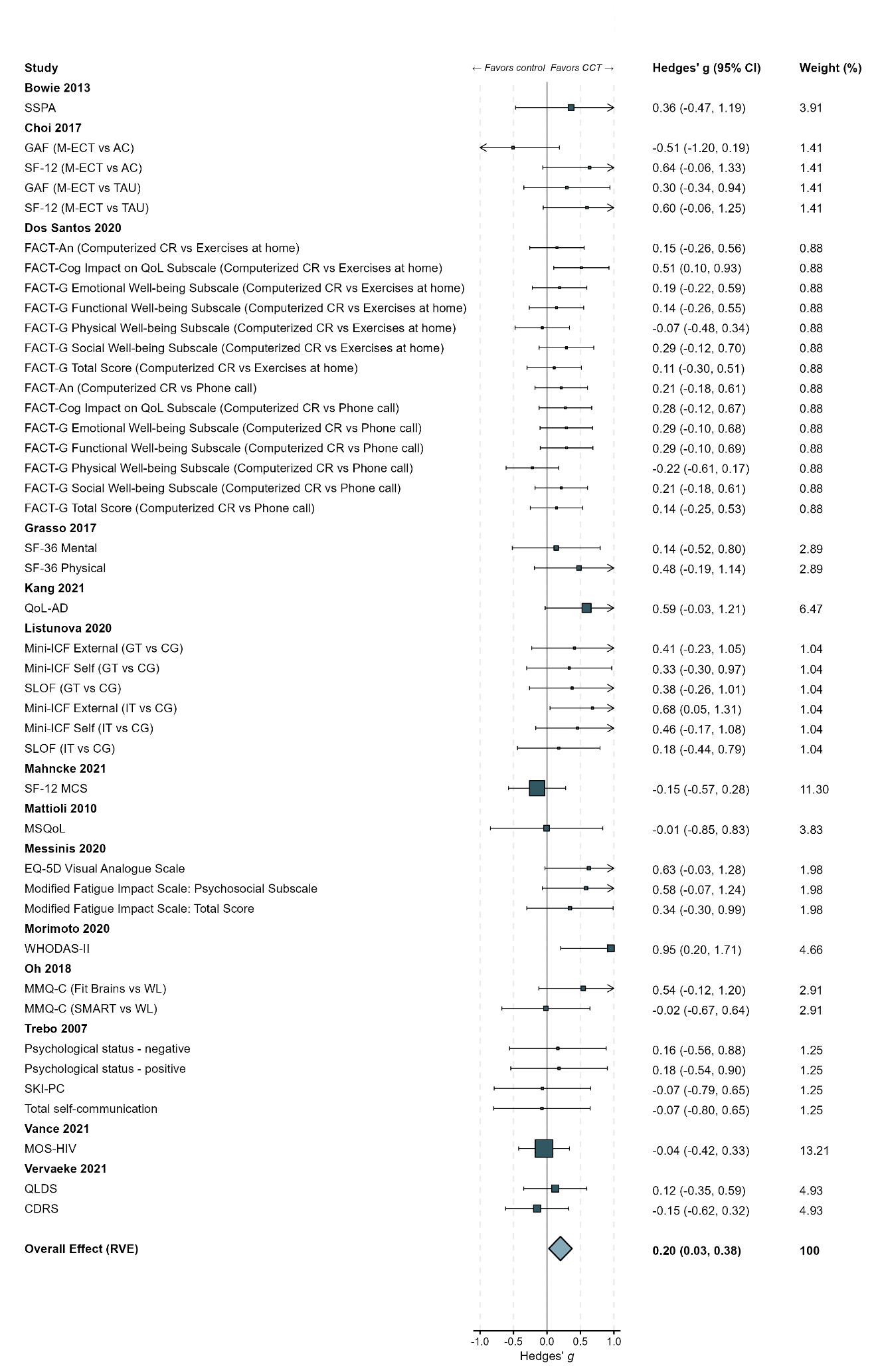

**eFigure 6.** Forest Plot of Psychosocial Functioning..continued…

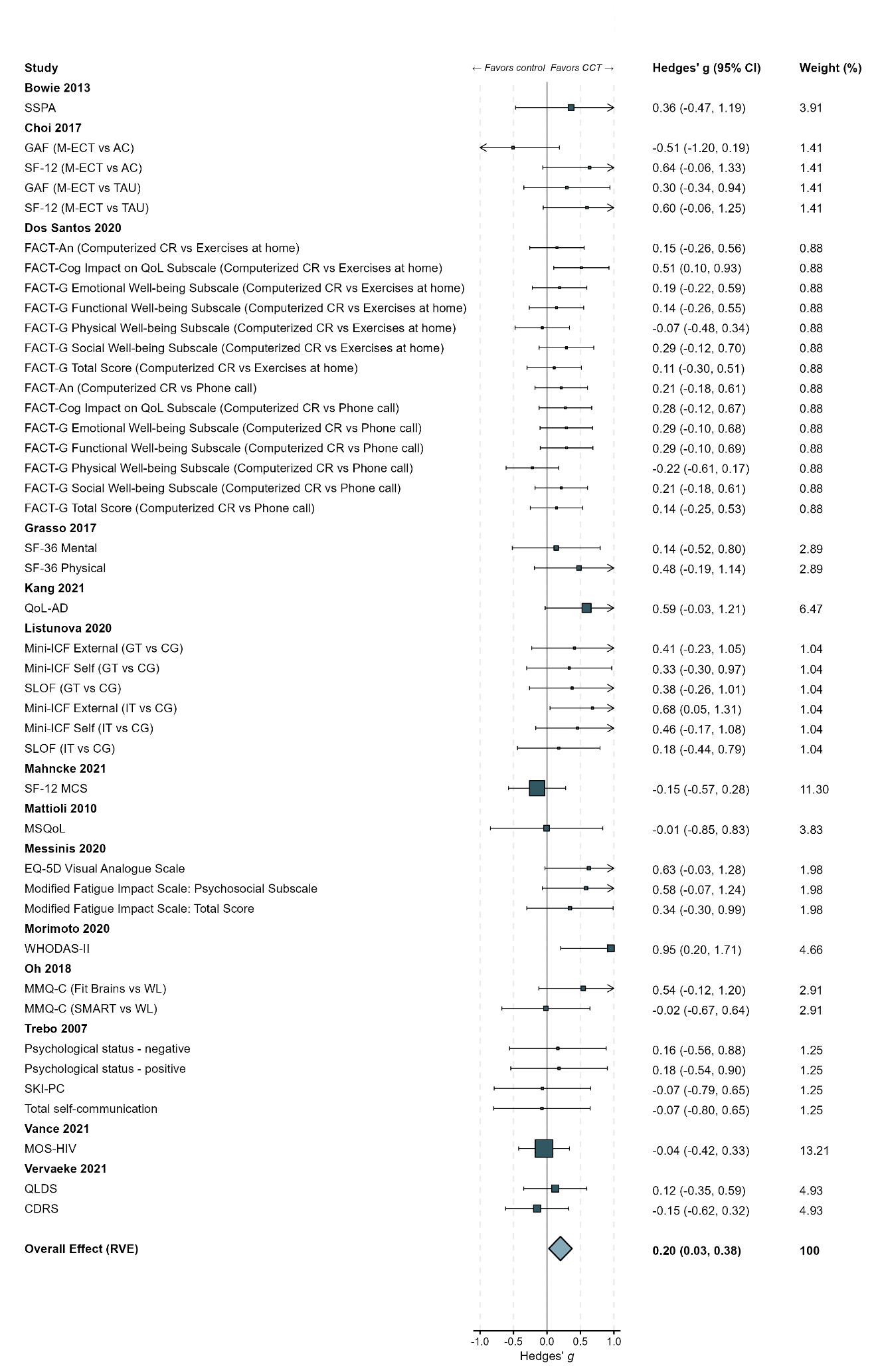

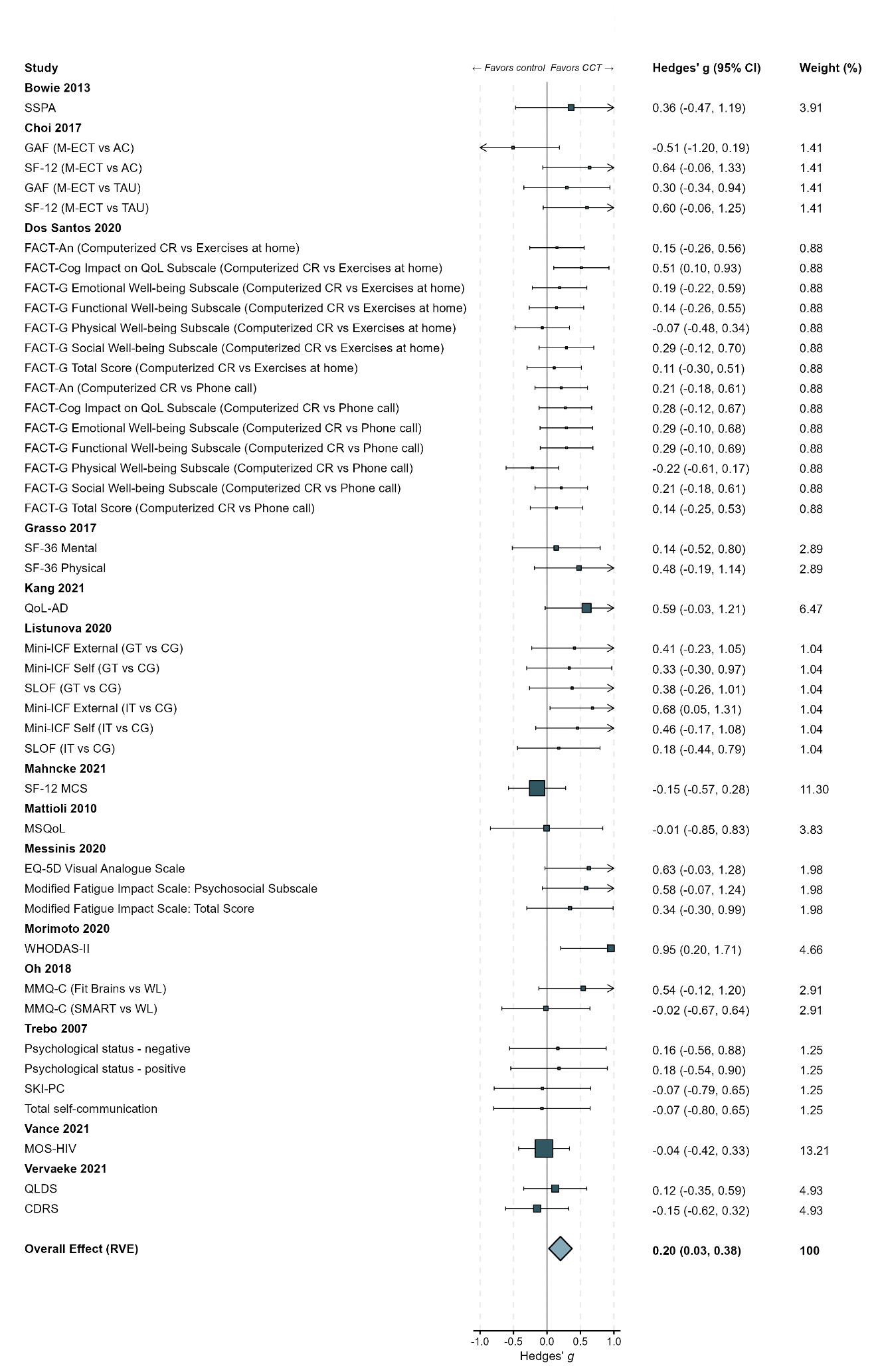

**eFigure 7.** Funnel Plot of Psychosocial Functioning
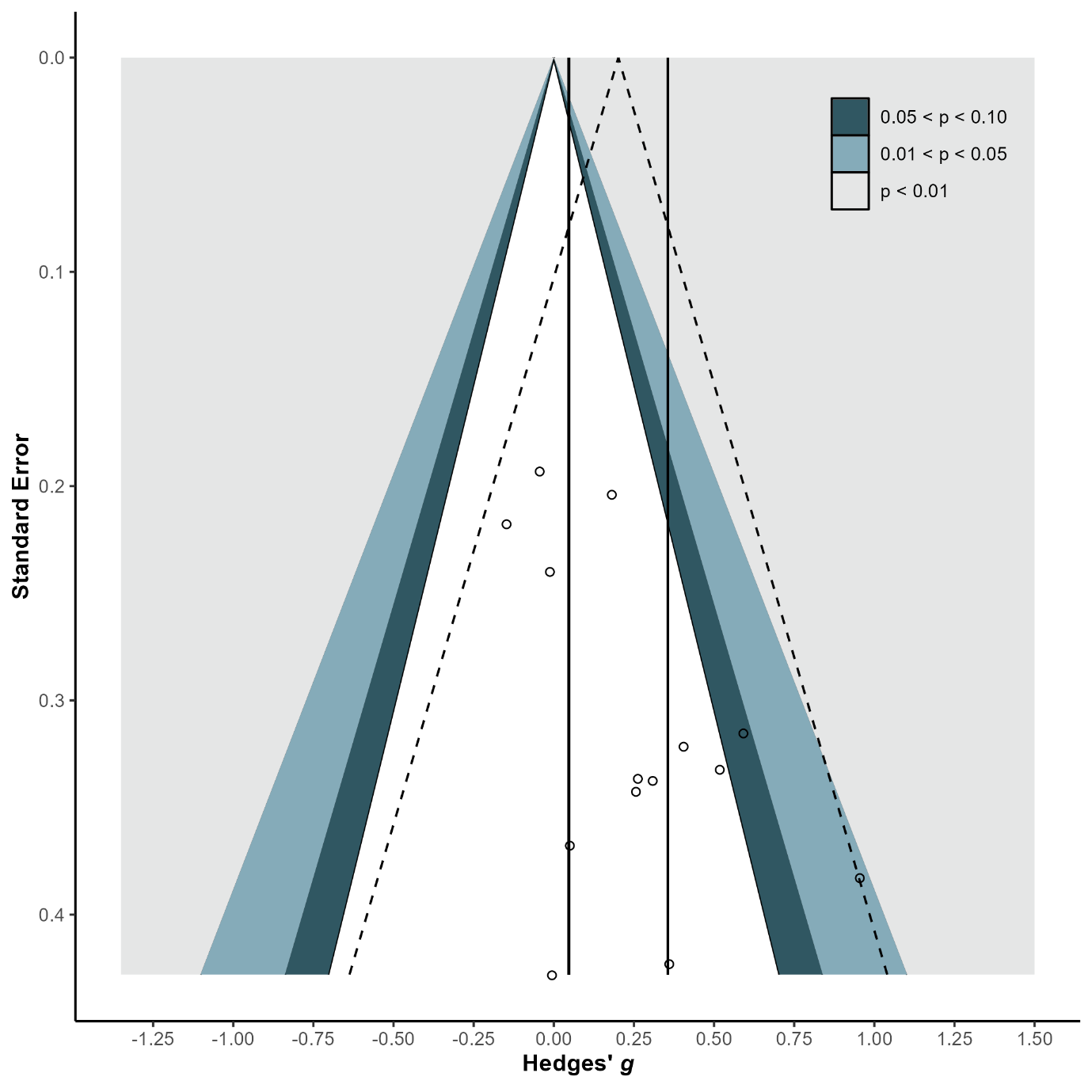

**eFigure 8.** Forest Plot of Psychiatric Symptoms

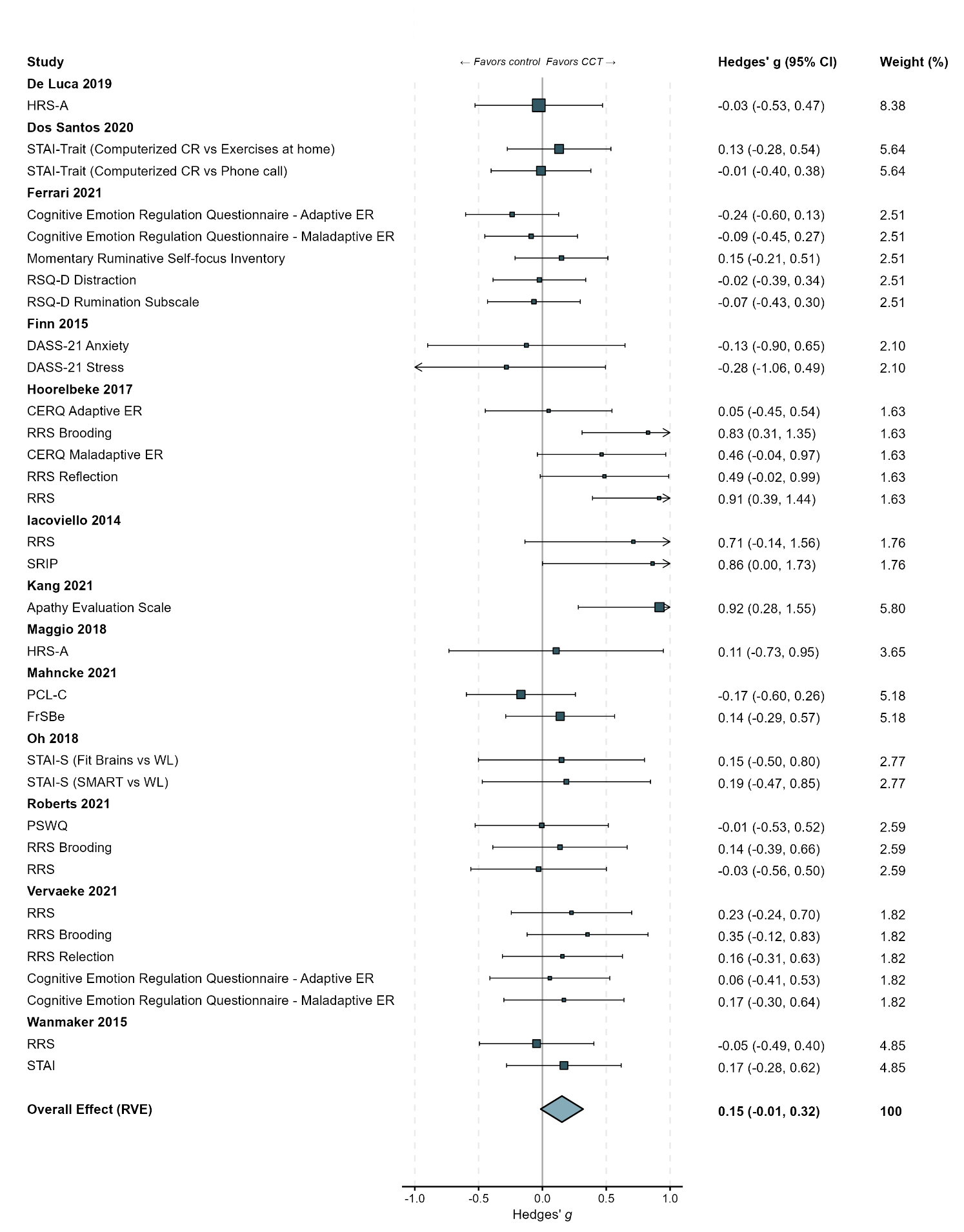

**eFigure 9.** Funnel Plot of Psychiatric Symptoms
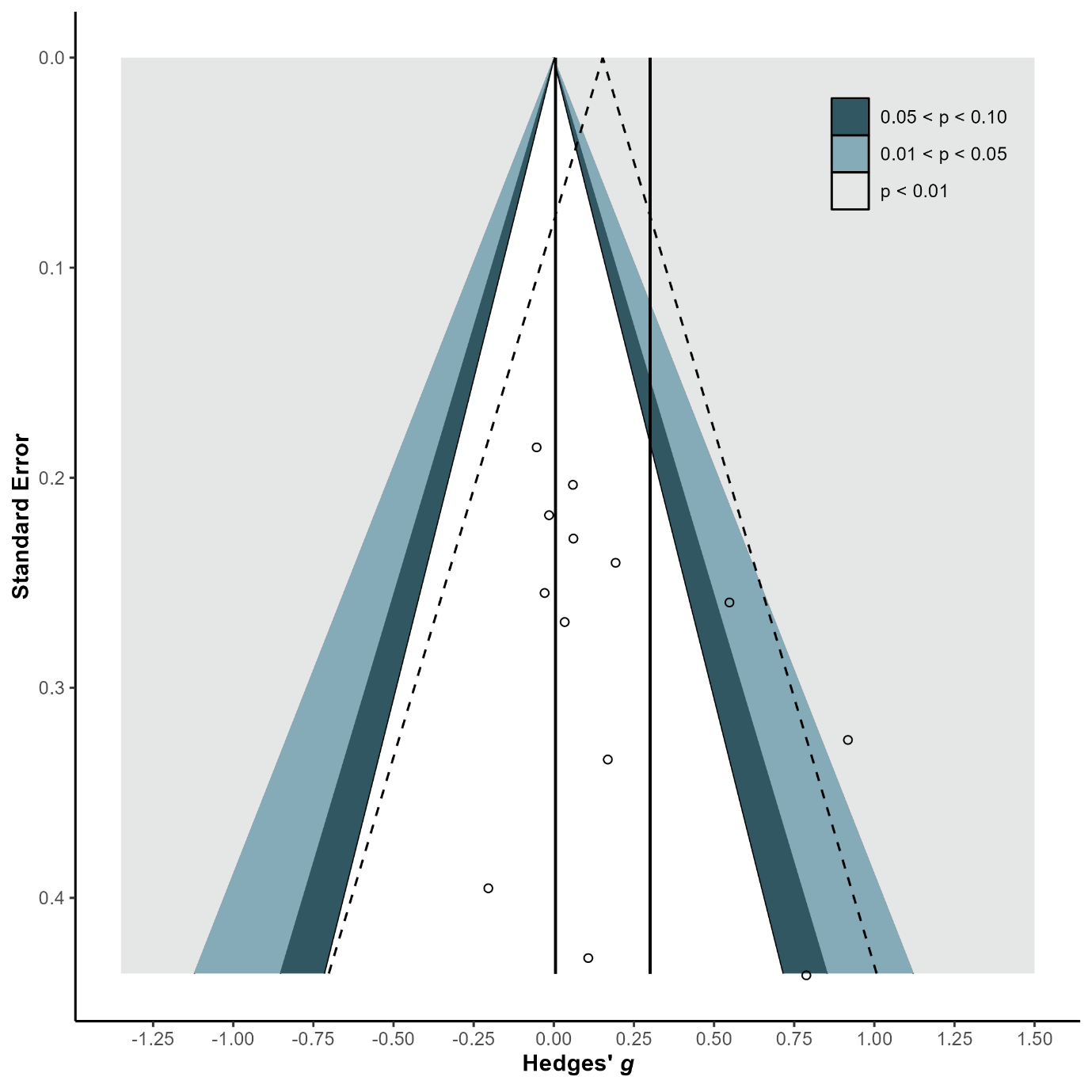

**eFigure 10.** Forest Plot of Subjective Cognition

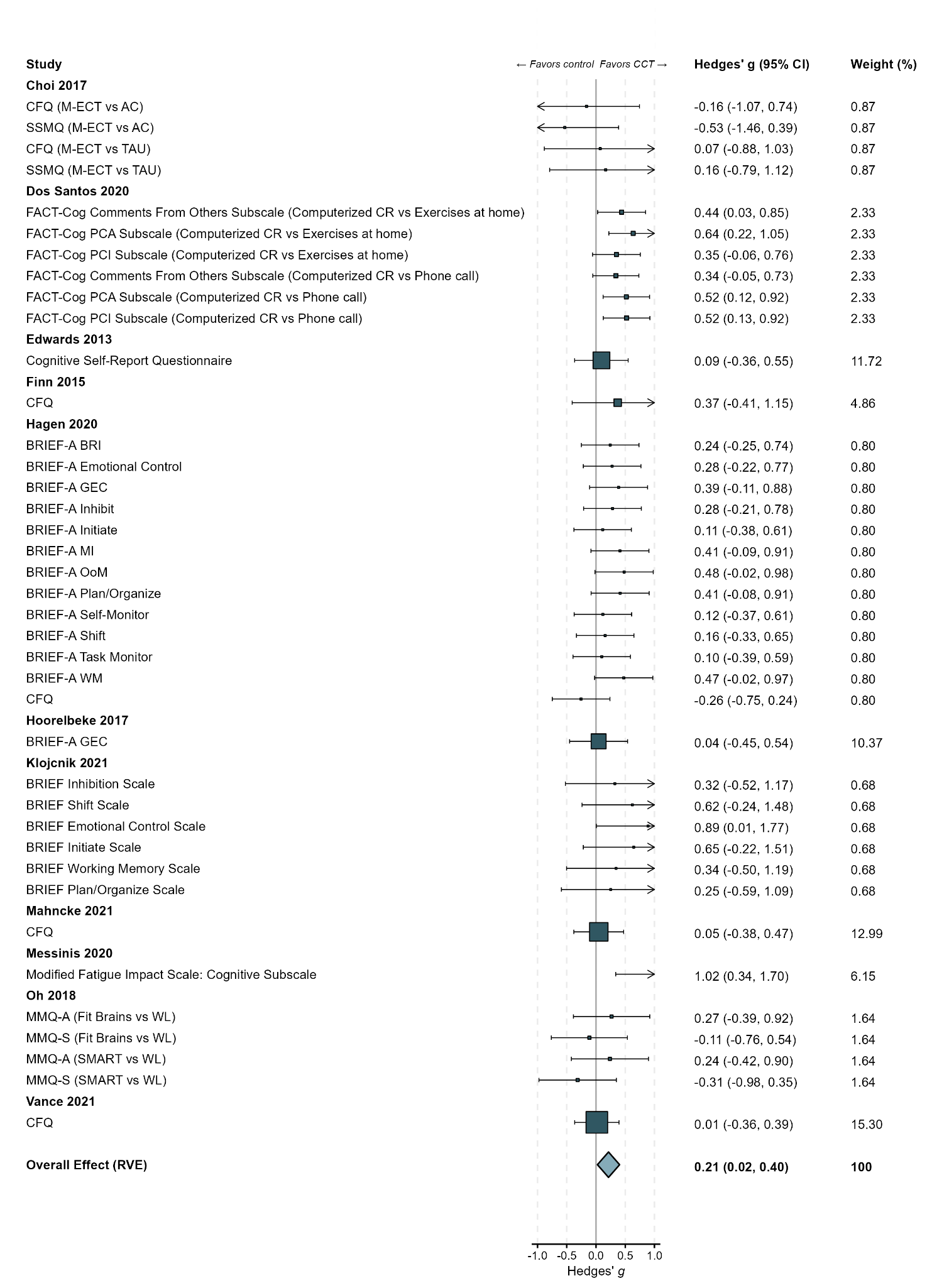

**eFigure11.** Funnel Plot of small study Subjective Cognition
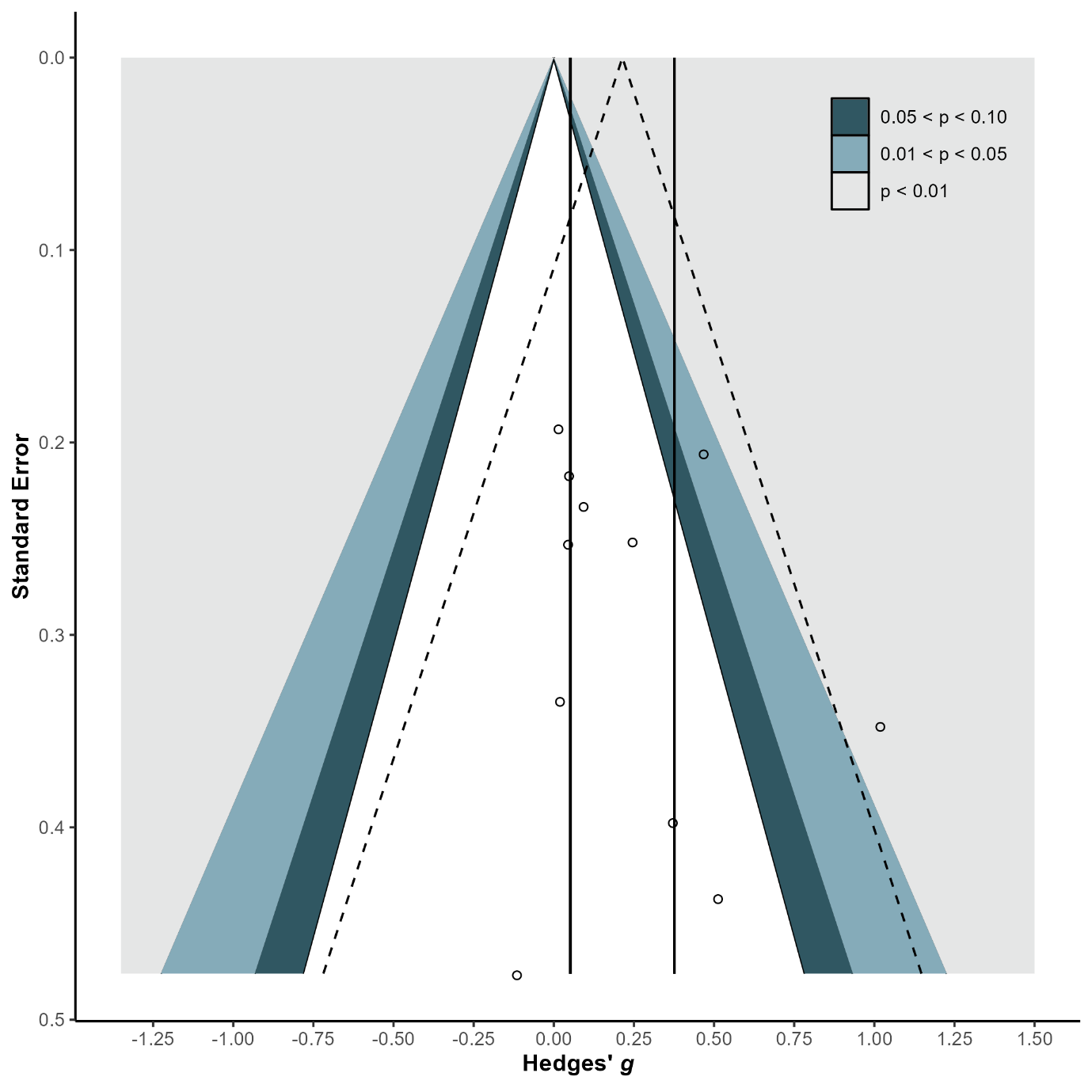

**eFigure 12.** Forest Plot of Global Cognition

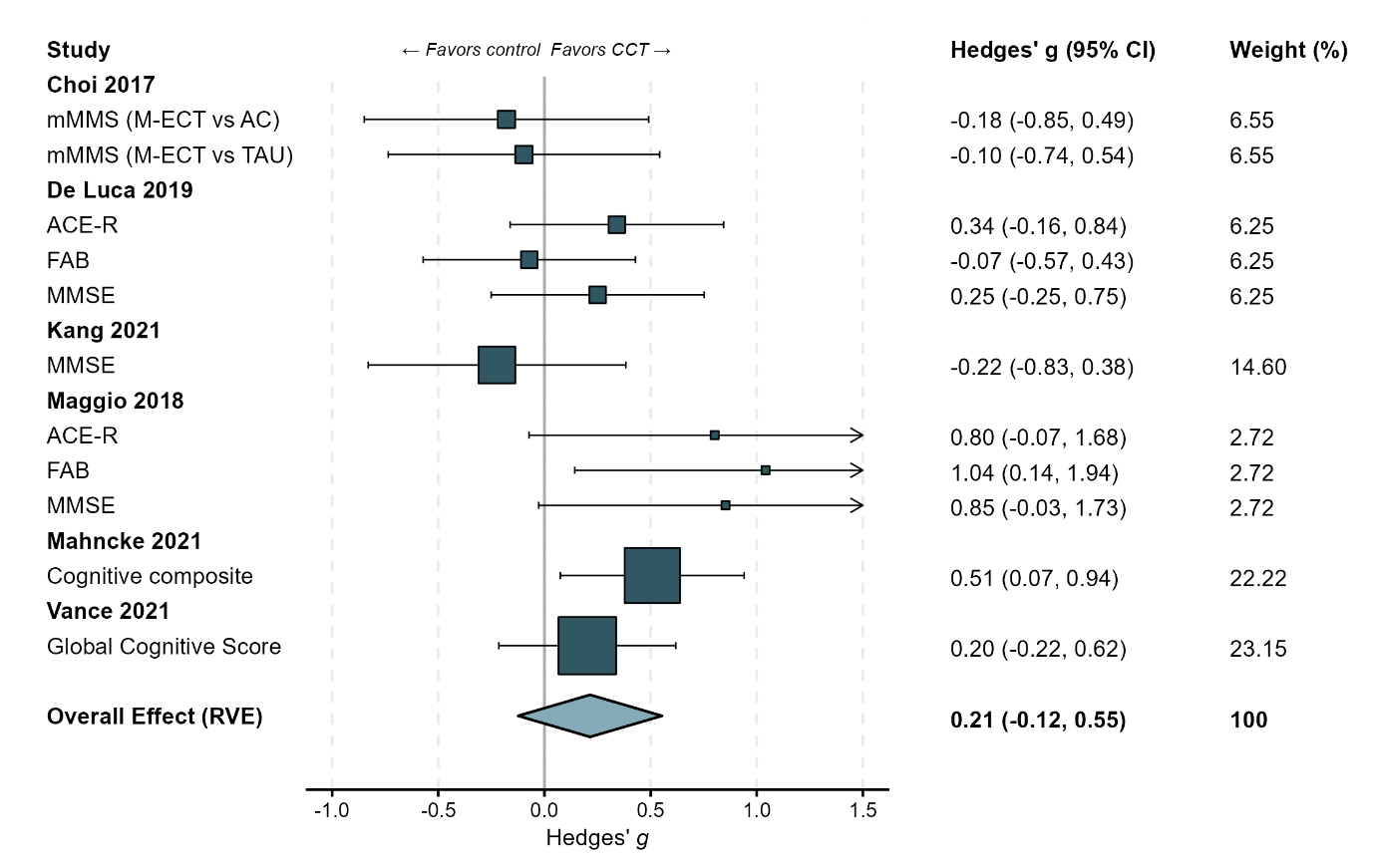

**Figure 13.** Funnel Plot of Global Cognition
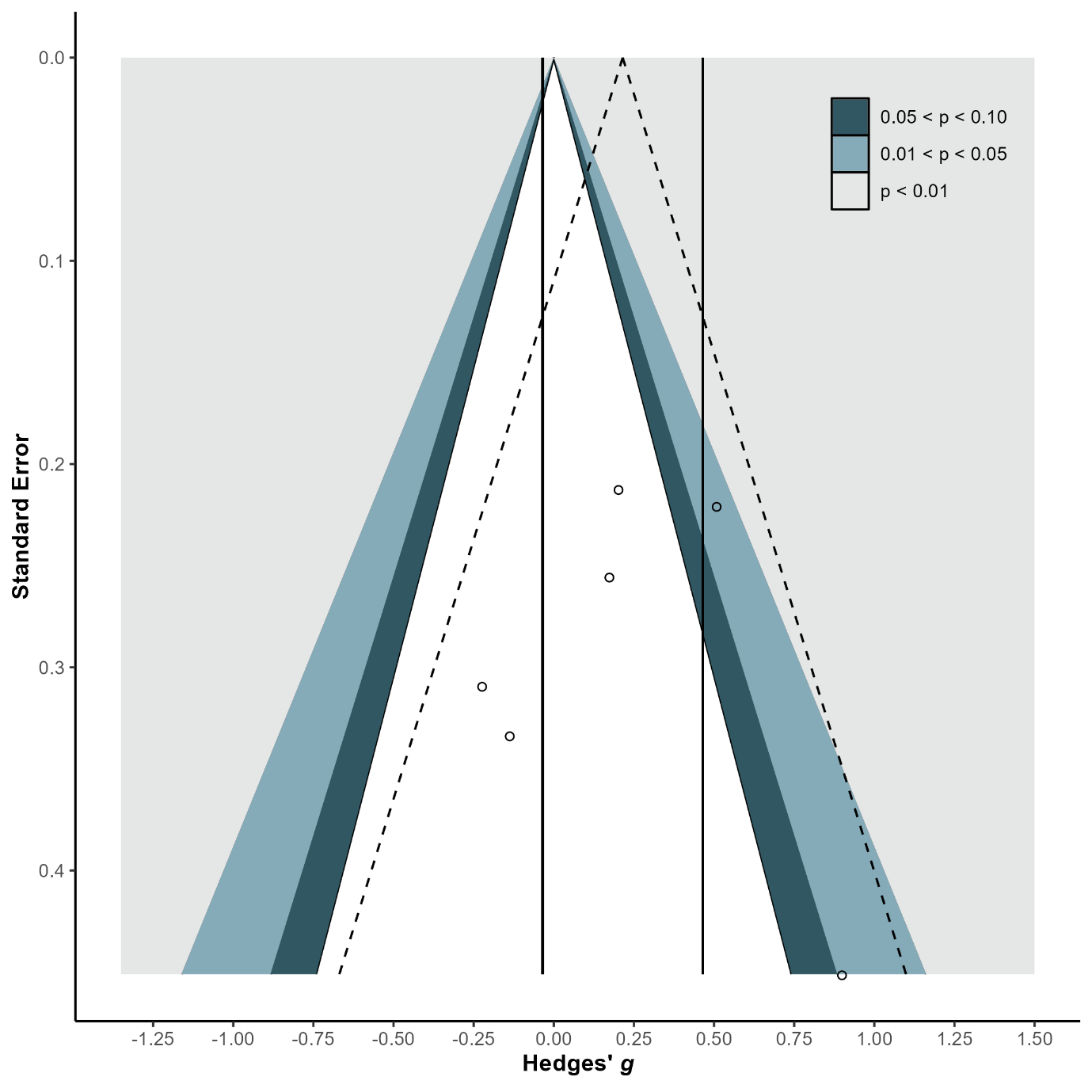

**eFigure 14.** Forest Plot of Fluid Reasoning

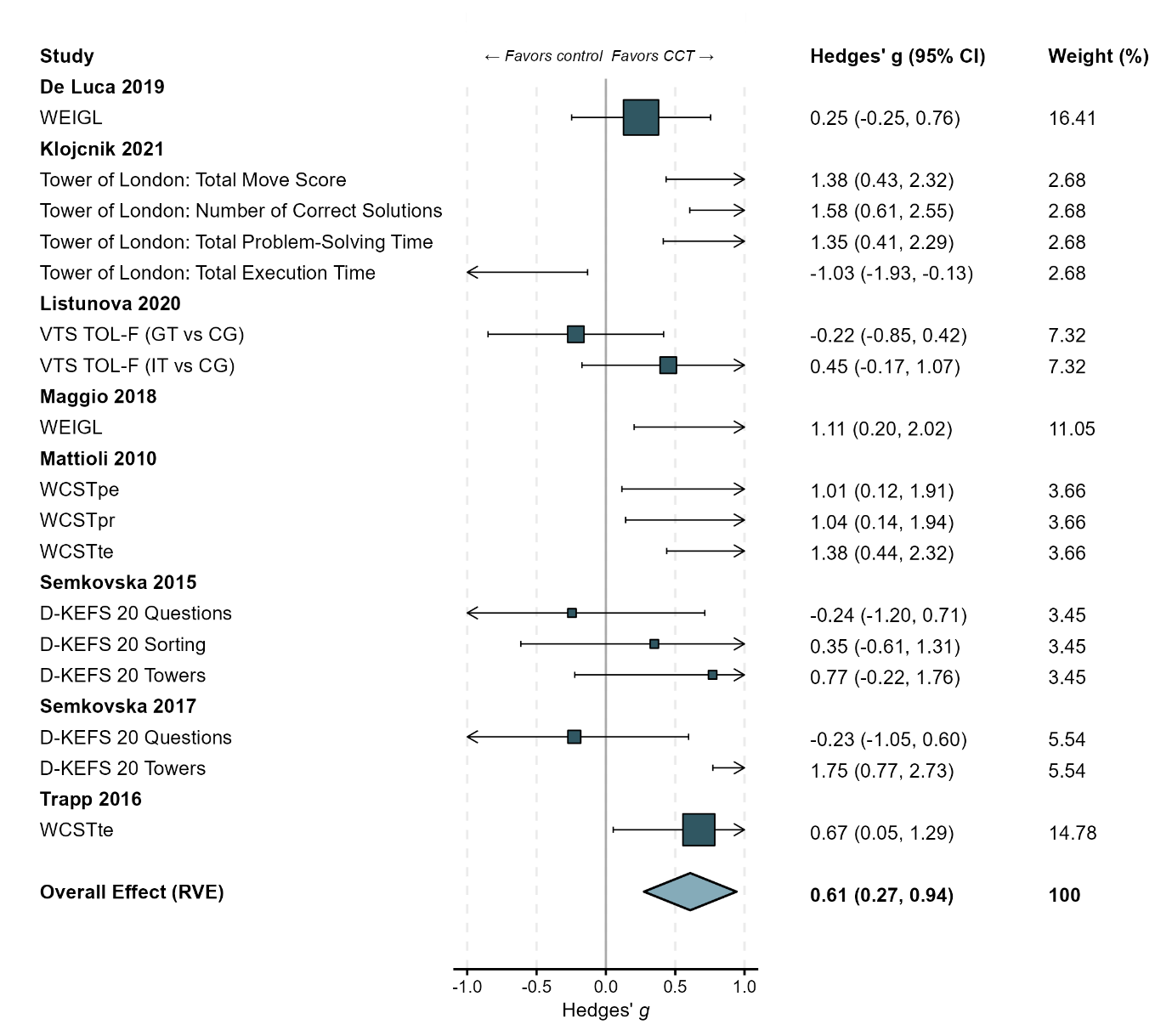

**eFigure 15.** Funnel Plot of Fluid Reasoning
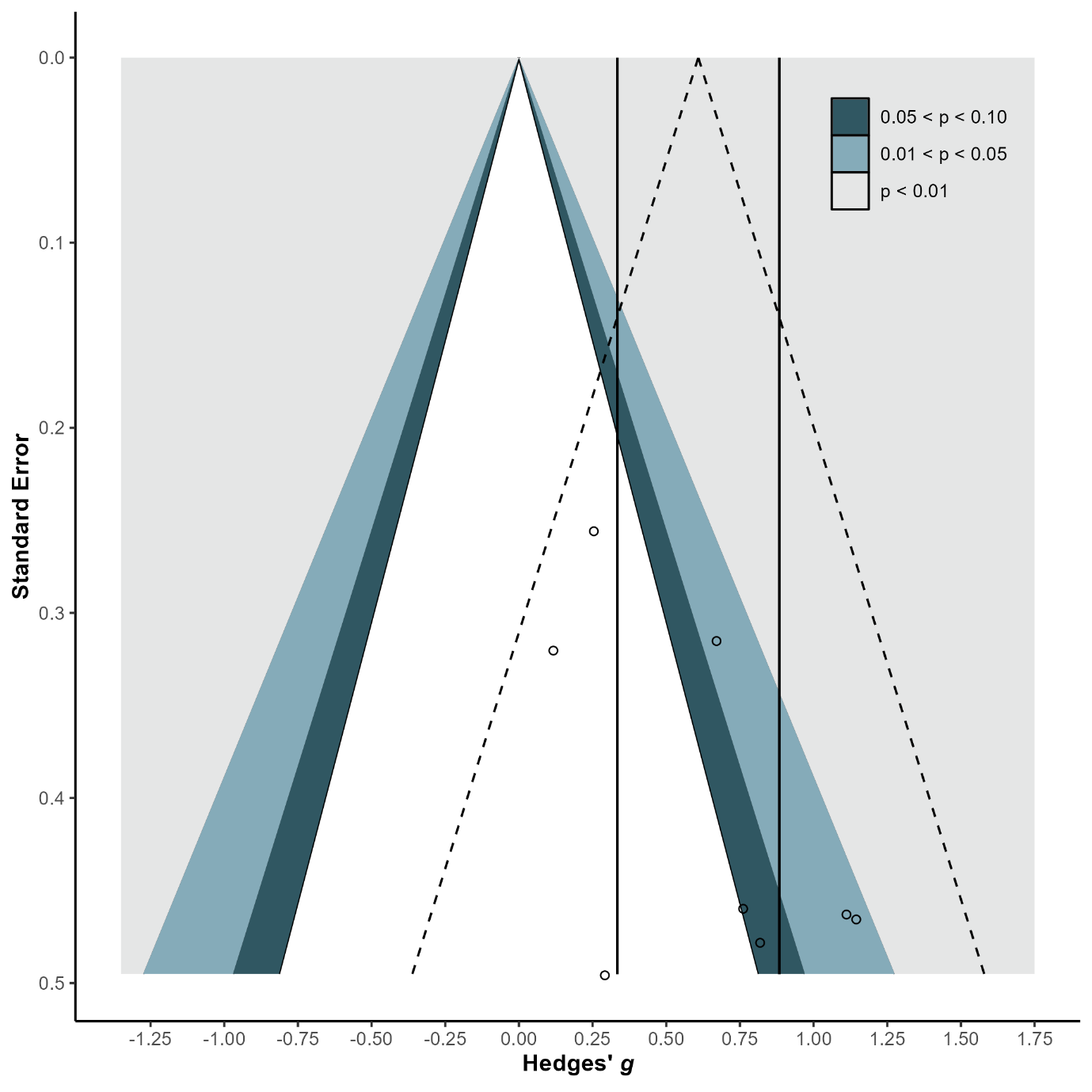

**eFigure 16.** Forest Plot of Abstract Reasoning

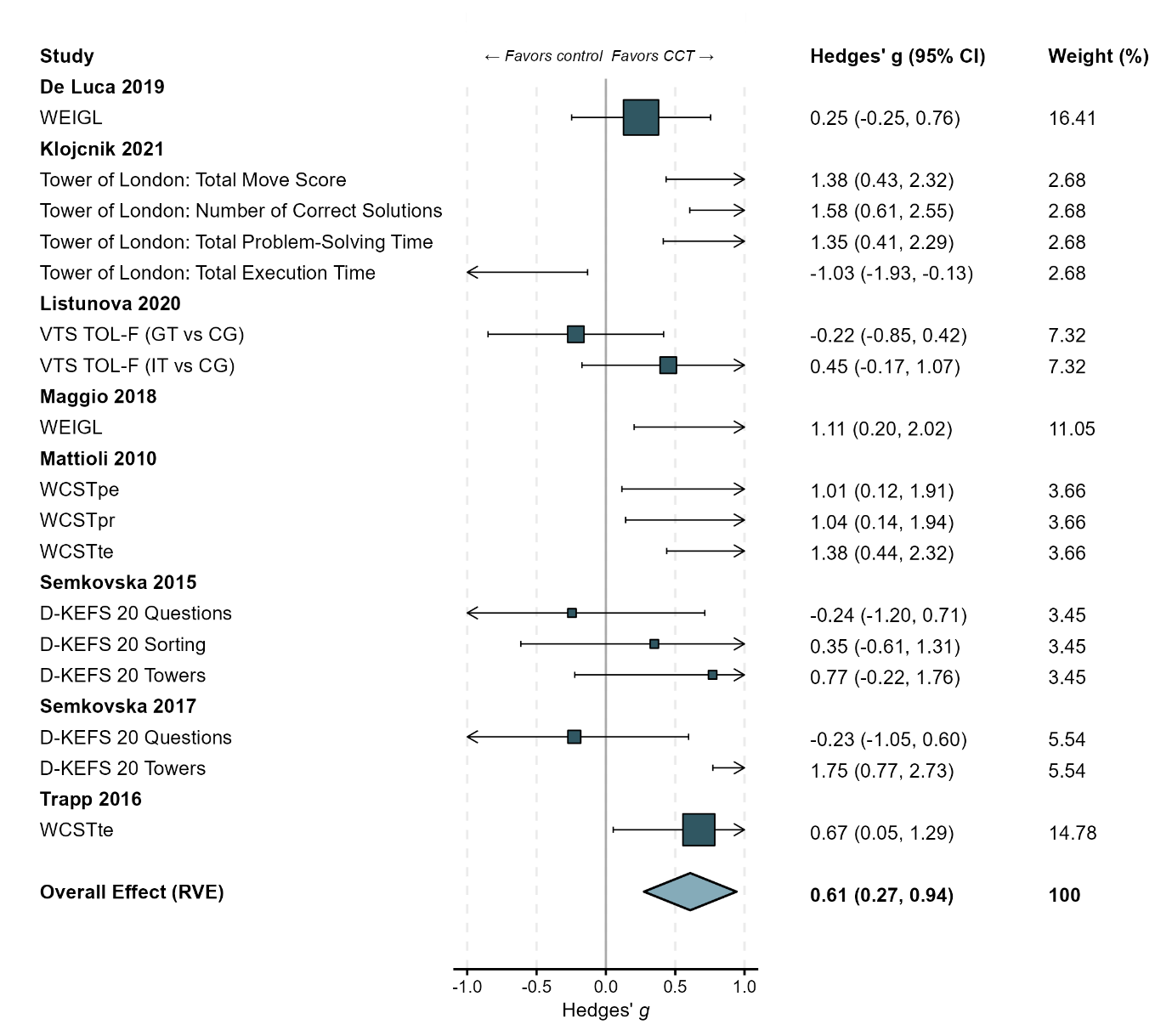

**eFigure 17.** Funnel Plot of Abstract Reasoning

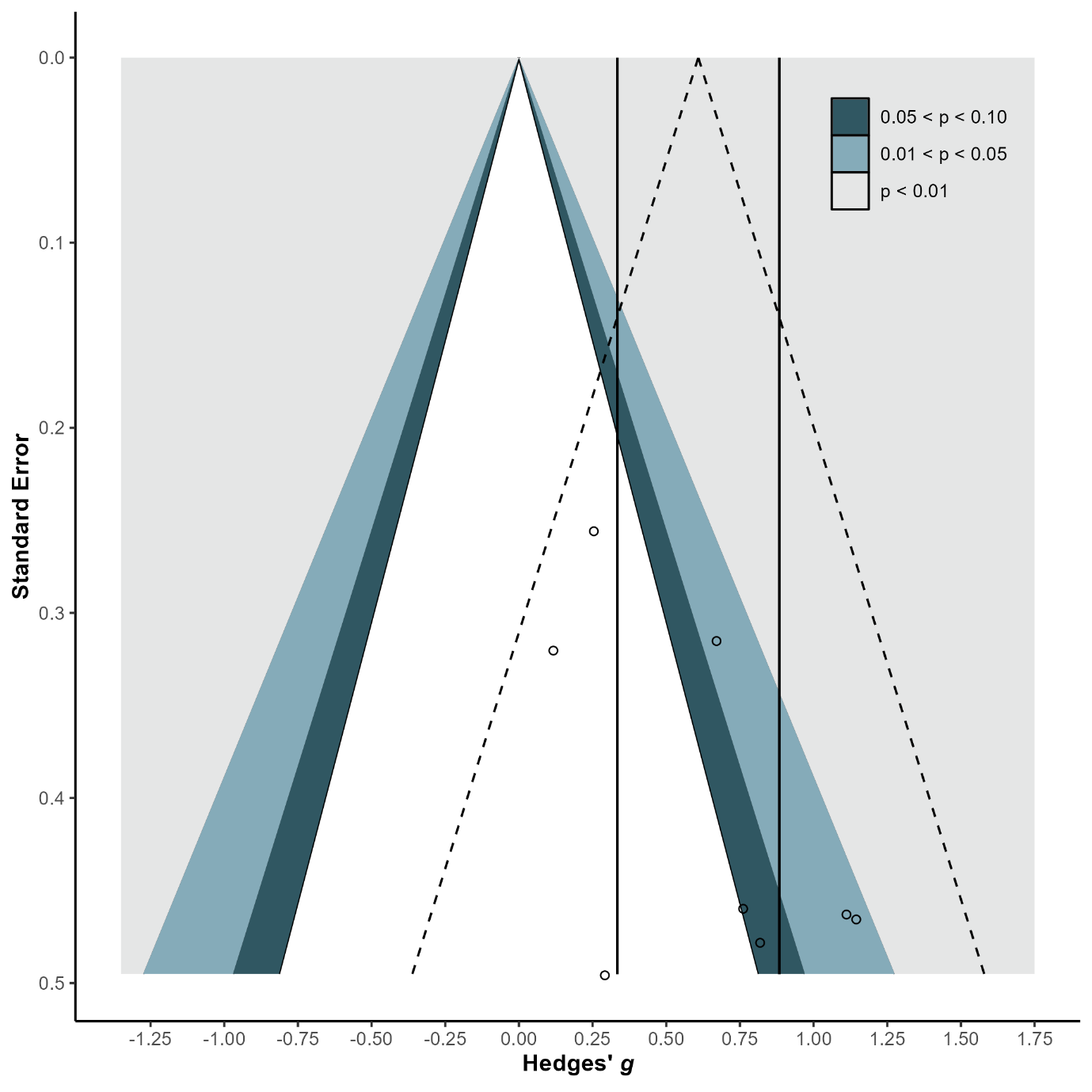

**eFigure 18.** Forest Plot of Long-term Memory and Retrieval

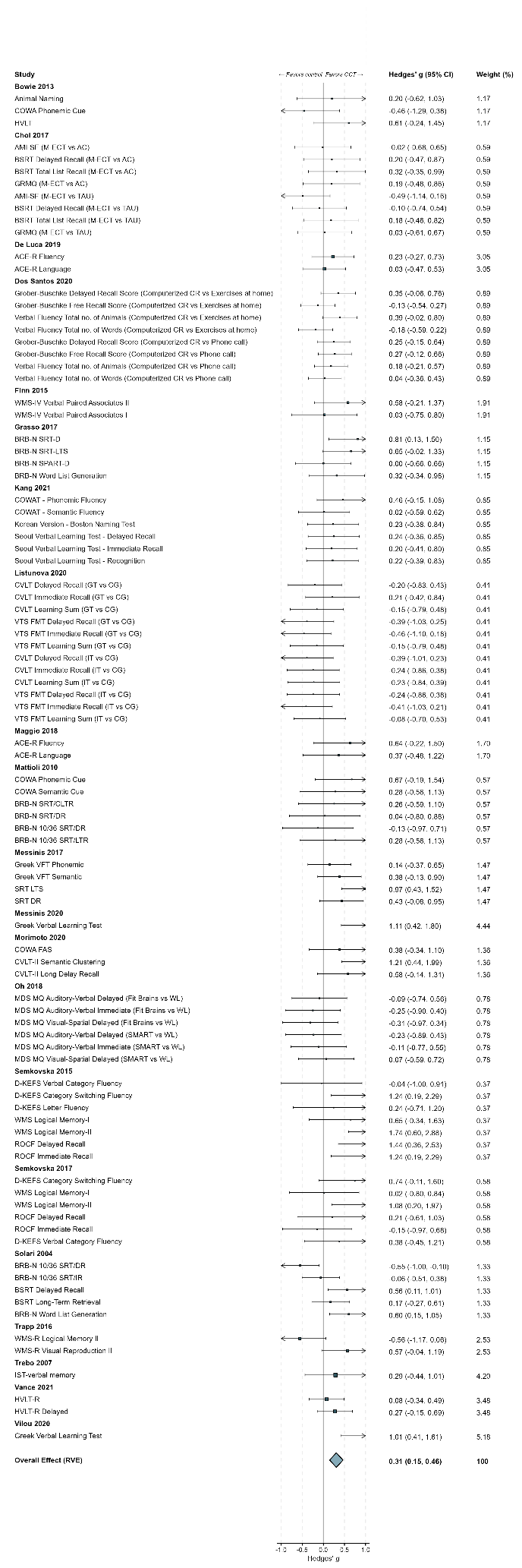

**eFigure 18.** Forest Plot of Long-term Memory and Retrieval…continued…

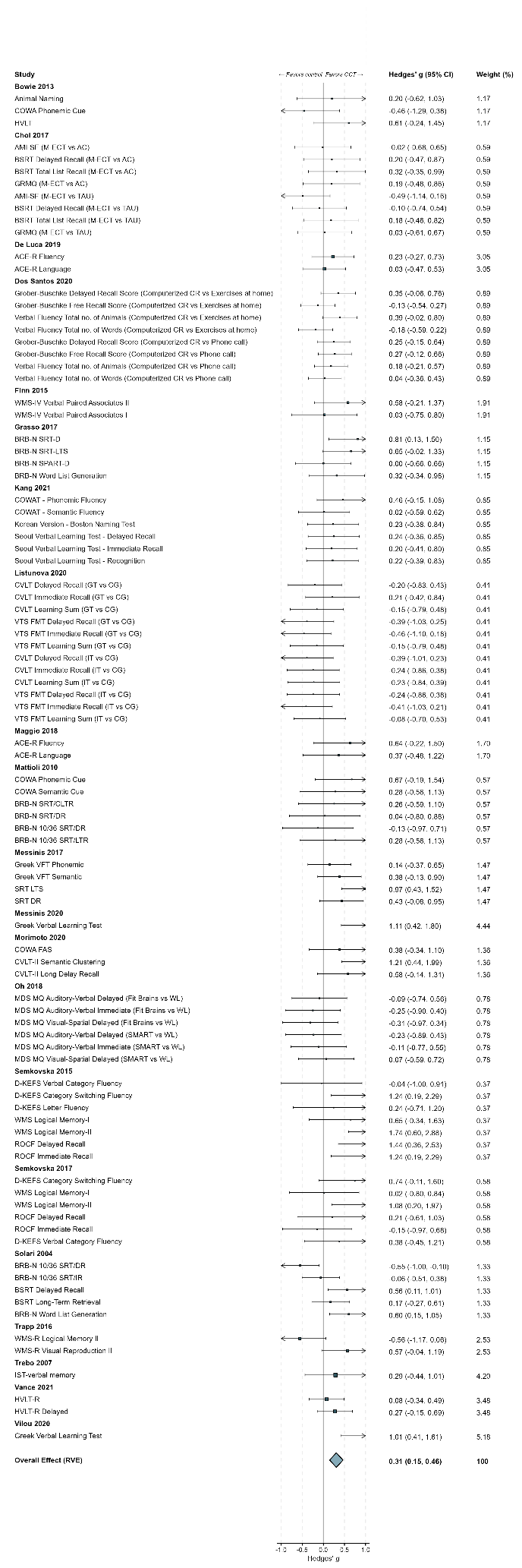

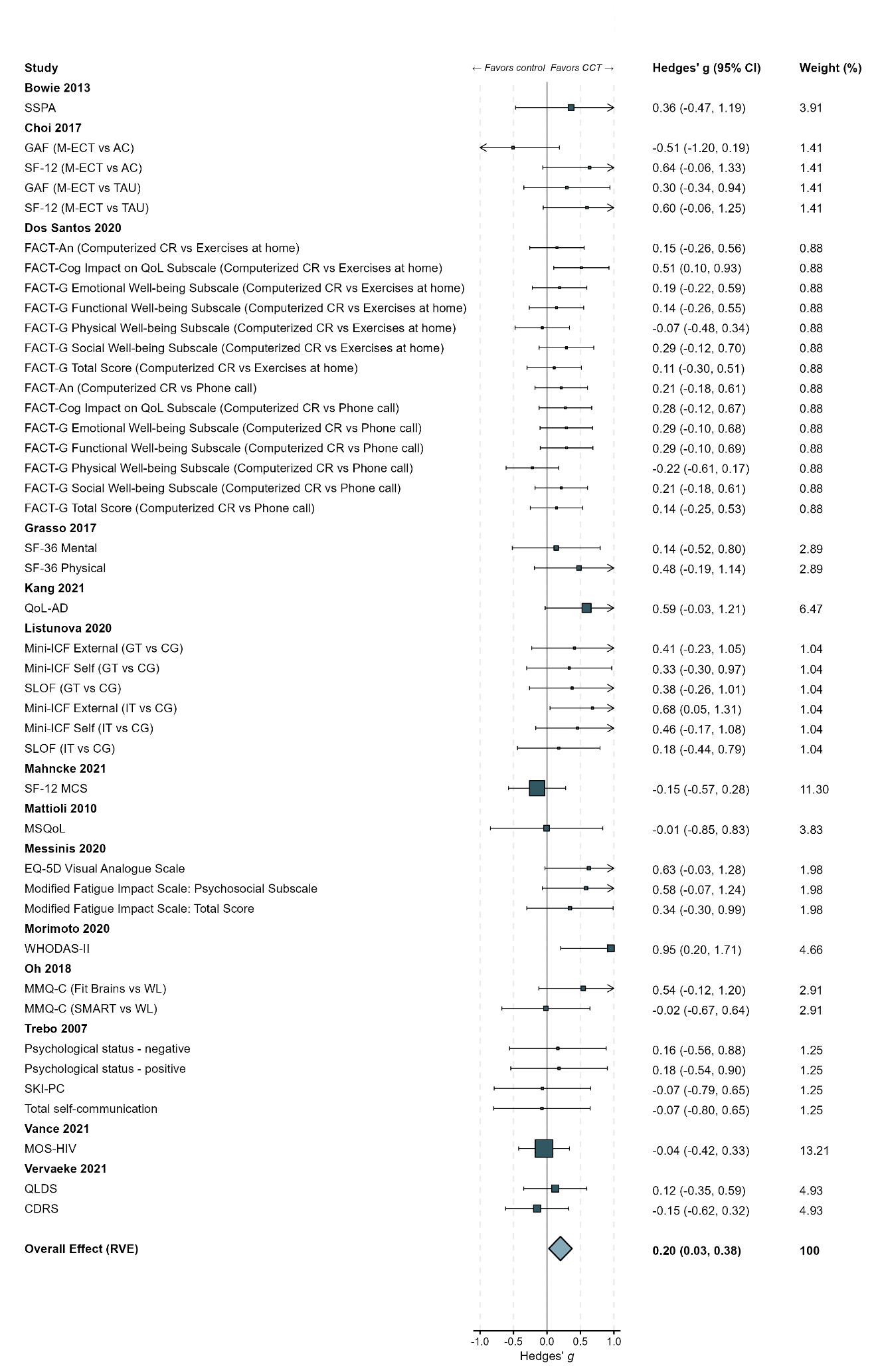

**eFigure 19.** Funnel Plot of Long-term Memory and Retrieval
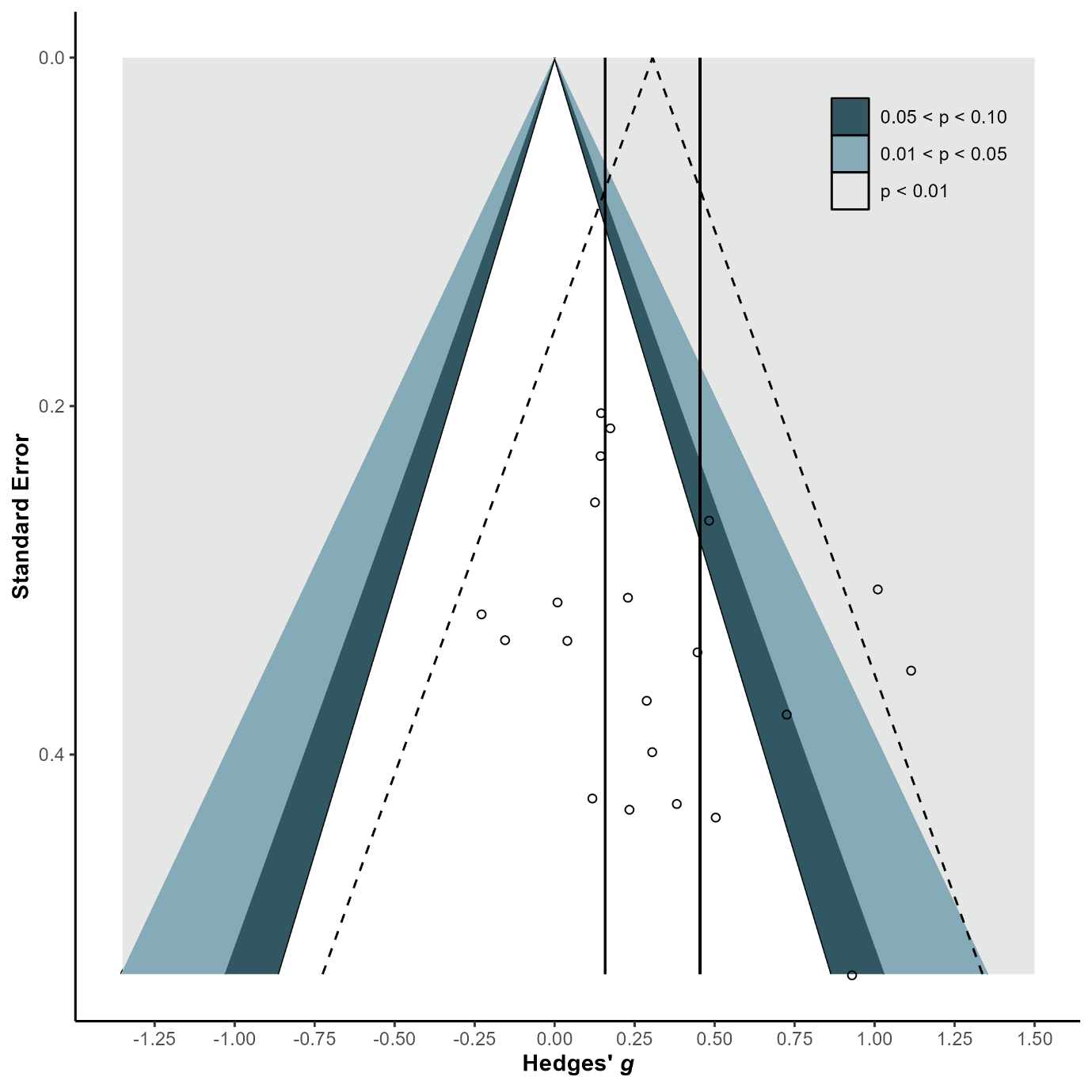

**eFigure 20.** Forest Plot of Learning/Encoding Efficiency

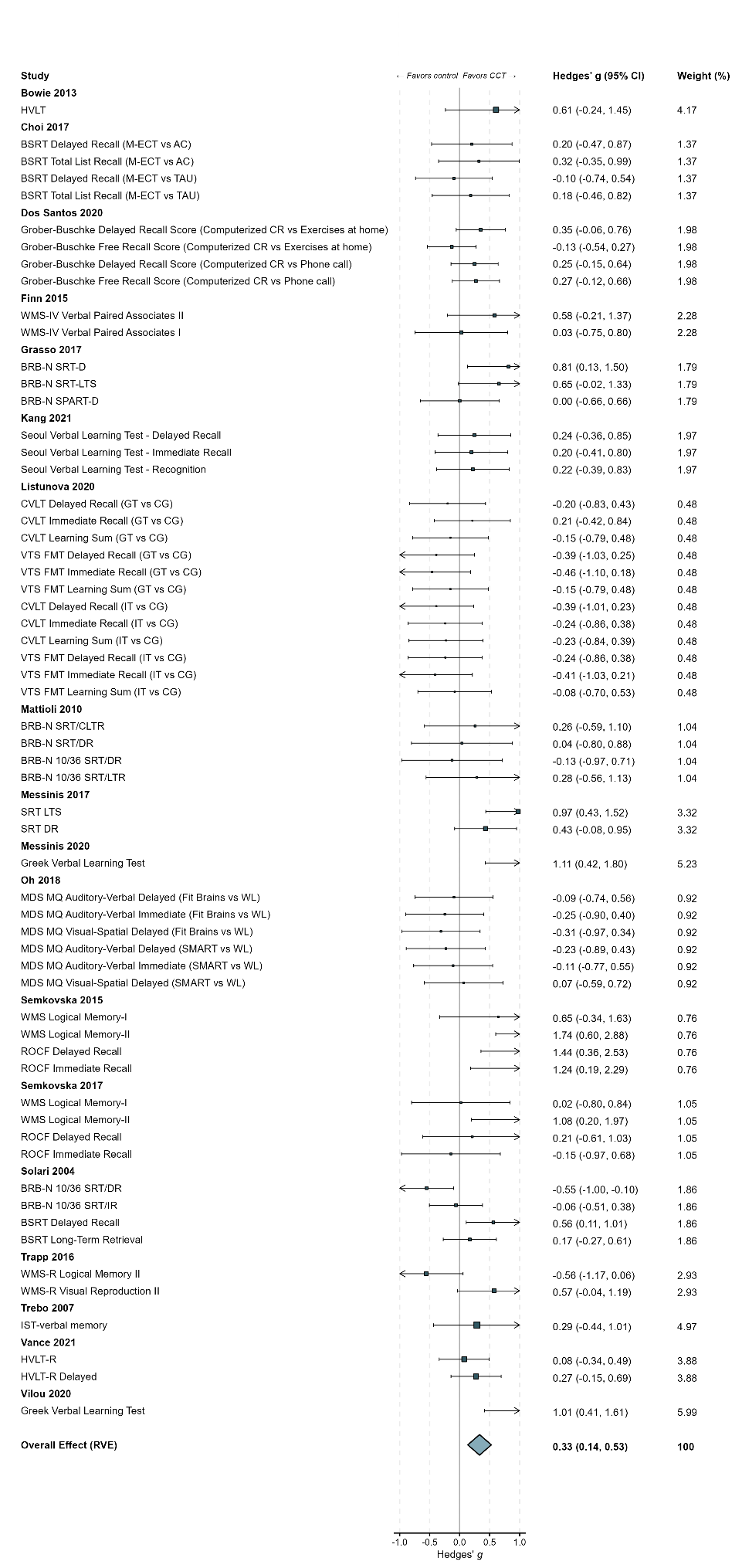

**eFigure 20.** Forest Plot of Learning/Encoding Efficiency…continued…

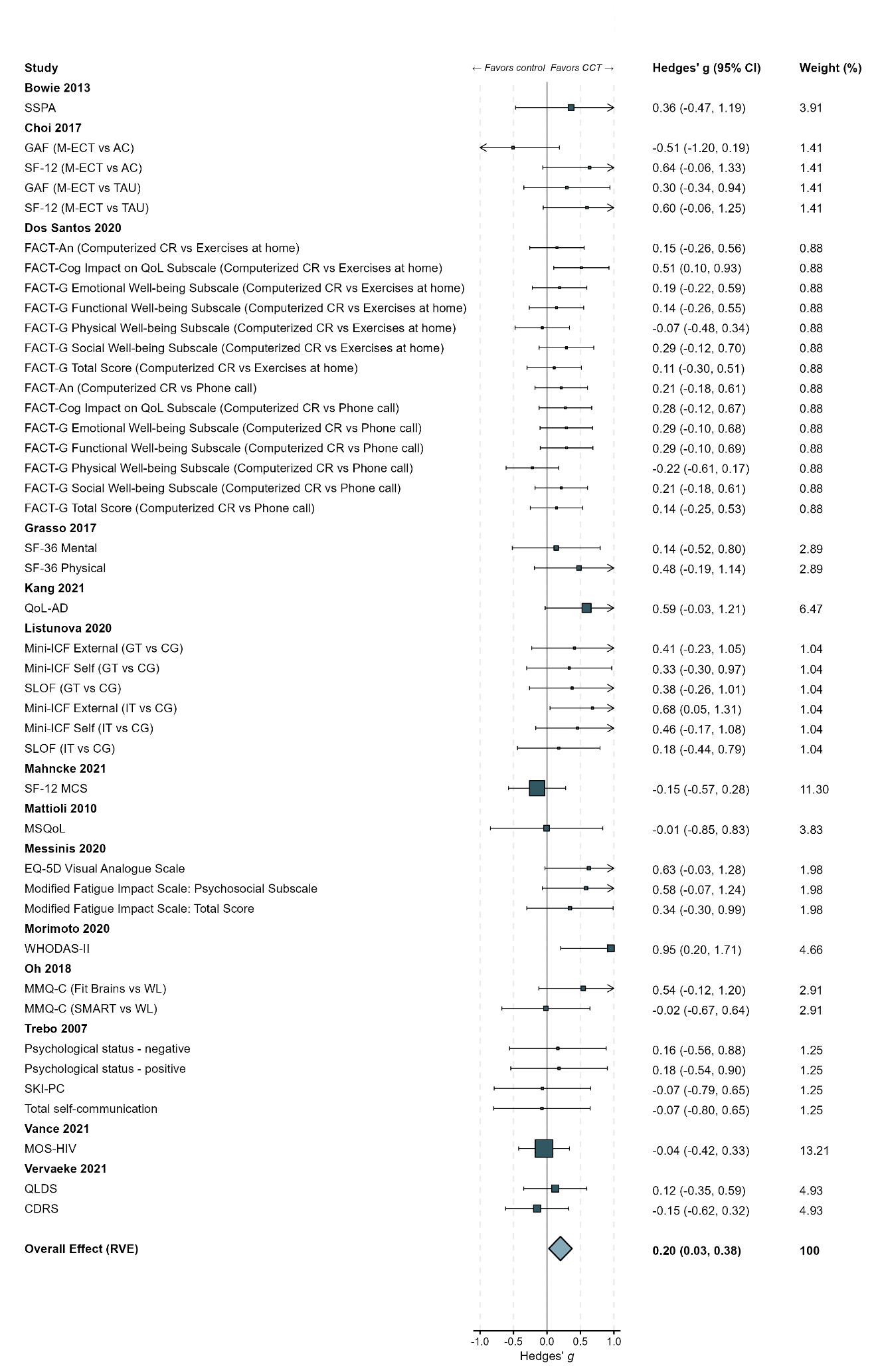

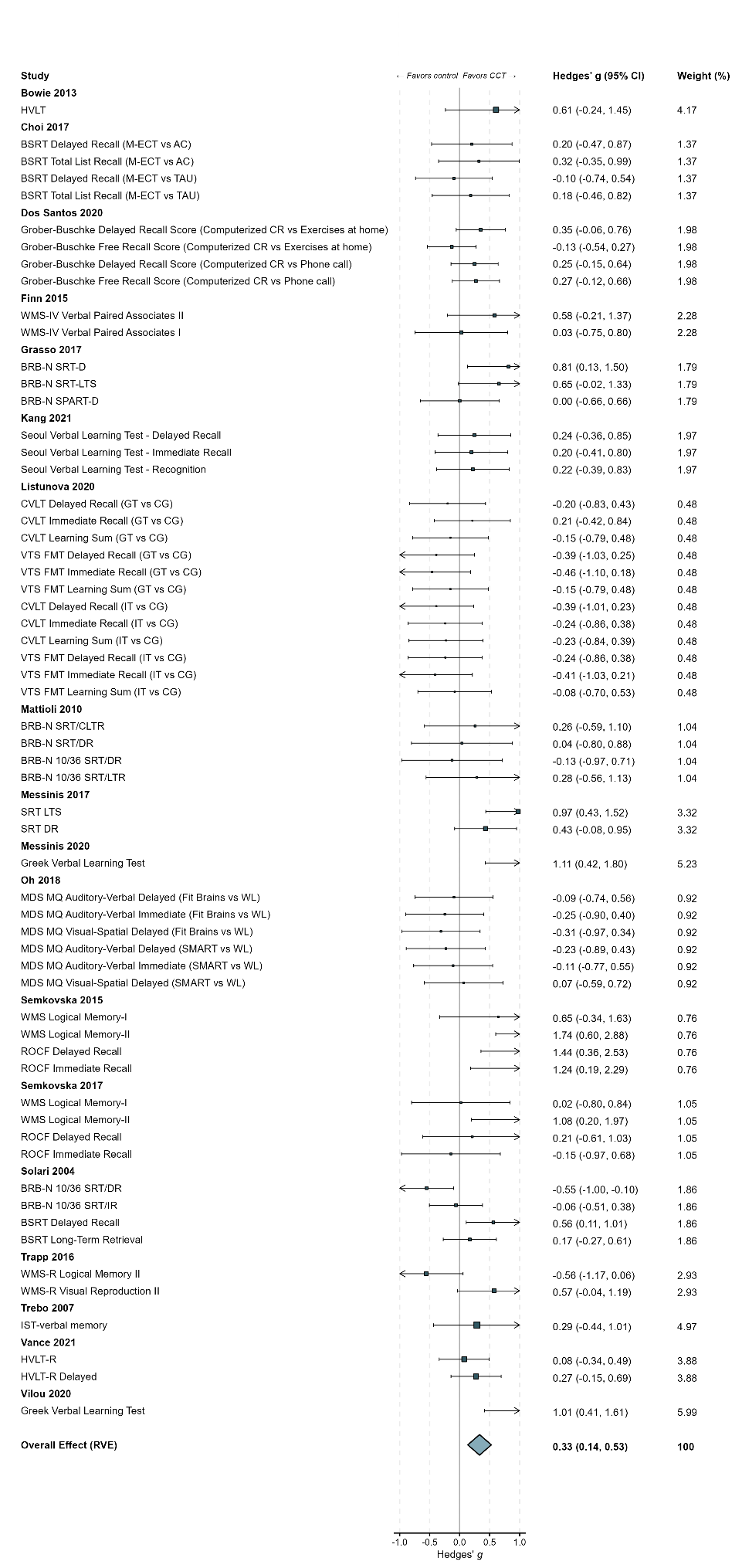

**eFigure 21.** Funnel Plot of Learning/Encoding Efficiency
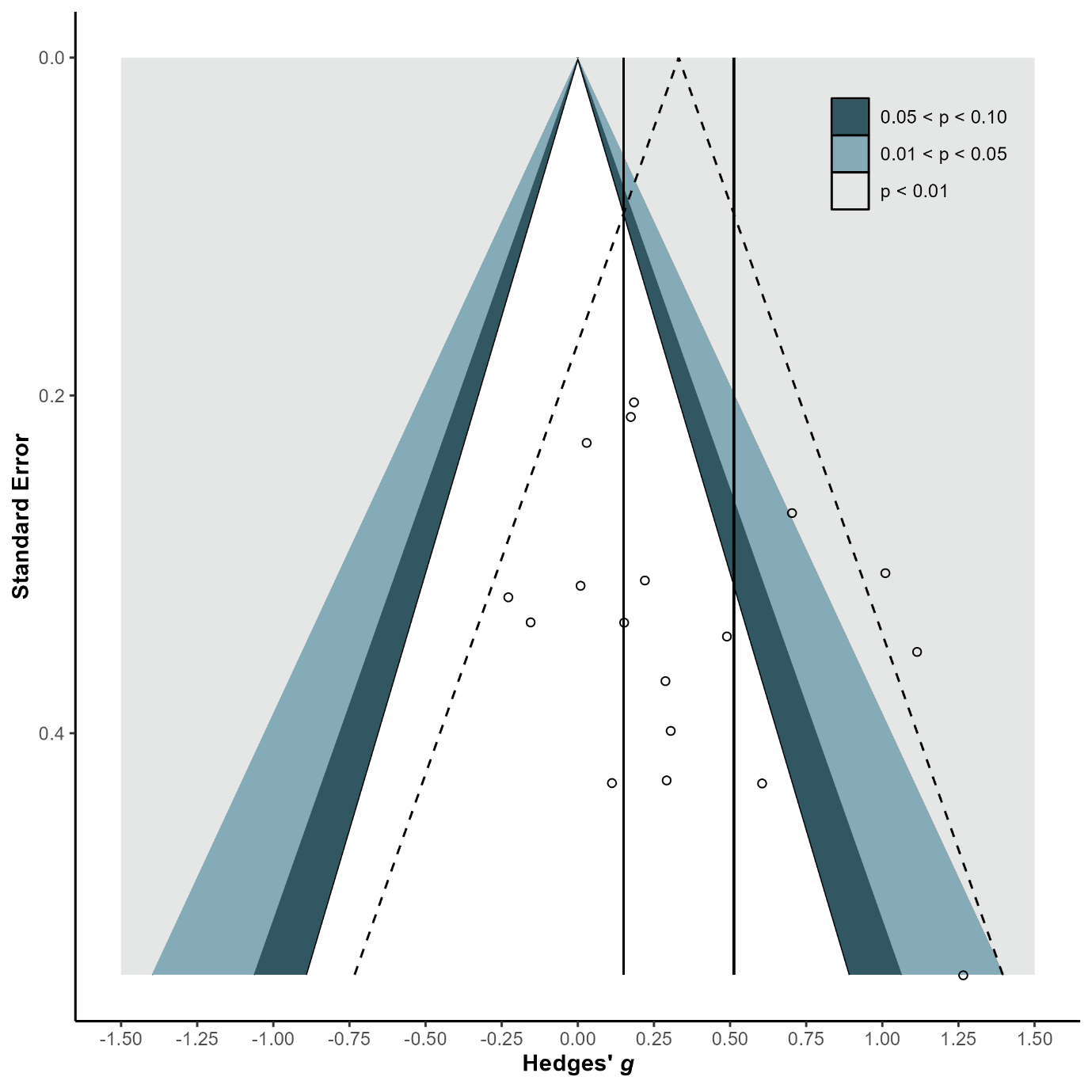

**eFigure 22.** Forest Plot of Retrieval Fluency

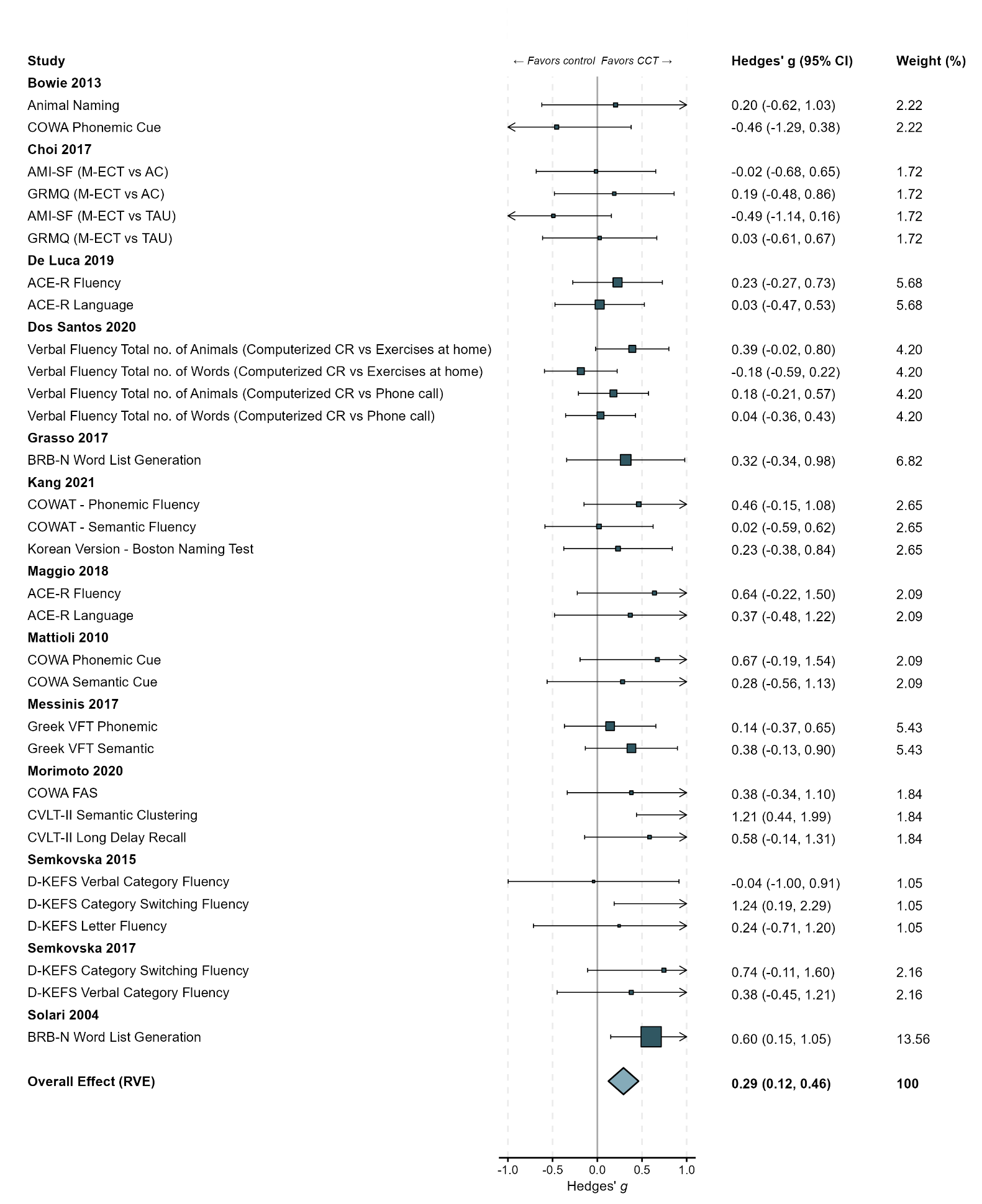

**eFigure 23.** Funnel Plot of Retrieval Fluency
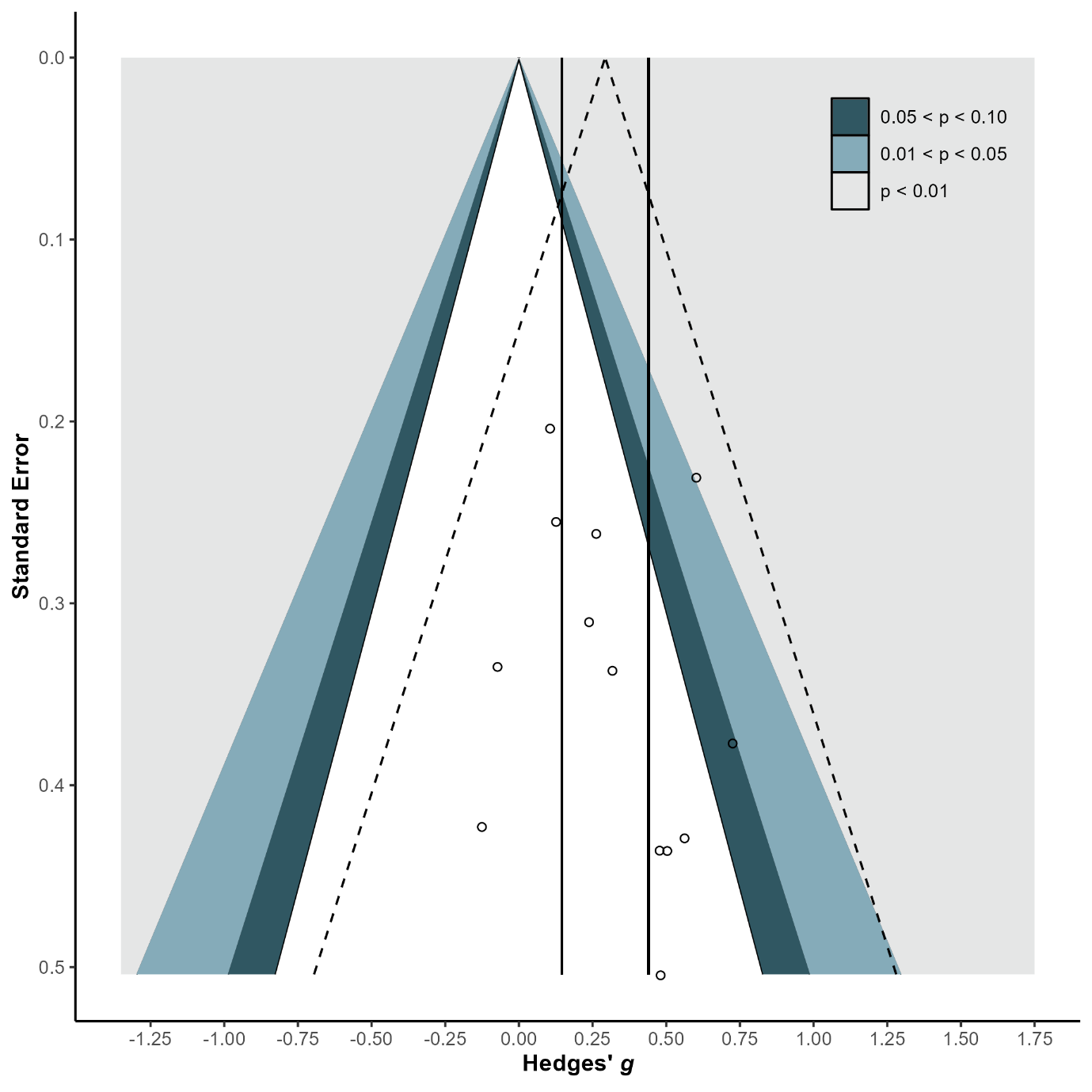

**eFigure 24.** Forest Plot of General Short-term Memory

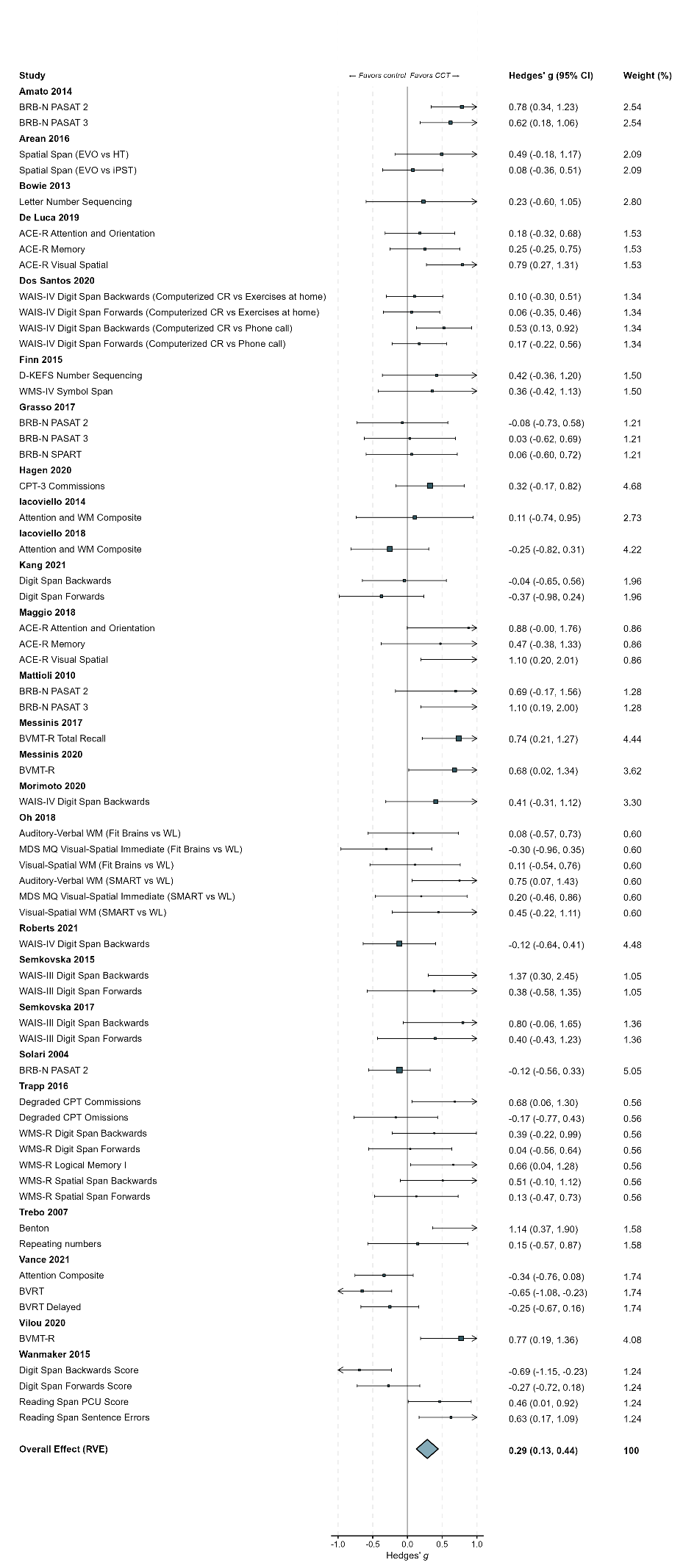

**eFigure 24.** Forest Plot of General Short-term Memory…continued…

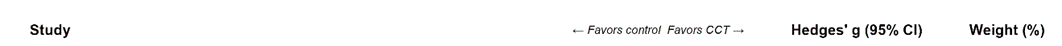

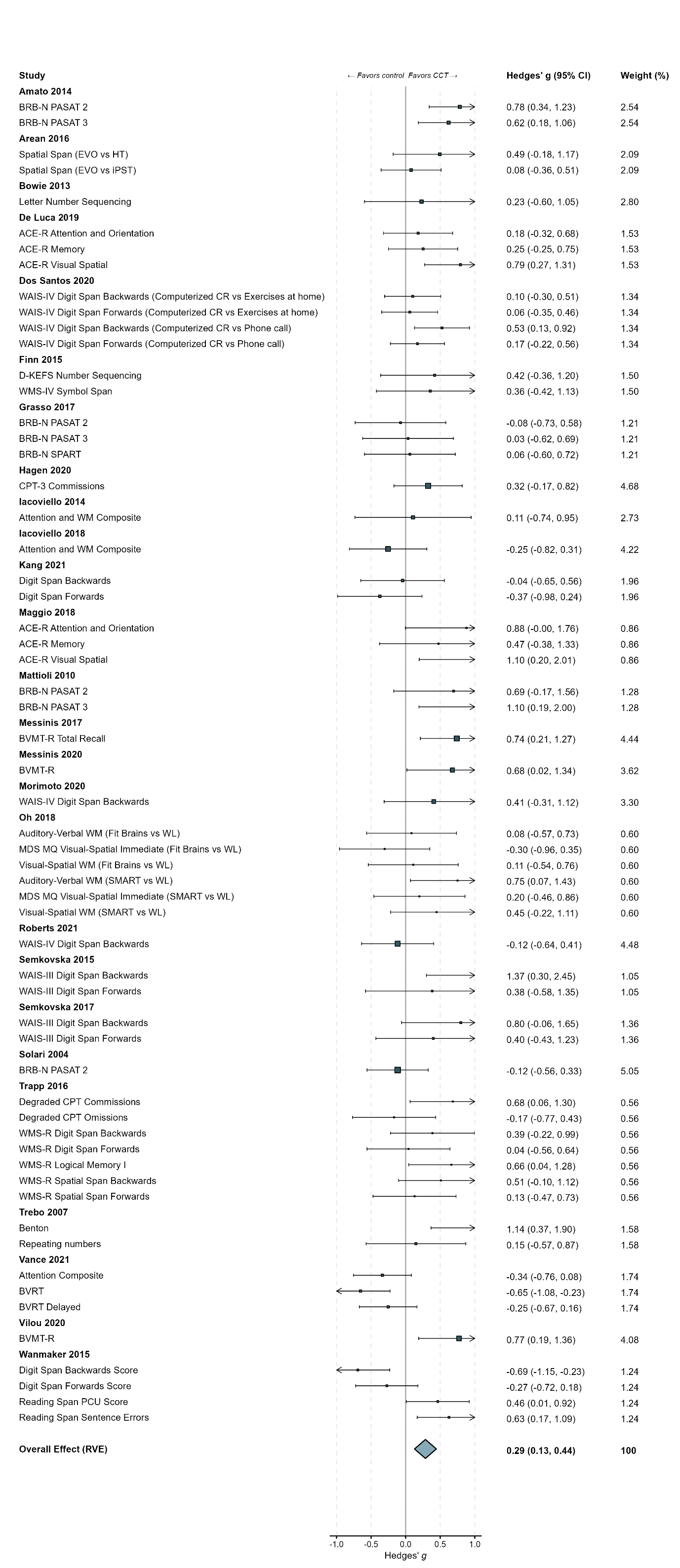

**eFigure 25.** Funnel Plot of General Short-term Memory
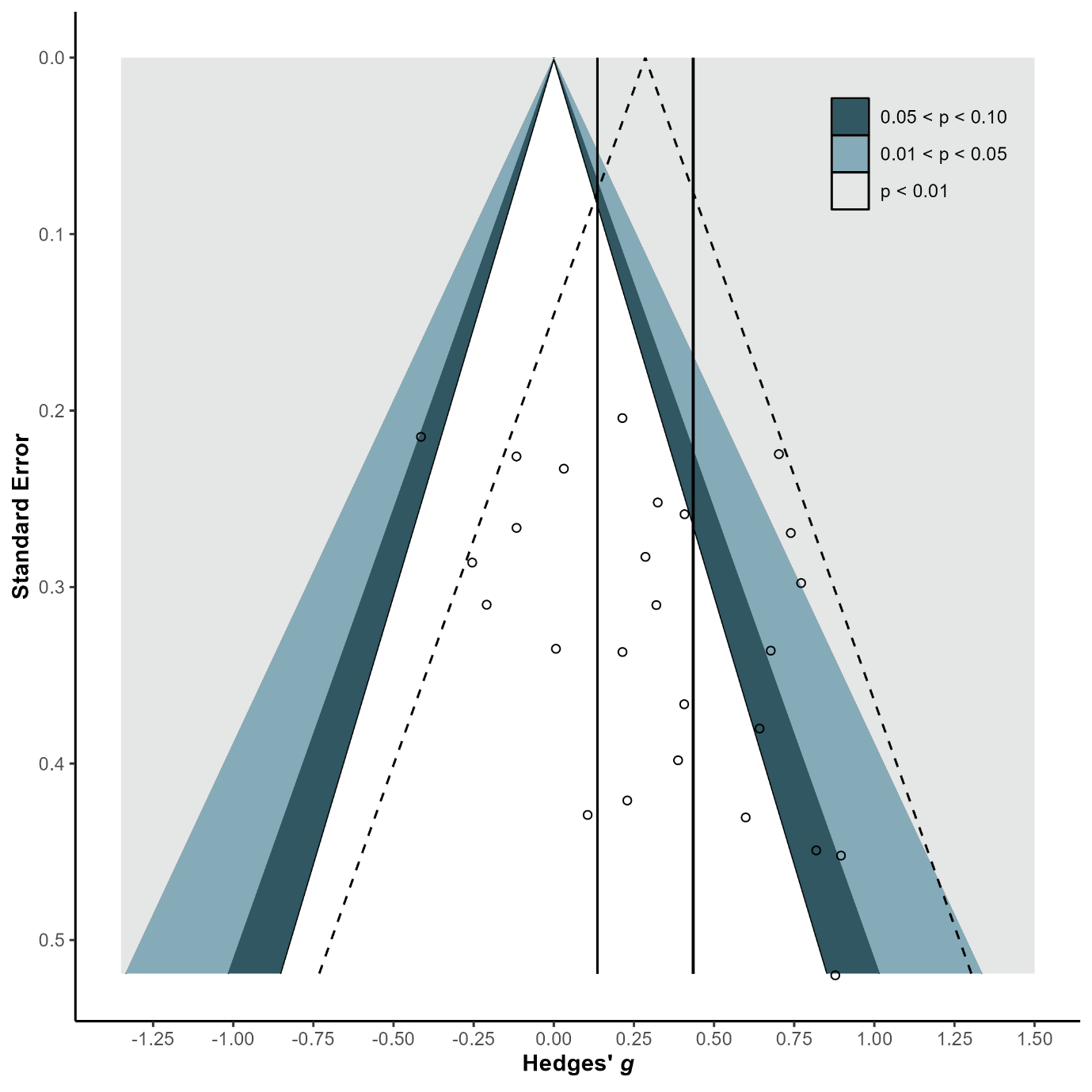

**eFigure 26.** Forest Plot of High Working Memory

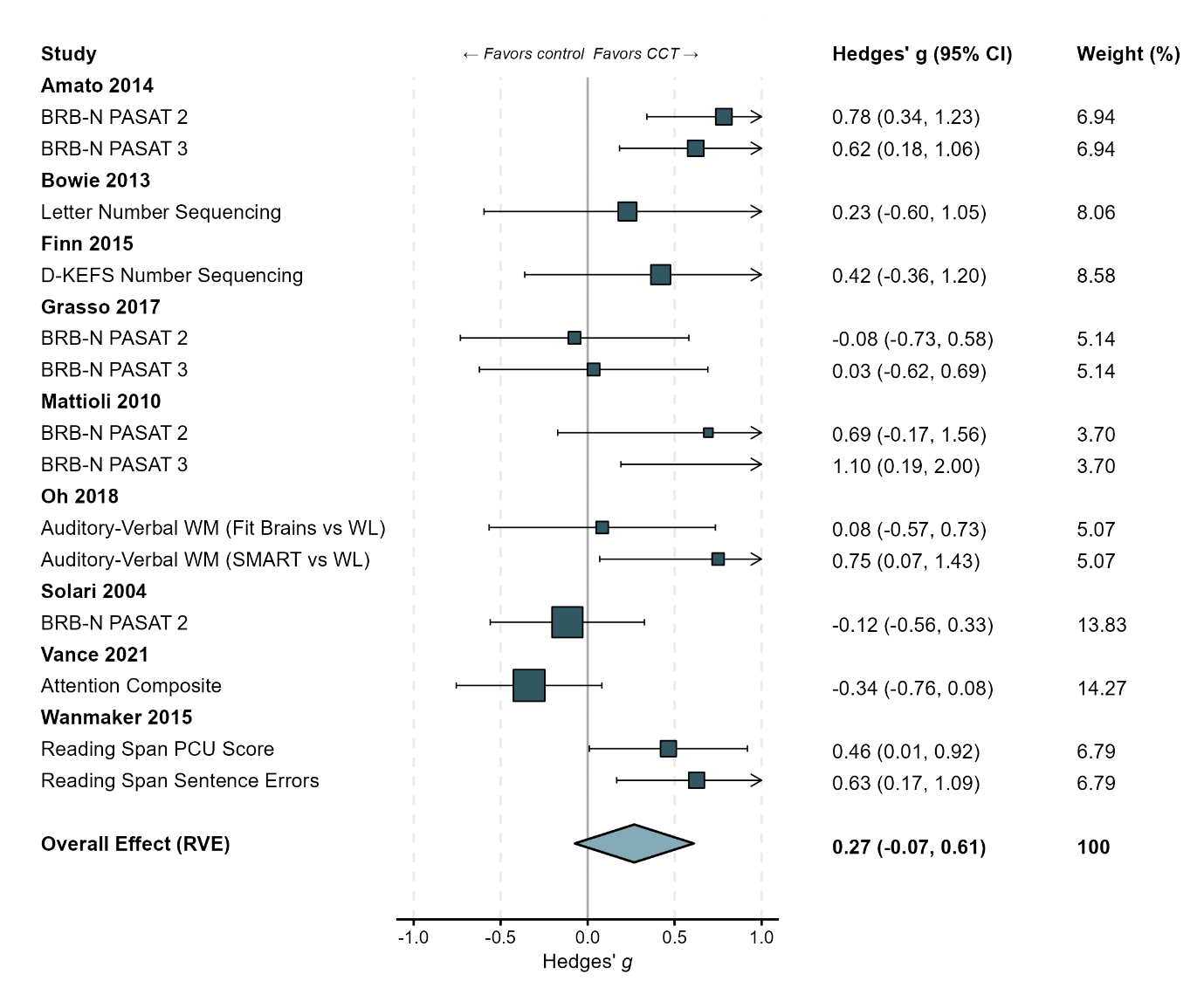

**eFigure 27.** Funnel Plot of High Working Memory
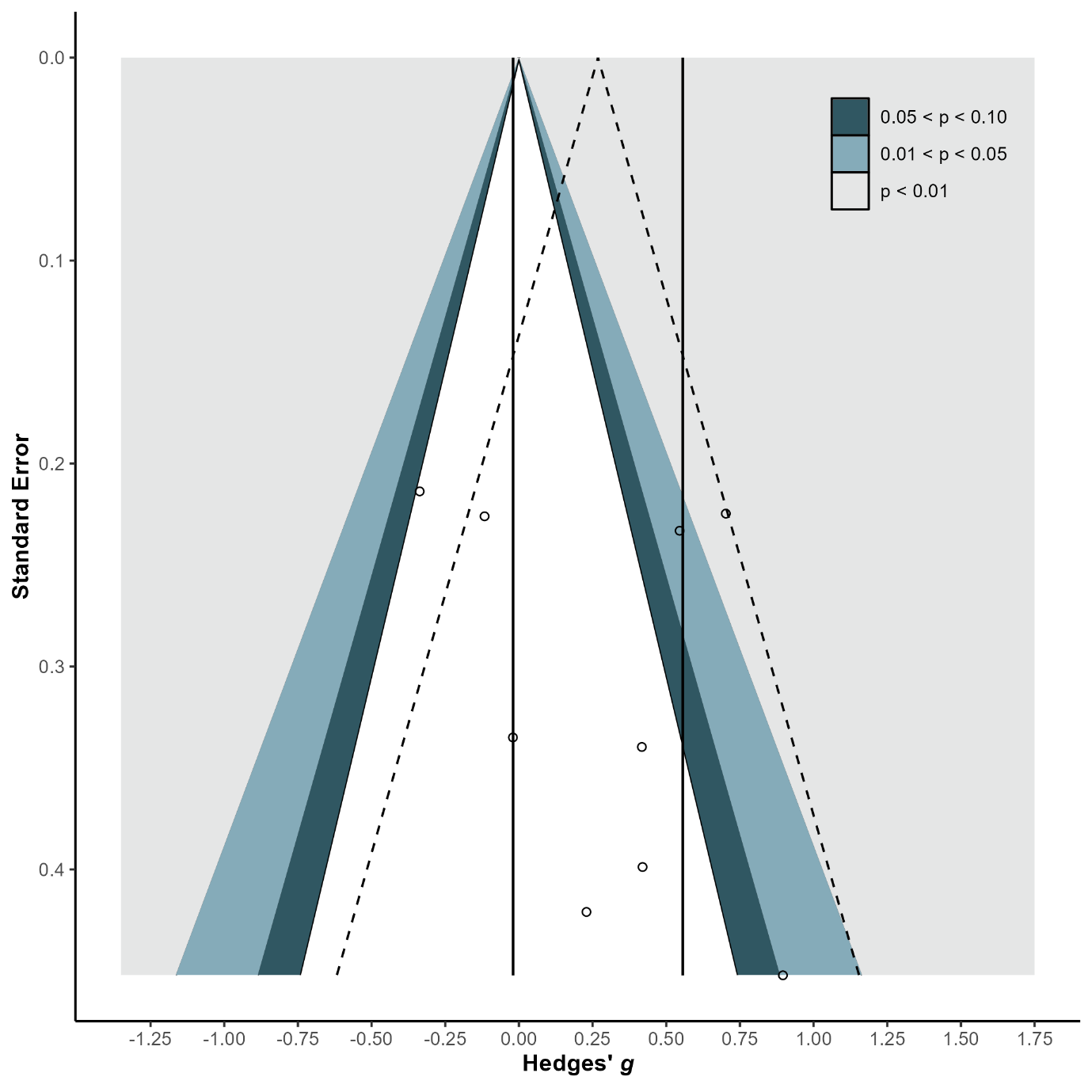

**eFigure 28.** Forest Plot of Low Working Memory

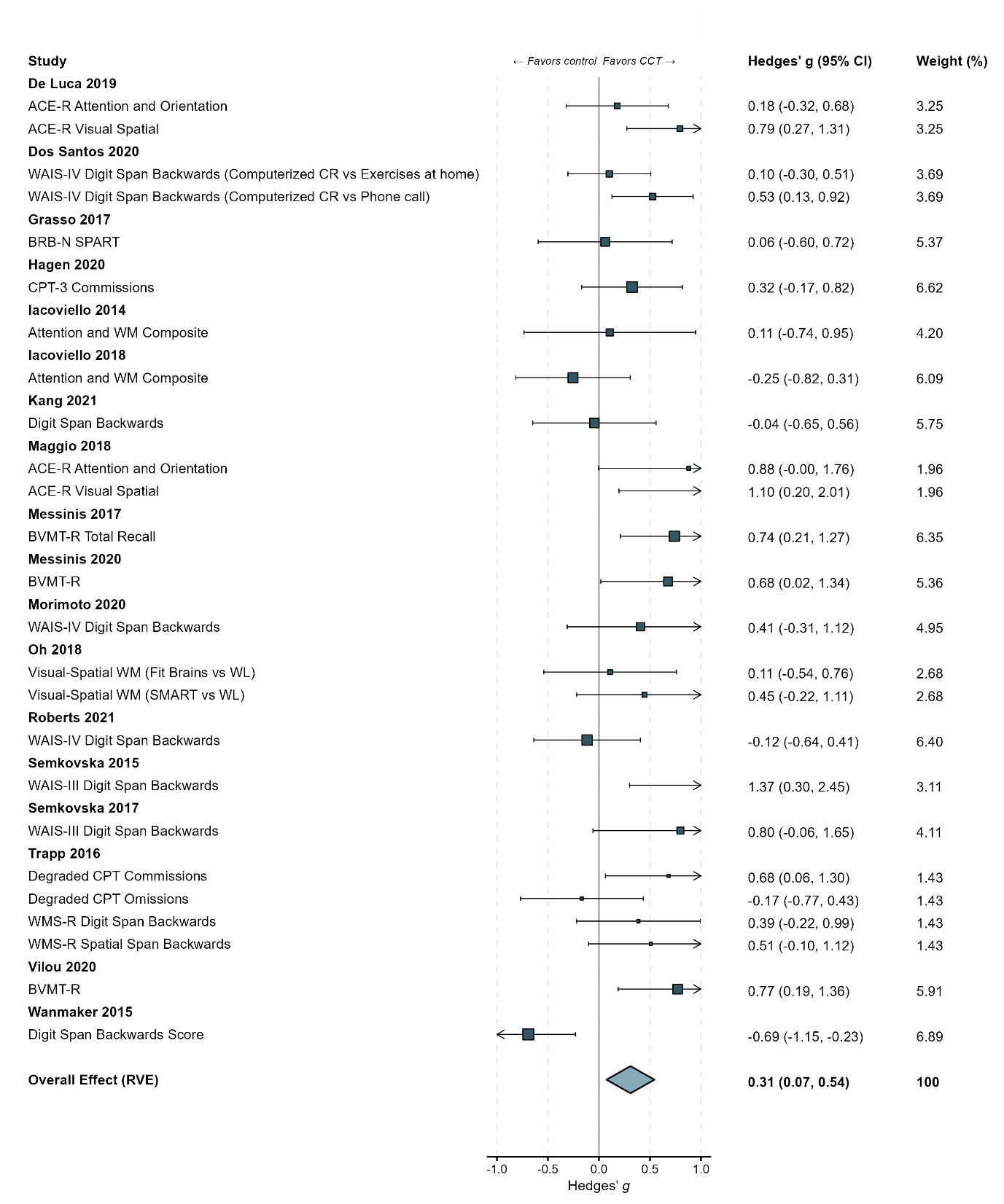

**eFigure 29.** Funnel Plot of Low Working Memory
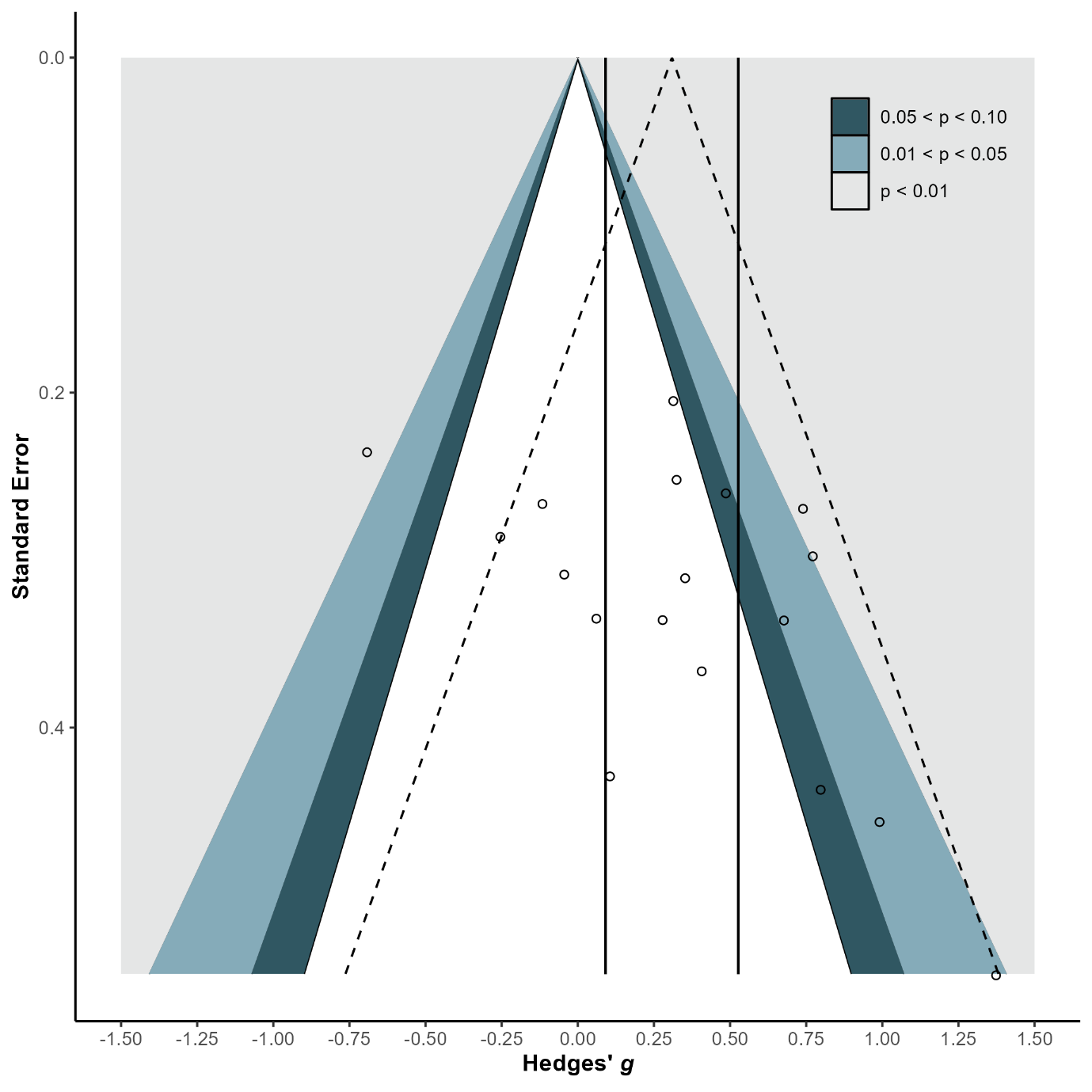

**eFigure 30.** Forest Plot of Short-term Memory

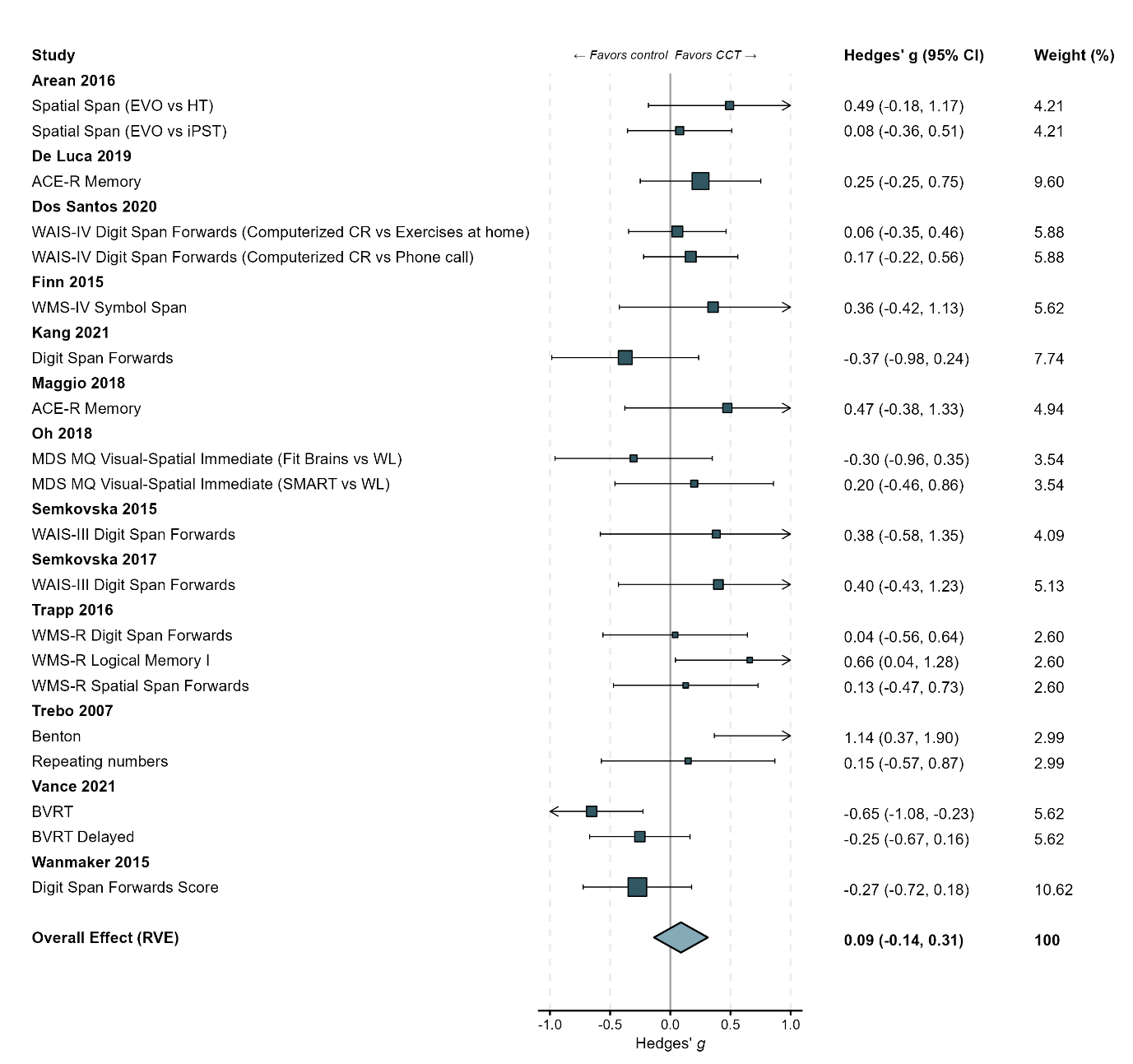

**eFigure 31.** Funnel Plot of Short-term Memory

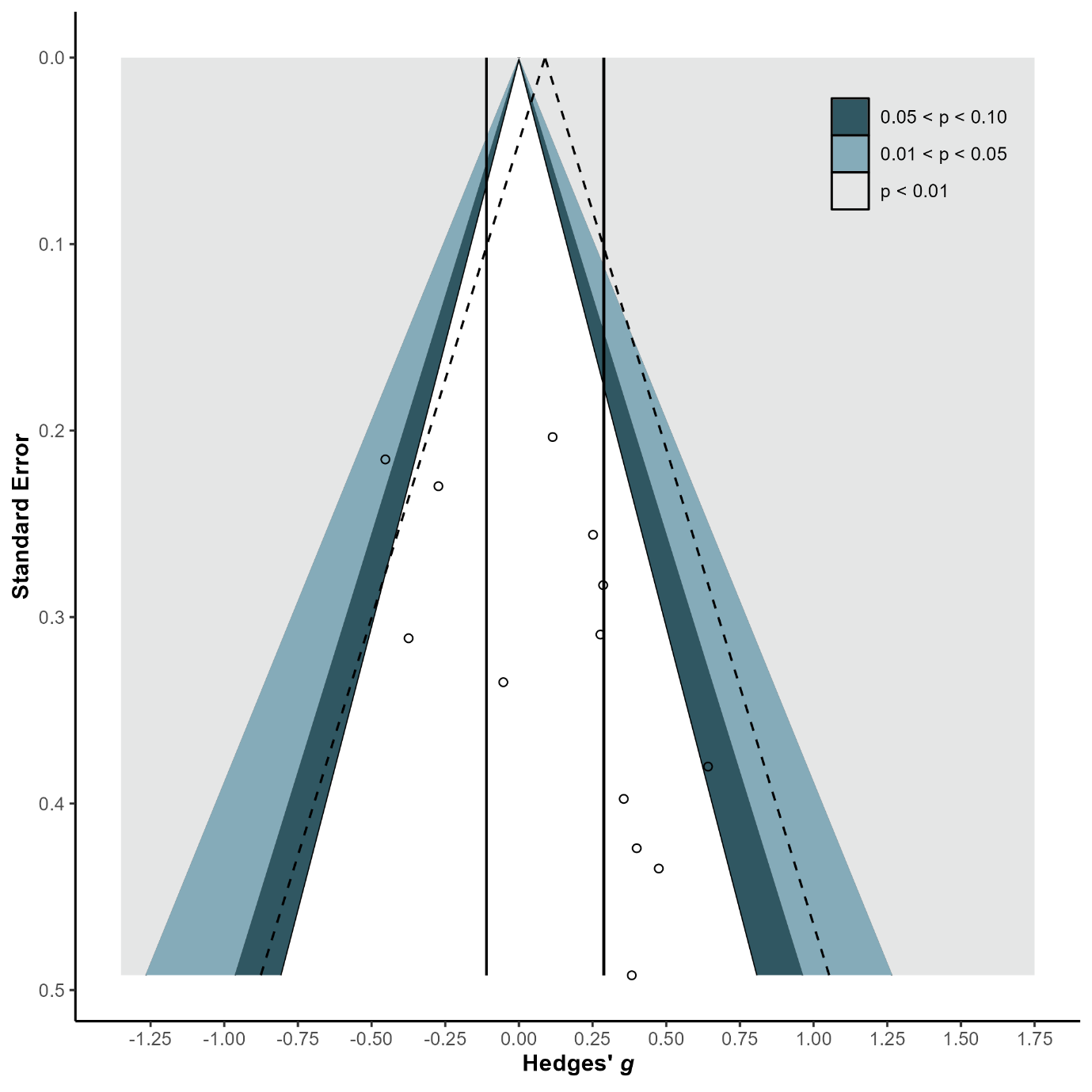

**eFigure 32.** Forest Plot of Executive Function

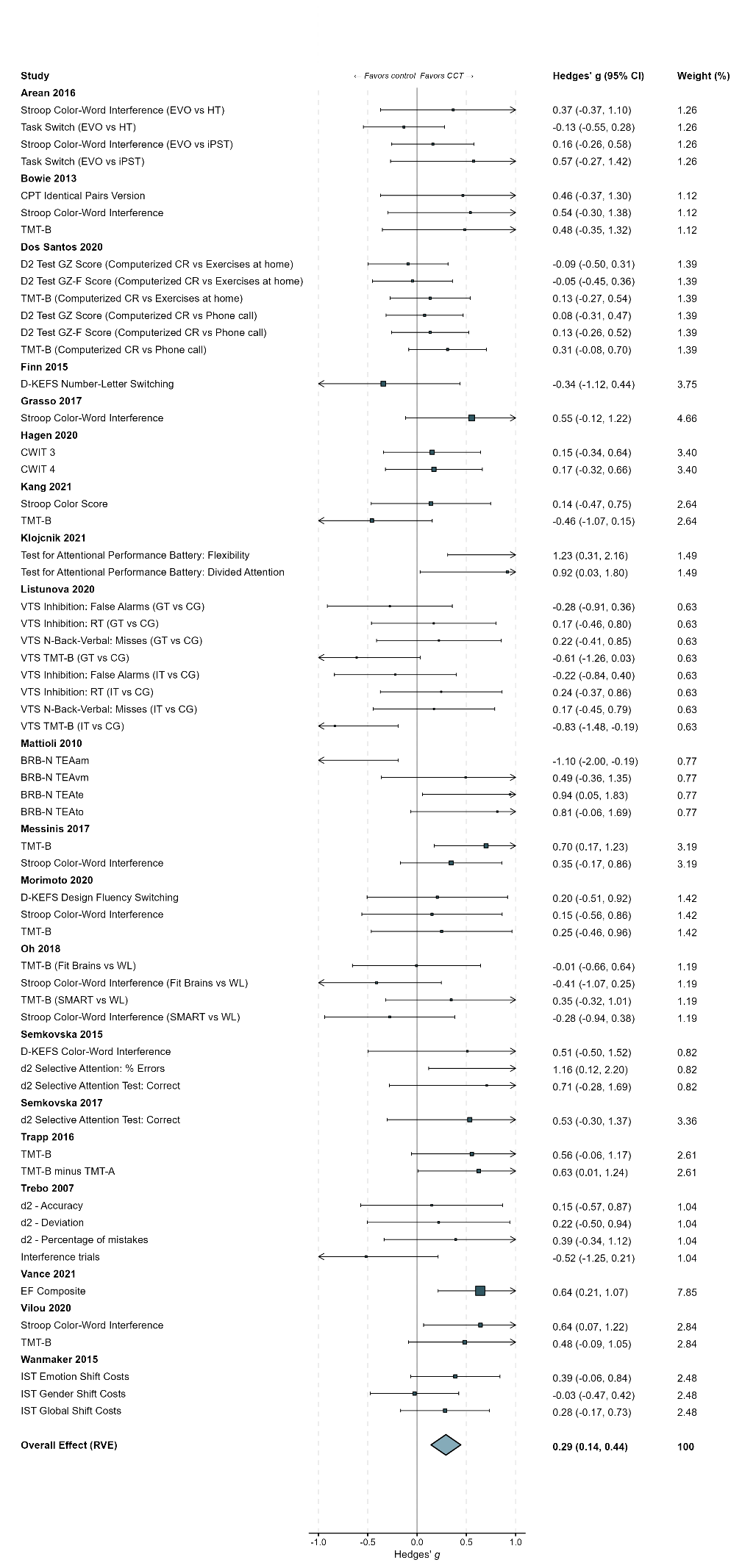

**eFigure 32.** Forest Plot of Executive Function…continued…

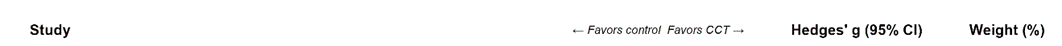

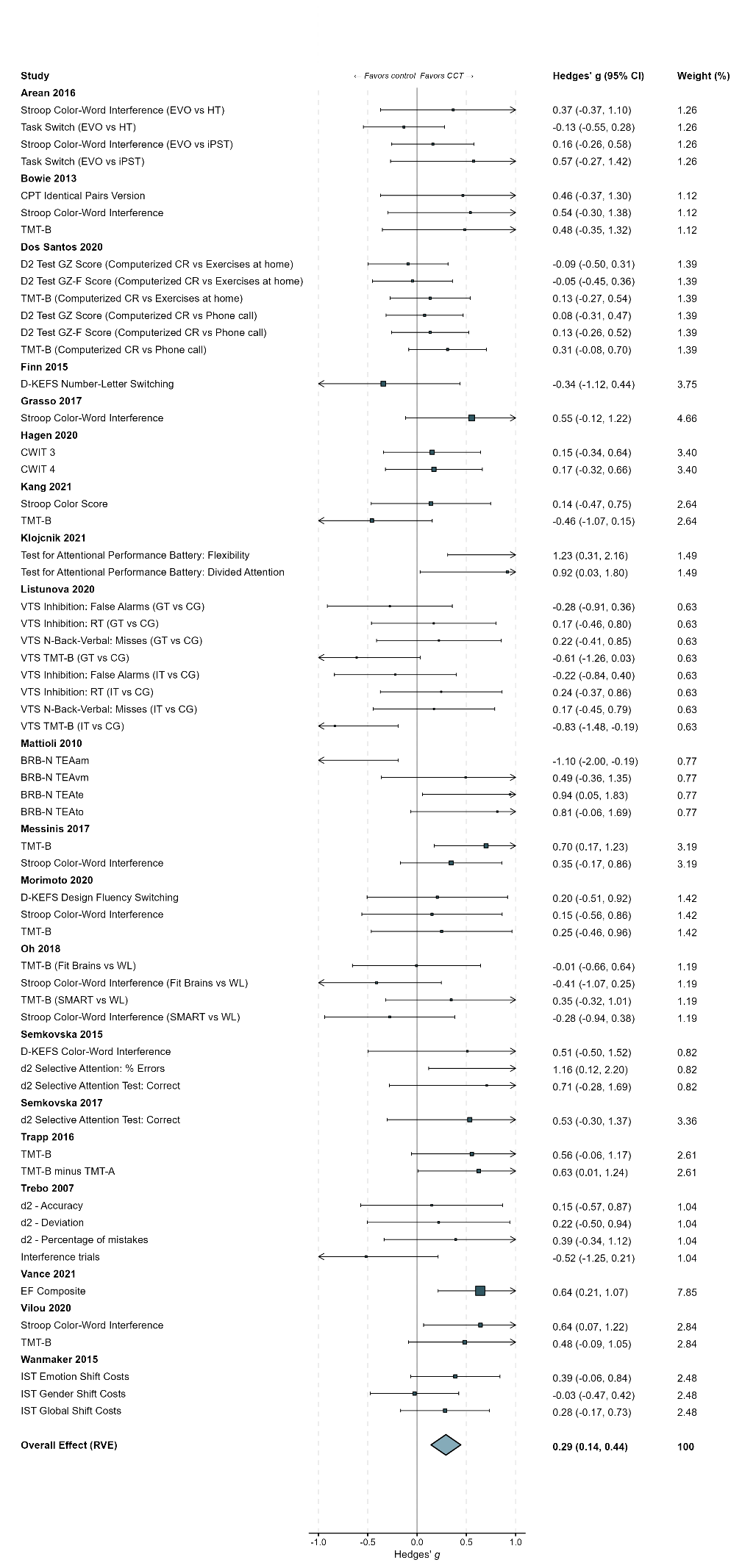

**eFigure 33.** Funnel Plot of Executive Function
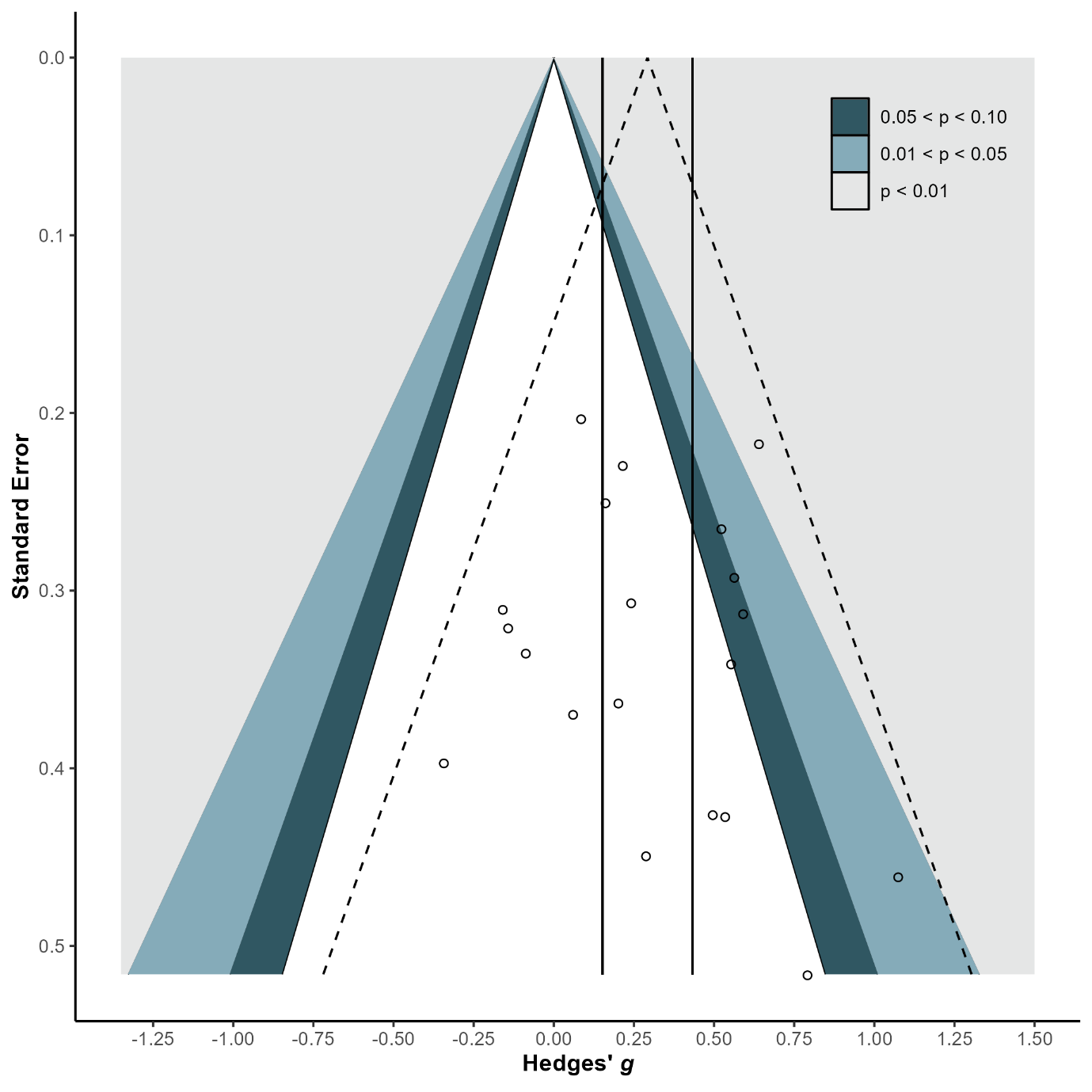

**eFigure 34.** Forest Plot of Shifting

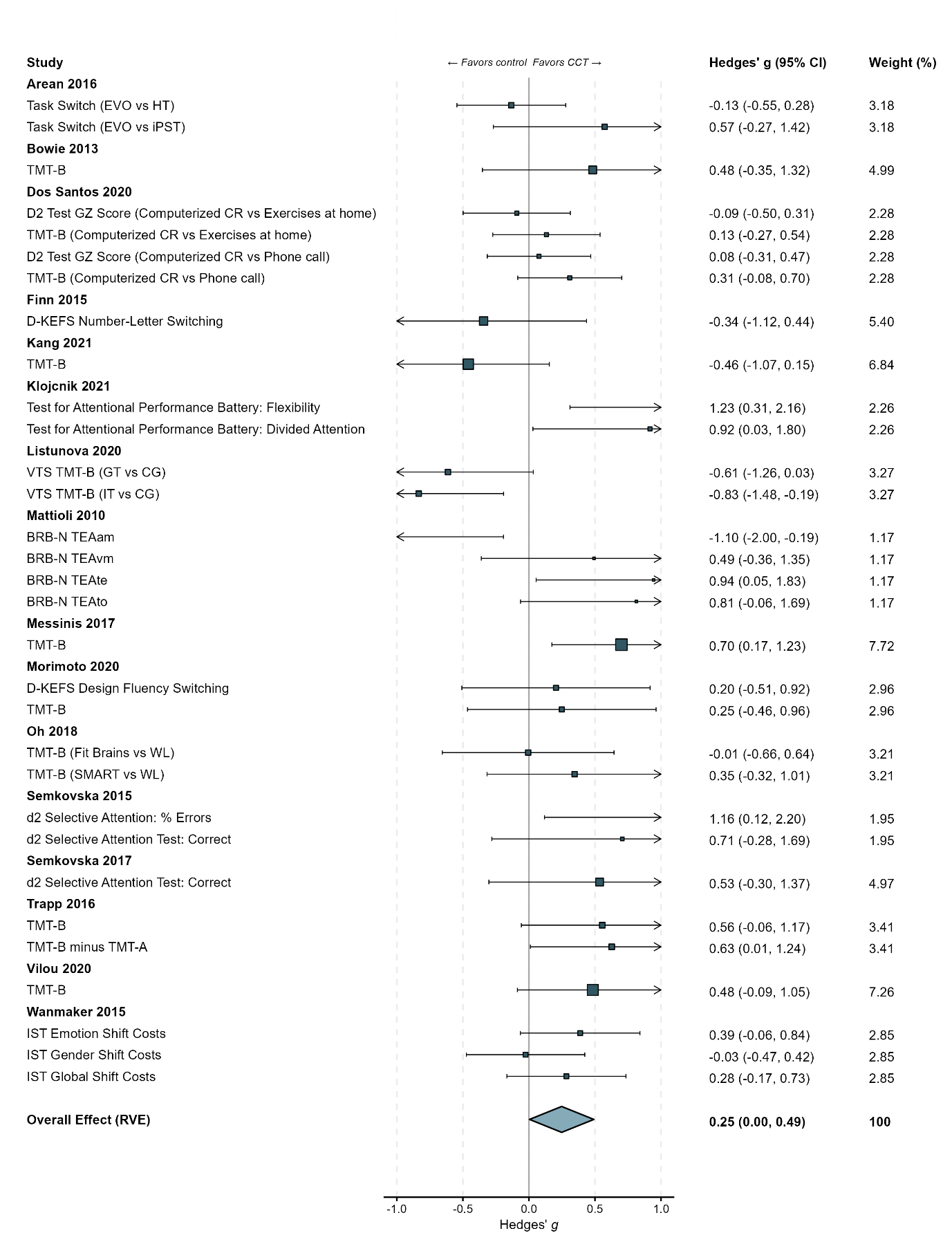

**eFigure 35.** Funnel Plot of Shifting

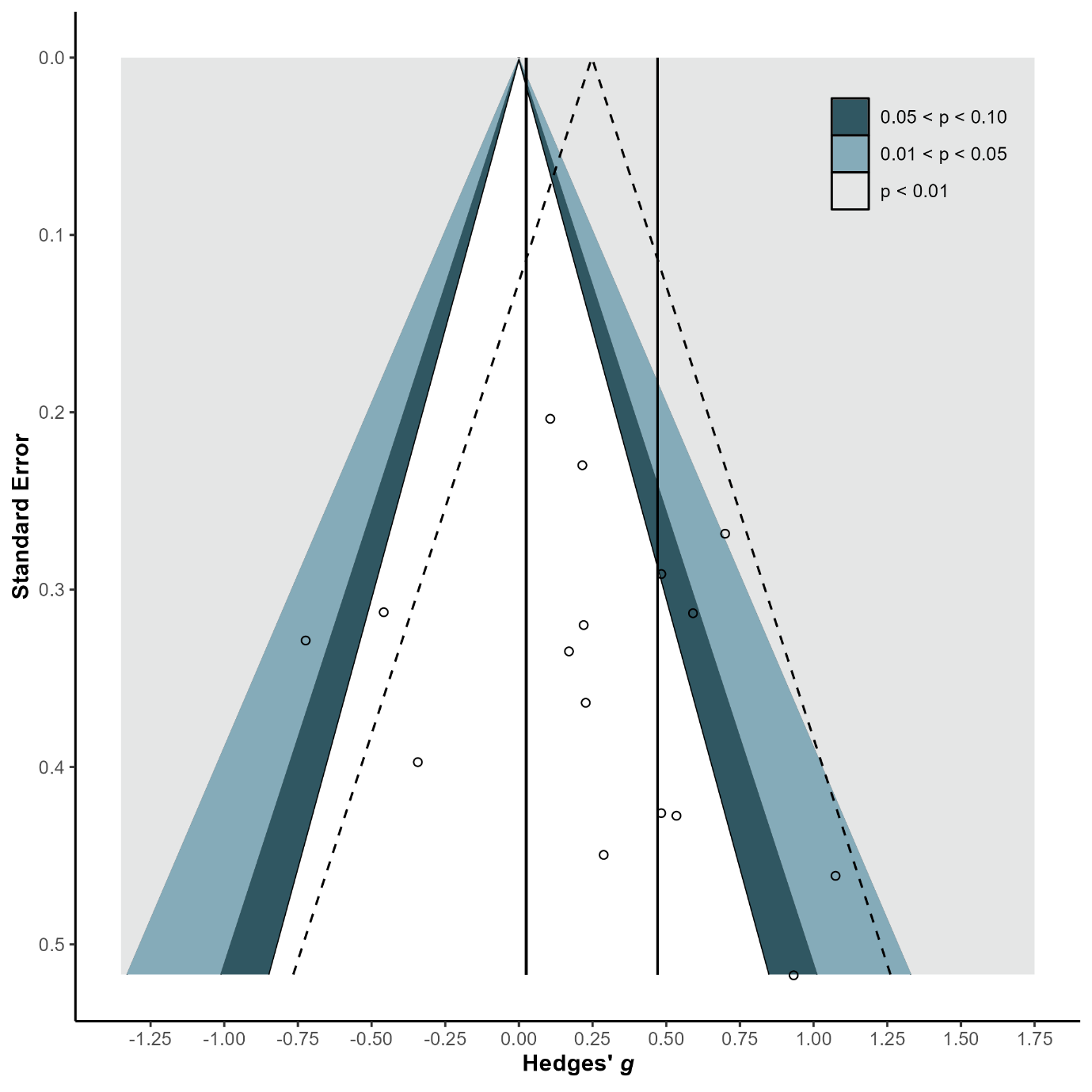

**eFigure 36.** Forest Plot of Inhibition

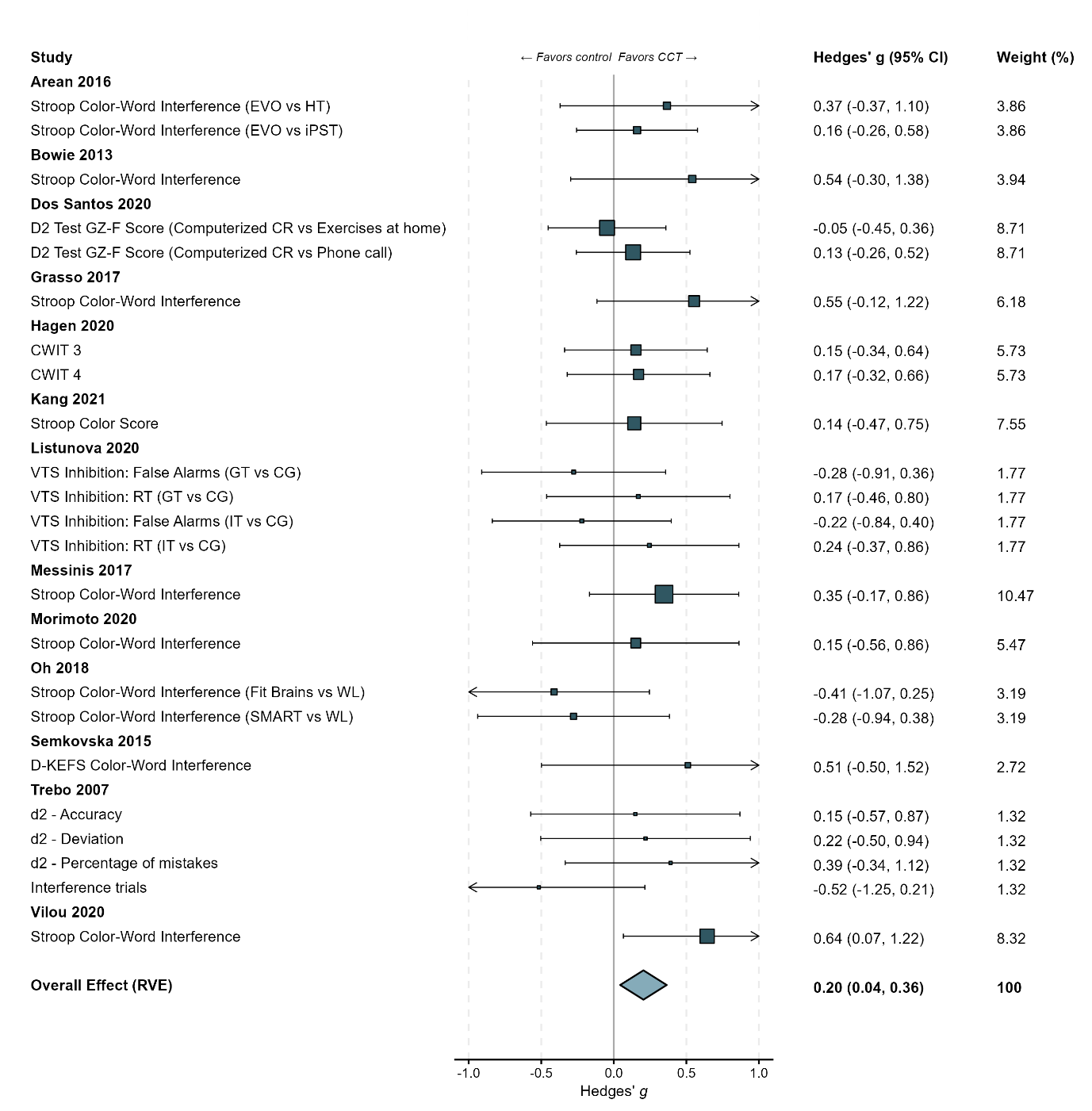

**eFigure 37.** Funnel Plot of Inhibition

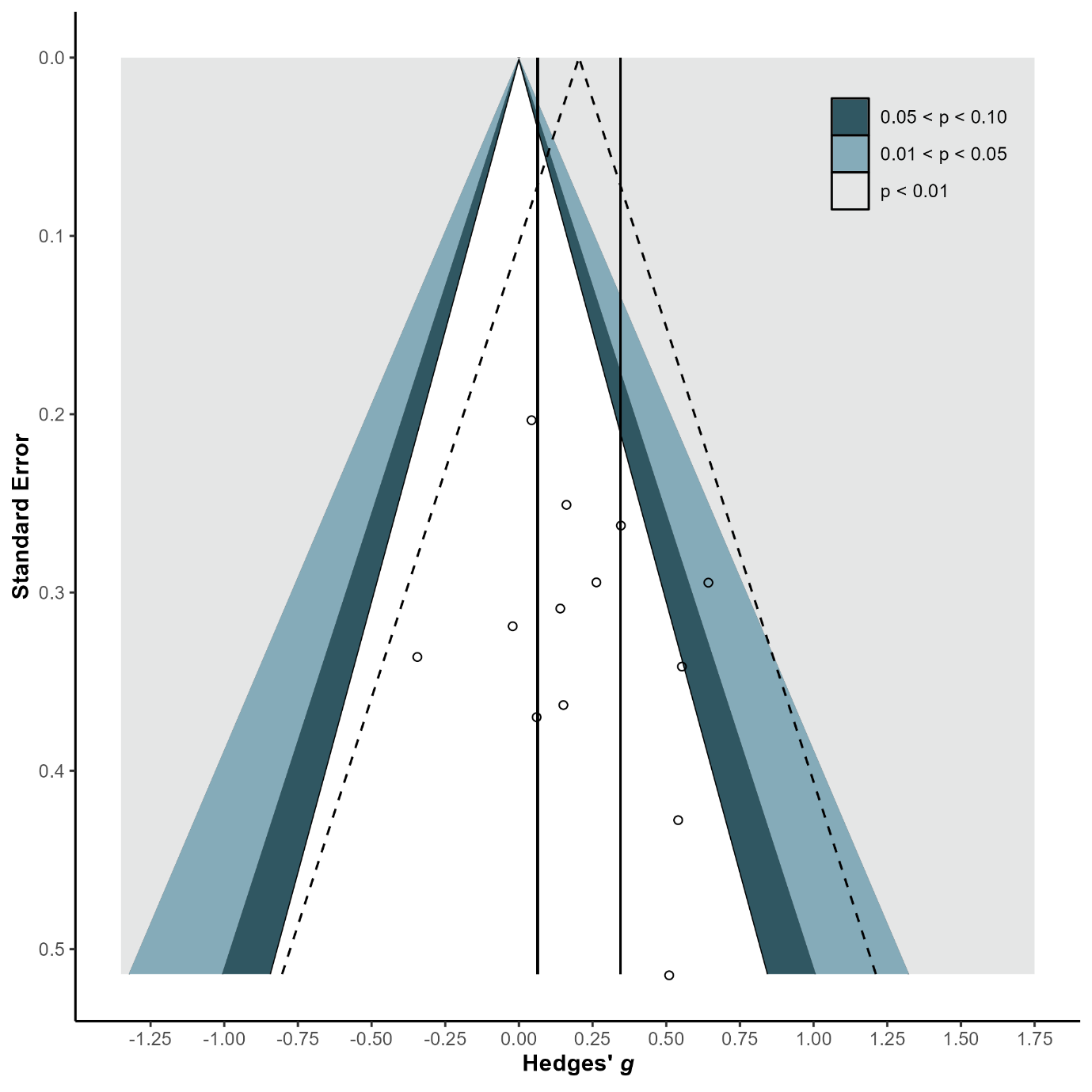

**eFigure 38.** Forest Plot of Processing Speed

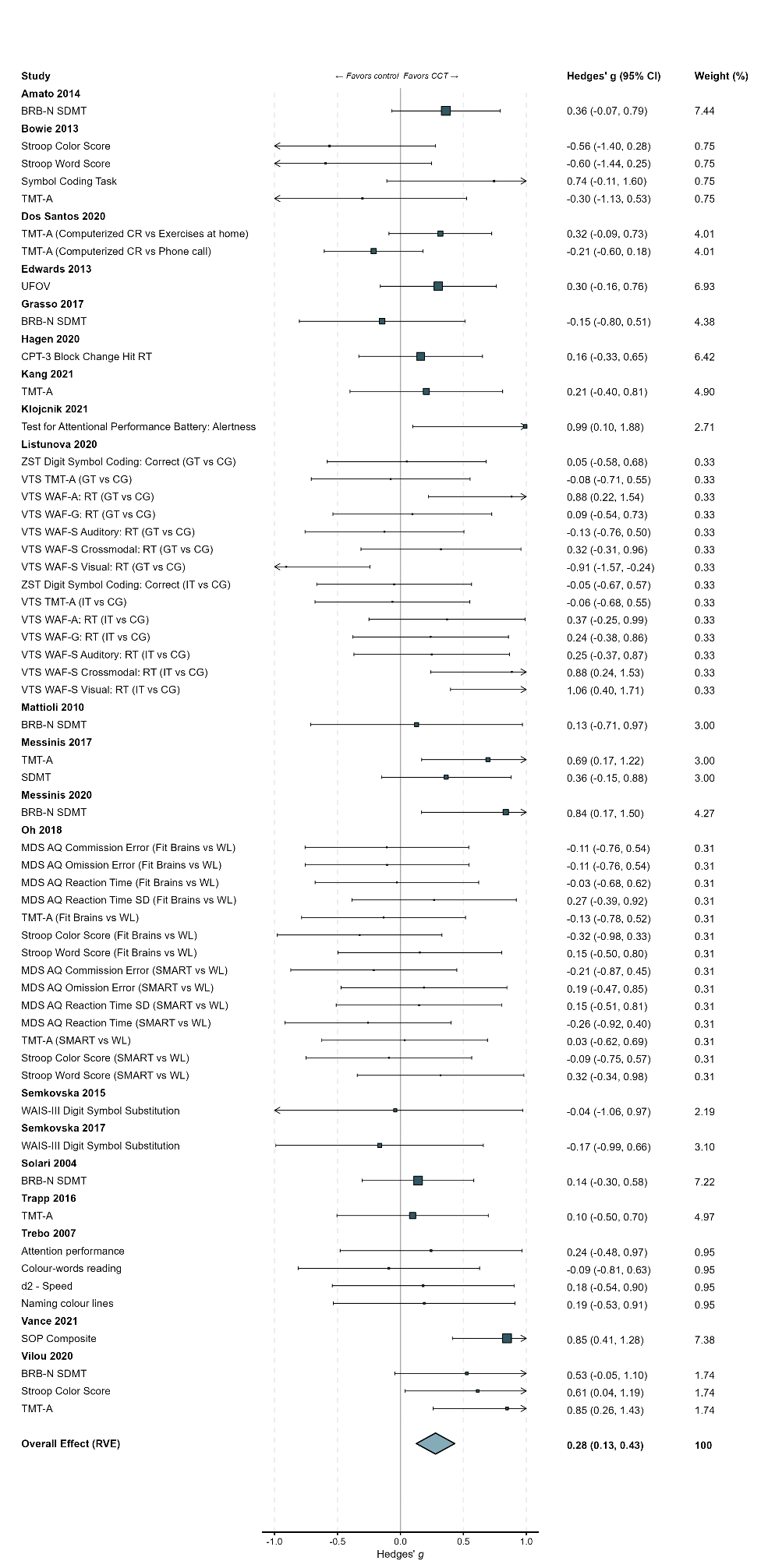

**eFigure 38.** Forest Plot of Processing Speed…continued…

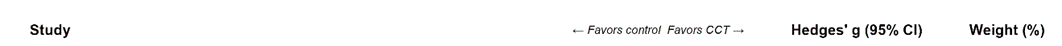

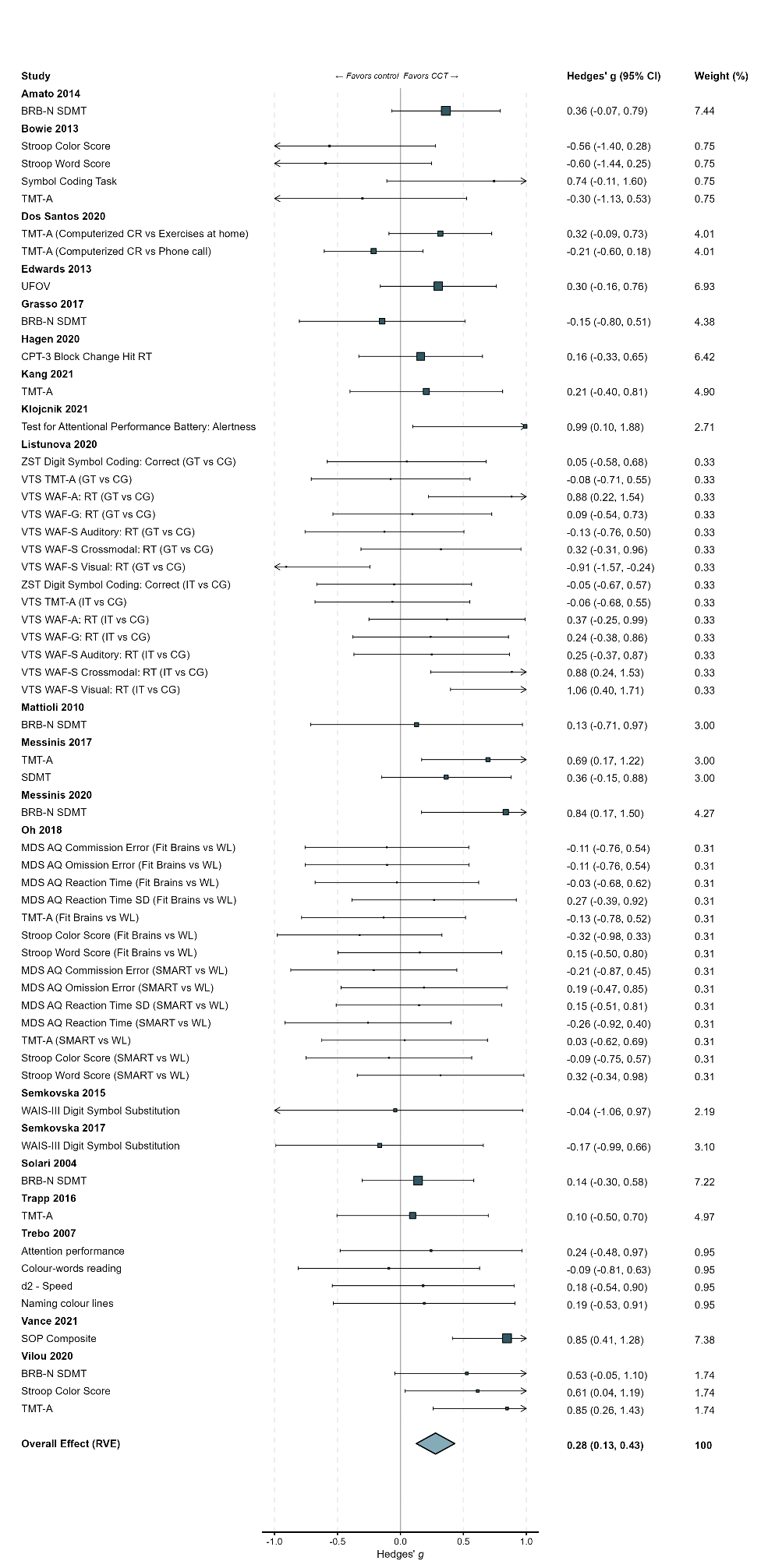

**eFigure 39.** Funnel Plot of Processing Speed

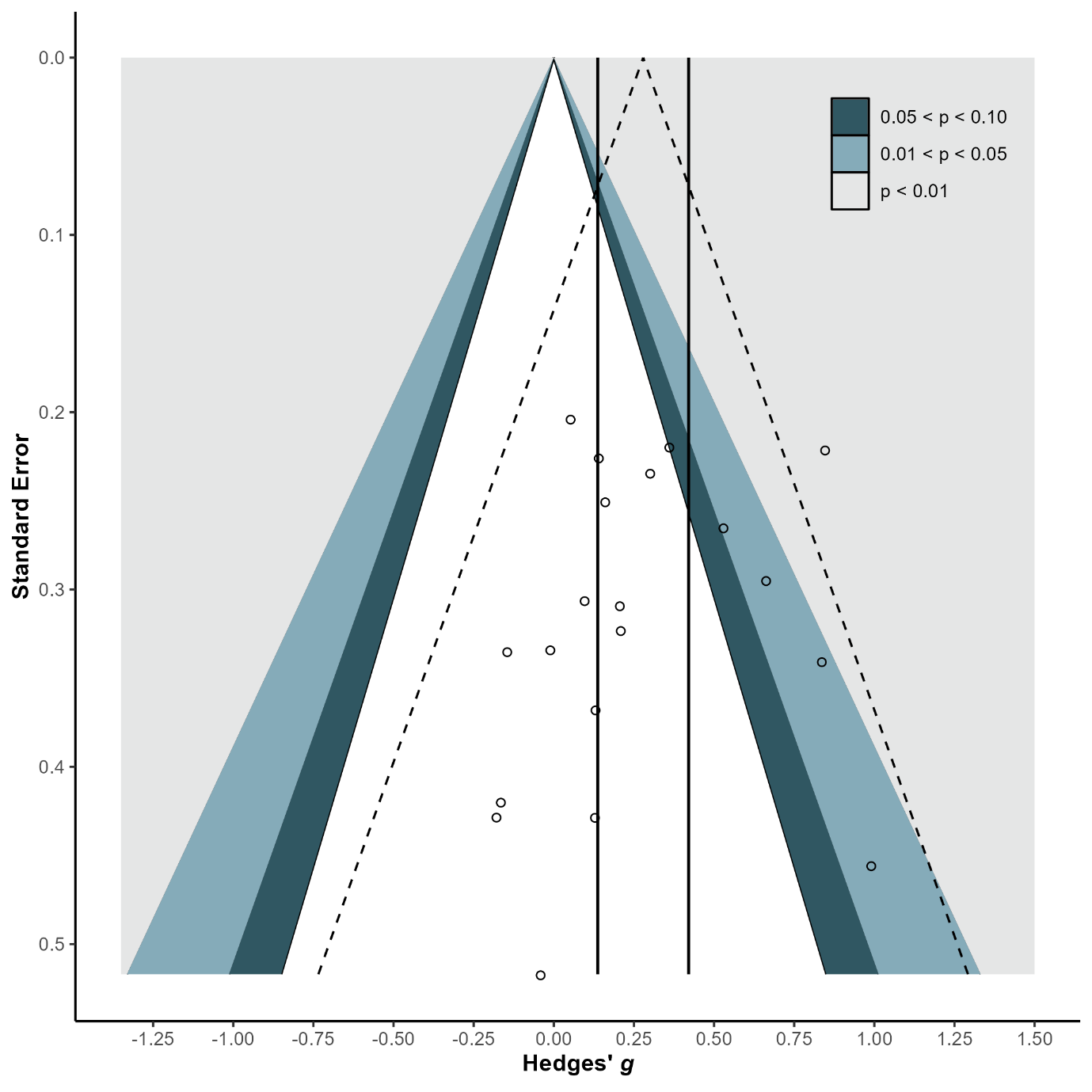

**eFigure 40.** Forest Plot of Perceptual Speed

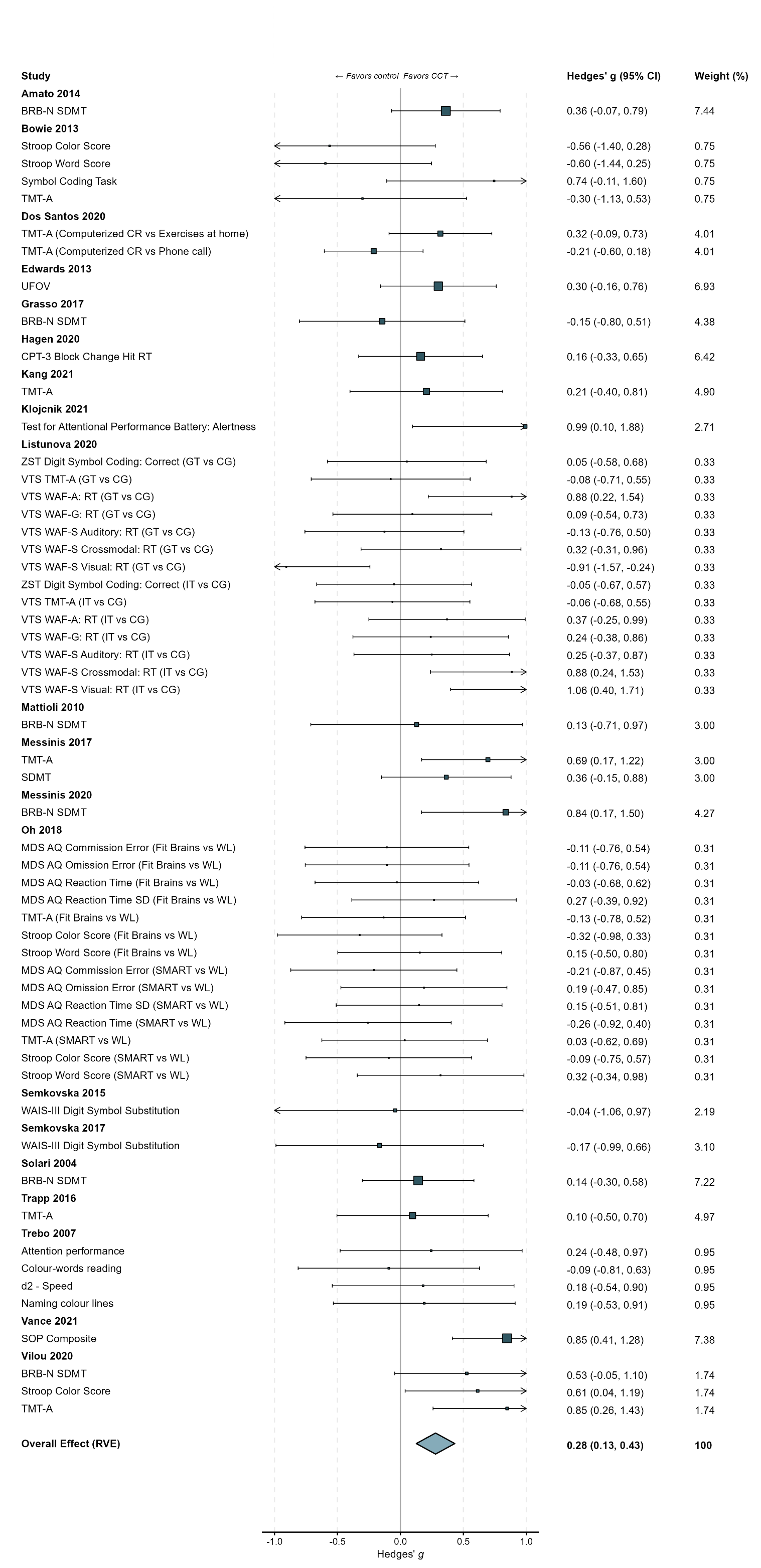

**eFigure 40.** Forest Plot of Perceptual Speed …continued…

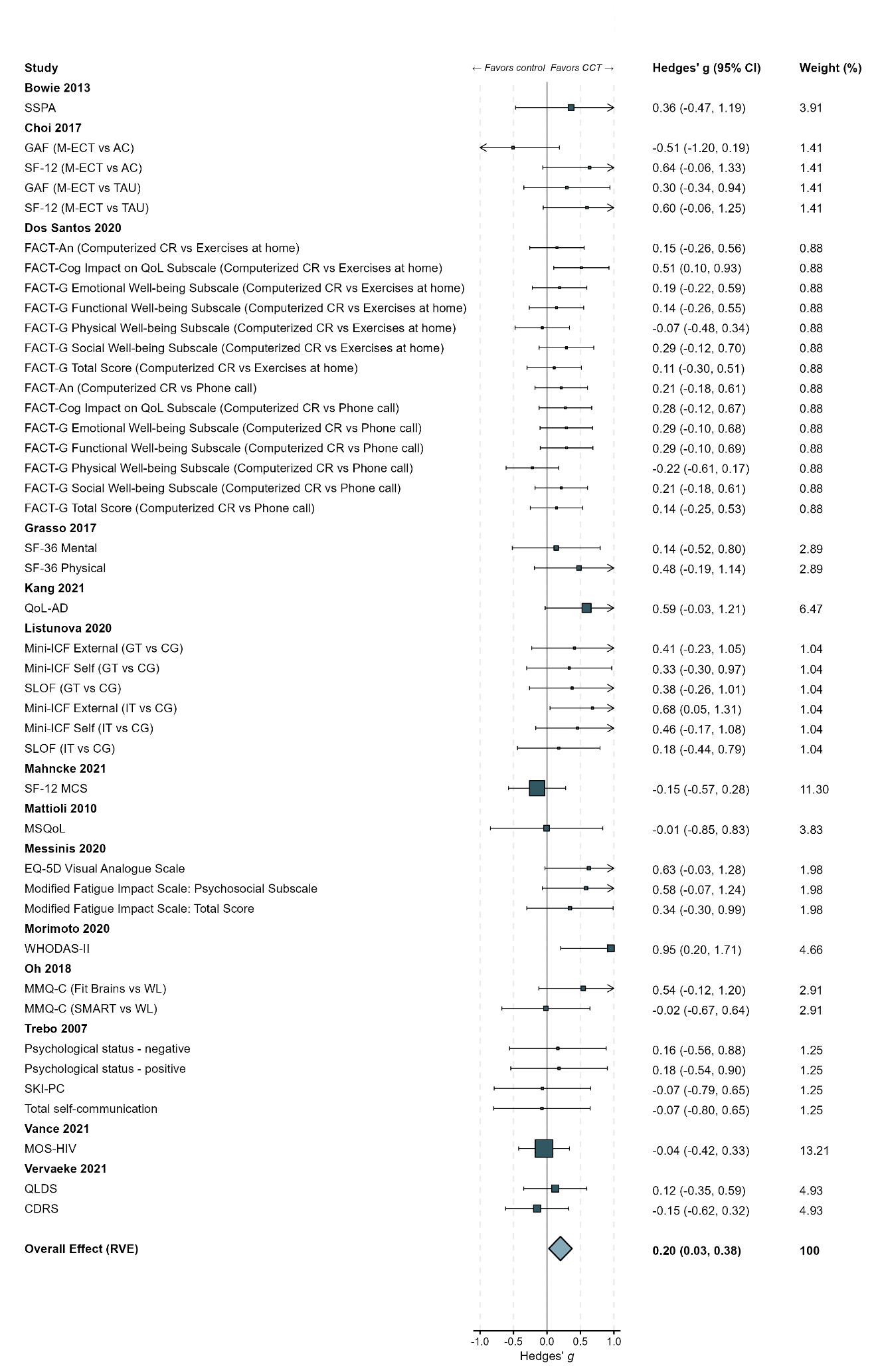

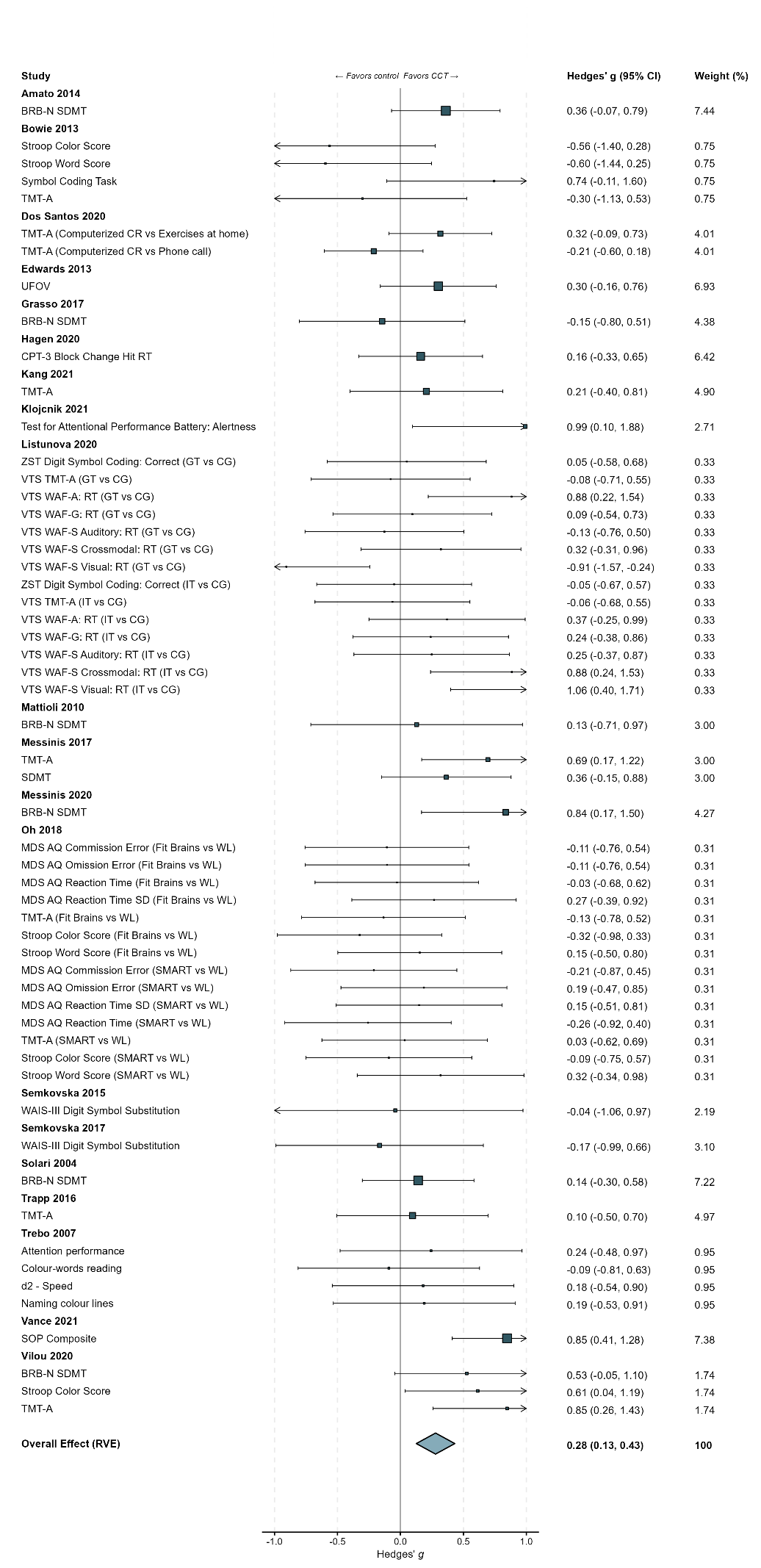

**eFigure 41.** Funnel Plot of Perceptual Speed
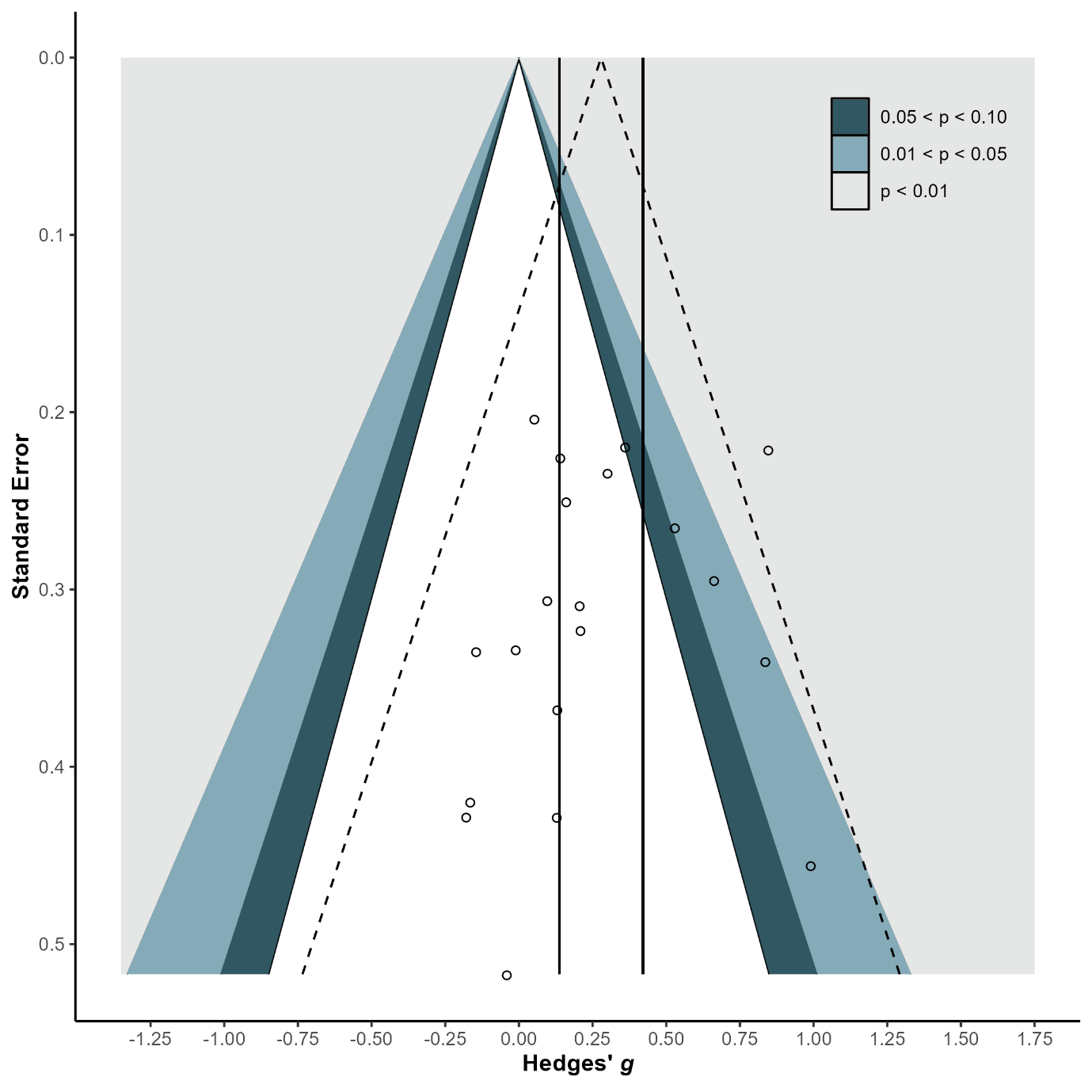

**eFigure 42.** Forest Plot of Visual Processing

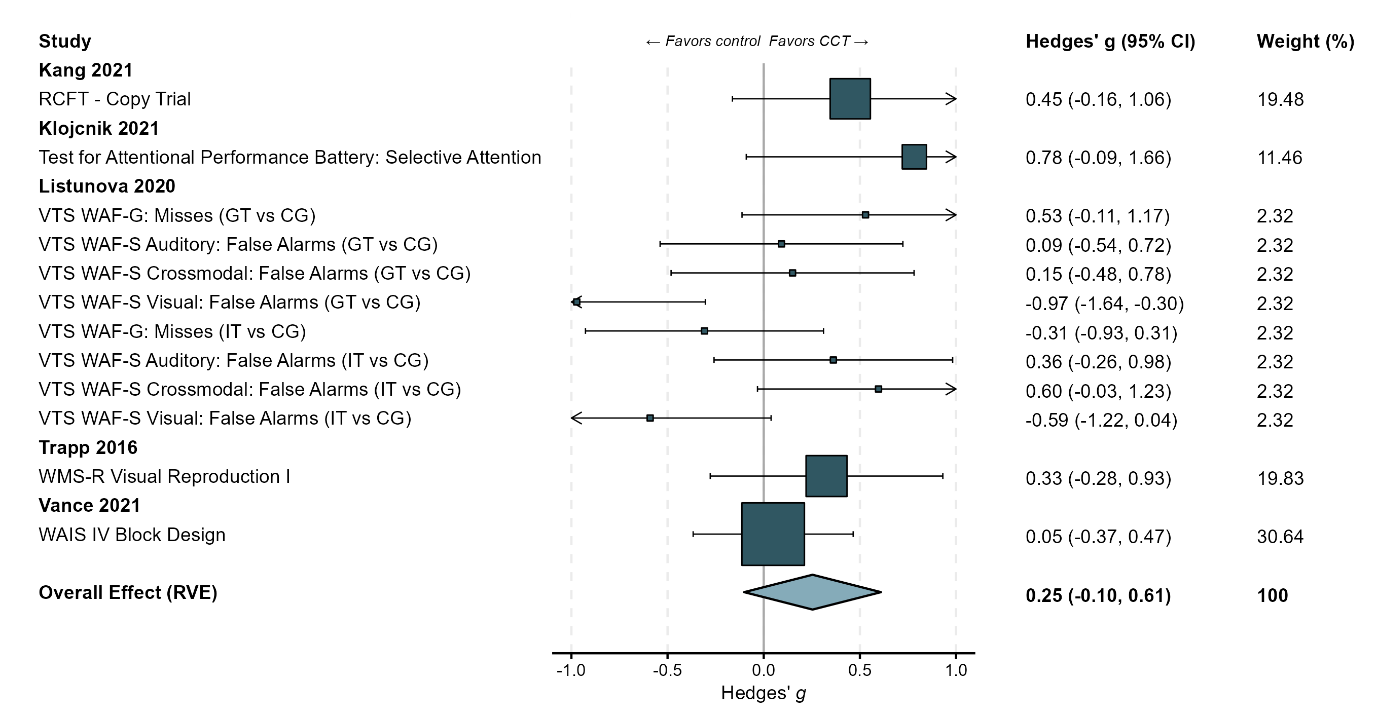

**eFigure 43.** Funnel Plot of Visual Processing

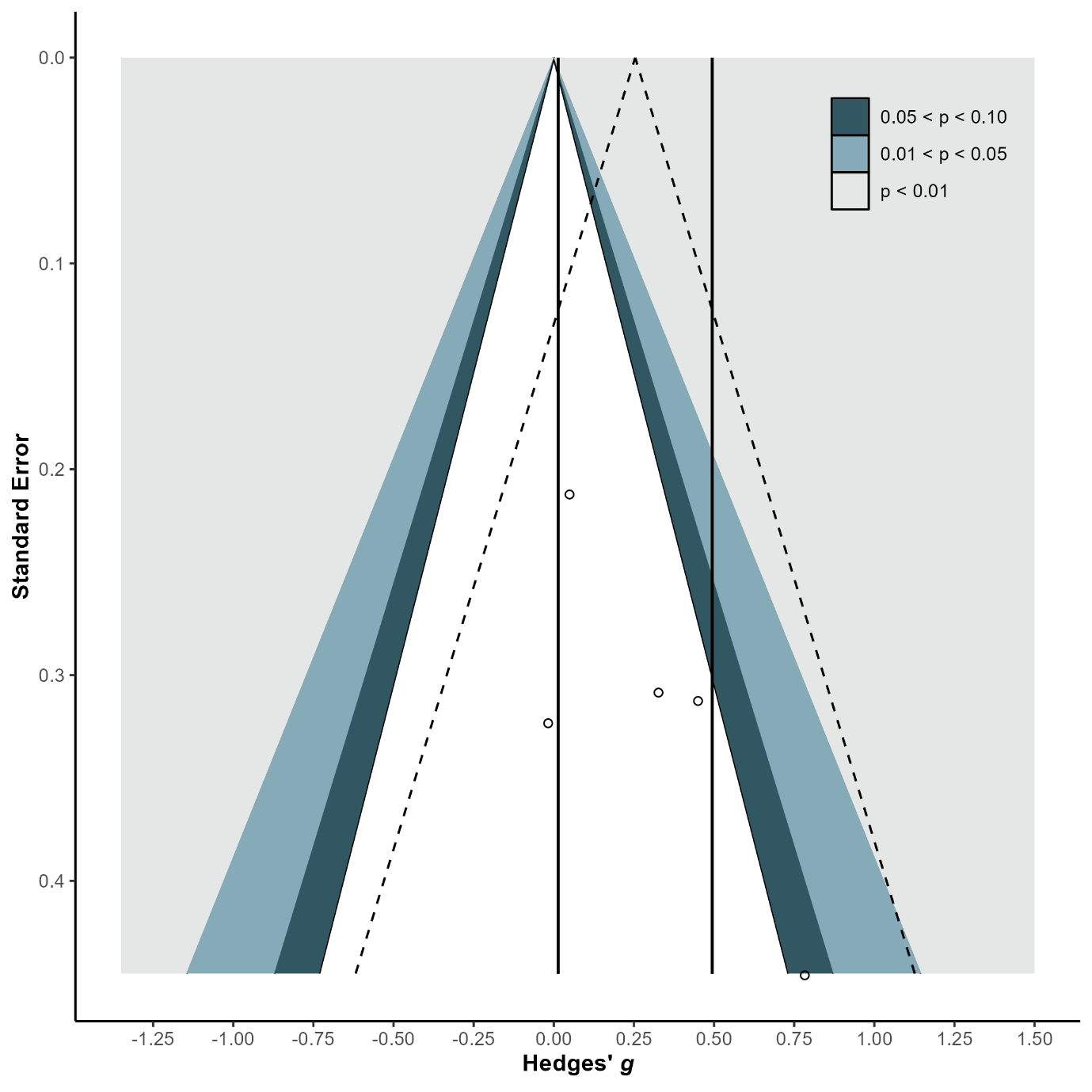

**eFigure 44.** Forest Plot of Sensory Perception

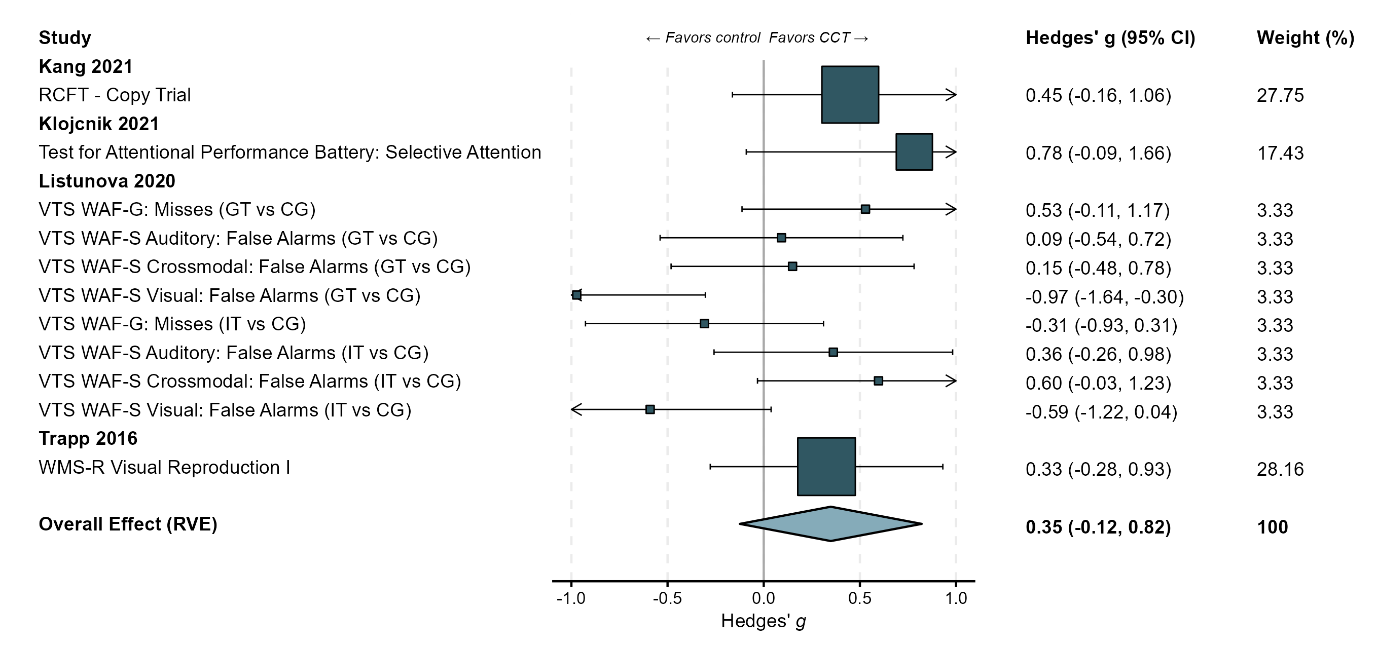

**eFigure 45.** Funnel Plot of Sensory Perception

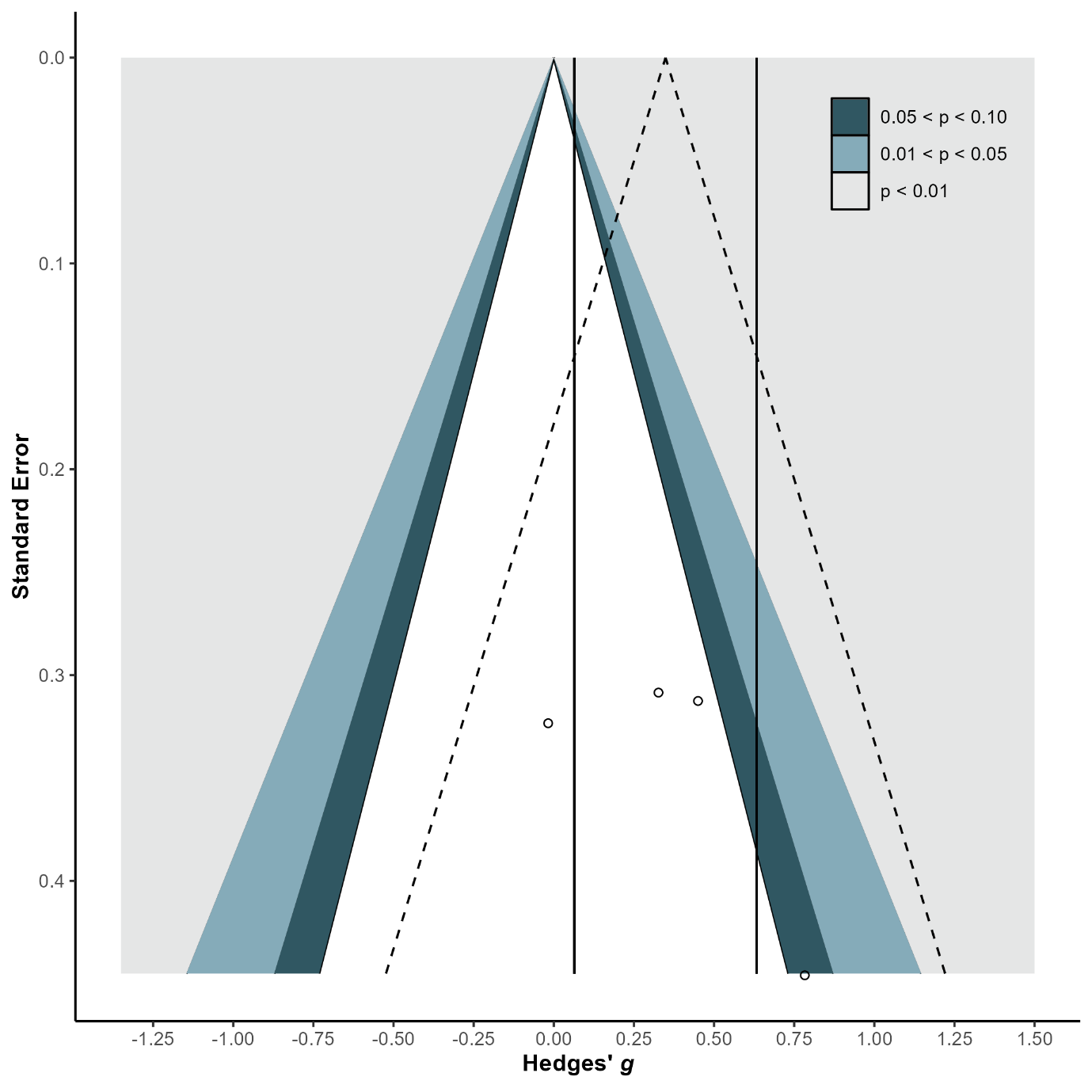
